# Multi-ancestry fine-mapping of the chromosome 17q12-q21 asthma locus identifies independent associations implicating lymphocyte and eosinophil levels in the causal pathway

**DOI:** 10.1101/2025.08.05.25333026

**Authors:** Chief Ben-Eghan, Markus Münter, Chikashi Terao, Alex Diaz-Papkovich, Simon Gravel, G. Mark Lathrop, Audrey V. Grant

## Abstract

**Background:** Asthma pathophysiology varies by age-of-onset and involves diverse immune processes reflected in white blood cell (WBC) subsets. To investigate the genetic architecture of asthma and potential endophenotypes, we analyzed the chr17q12-q21 locus, a robustly replicated asthma locus, across European (EUR), African (AFR), East Asian (EAS), and South Asian (SAS) ancestry groups from the UK Biobank (UKB) and Biobank Japan (BBJ). The largest EUR sample was further stratified by age-of-onset as a proxy for etiological heterogeneity.

**Results:** Eight independent asthma signals were identified in UKB-EUR, including two novel associations (Signal 2—rs72832915 and Signal 6—rs507671). Signal 4, corresponding to the originally identified pediatric signal, showed the strongest cross-ancestry evidence, with asthma risk and diminished lymphocyte count co-localizing in three populations. Signal 8 was distinguished by multiple lines of evidence converging on rs112401631 as a likely causal variant, including fine-mapping, colocalization with eosinophil and lymphocyte counts, and mediation of asthma risk through eosinophil count. Signal 6 implicated *RARA* expression, suggesting vitamin A metabolism impacts on late-onset asthma, the only associated stratum. Notably, integrating WBC traits and Bayesian fine-mapping enabled leveraging non-European ancestry groups to strengthen causal inference despite smaller sample sizes.

**Conclusion:** These findings illustrate how combining age-of-onset stratification, quantitative endophenotypes, and multi-ancestry analyses can reveal mechanistic heterogeneity and prioritize specific variants and pathways for functional validation. This framework is broadly applicable to complex diseases with measurable quantitative endophenotypes.

## 1 Introduction

Asthma is a complex, chronic disorder characterized by bronchial inflammation and narrowing of the airways that affects an estimated 300 million people worldwide [1]. Prevalence of asthma differs across the globe and by ancestry, with distinct molecular and etiological features underlying childhood-adult– and late-onset asthma [2]. The 17q12-q21 locus was first identified in a European descent (EUR) genome-wide association (GWA) study for childhood asthma [3, 4] and stands out as the most highly replicated asthma locus in EUR [5] and Asian descent populations [6, 7, 8]. It is composed of several independent signals located in a 150 kb core region with high pairwise linkage disequilibrium (LD) and in flanking proximal and distal regions [5]. Many of the variants originally identified in European populations occur at lower frequencies in African descent populations, which can reduce statistical power for association testing in these groups [9, 10]. The core region signal has also been detected in multi-ancestry meta-analyses across continental populations including the TAGC [11] and EVE consortia [12]. Each sub-region has been associated with differential expression of several genes: *ORMDL3* and *GSDMB* (core), *GSDMA* (proximal) and *PGAP3* (distal) [5]. So far, fine-mapping efforts have not clearly related asthma phenotypic heterogeneity to independent association signals in the region, nor has causal implication of specific endophenotypes and genes been established on a signal-specific basis.

Levels of white blood cell (WBC) subtypes reflect immune health status [13], and particular profiles may designate different forms of asthma [14, 15]. Specifically in asthma pathophysiology, exogenous allergens are detected by airway epithelial cells and sampled by dendritic cells, which activate lymphocytes that, in turn release cytokines driving the recruitment and activation of inflammatory effector cells–including eosinophils, as well as mast cells, basophils, neutrophils, monocytes, and macrophages [15]-thereby implicating several WBC subtypes in the lung. Recent GWA studies of basal whole blood WBC traits have resulted in the identification of thousands of genome-wide significant loci [16] including one study across 5 global populations [17]. Mendelian Randomization has shown evidence of causal association across specific WBC subtypes and asthma [16], and trans-ancestral meta-analysis has pointed to pleiotropy and colocalization of WBC subtypes with immune-related diseases [18]. Specific genetic variants underlying asthma risk may also impact WBC subtype distribution, with specific subtype levels mediating asthma genetic associations.

To better understand the contribution of the 17q12-q21 locus to asthma etiology, we adopted a multi-pronged strategy leveraging asthma and WBC subtype count data from the UK Biobank (UKB) across four ancestry groups (European, African, East Asian and South Asian or EUR, AFR, EAS, SAS) and Biobank Japan (BBJ) incorporating age-of-onset stratification in the largest UKB-EUR subset. To enhance power in non-EUR populations, we used quantitative WBC traits to identify immune-relevant signals, which were then integrated with asthma associations to characterize region-specific genetic architecture. While this does not fully overcome the lower power of asthma association testing, it facilitates interpretation of shared or distinct signals across phenotypes and ancestries. By integrating recent statistical genomics advances in Bayesian fine-mapping and colocalization, and summary statistic Mendelian randomization [19], our framework moves toward signal-specific causal pathway construction, highlighting combinations of WBC subsets and differentially expressed genes that may underlie asthma risk across ancestries and age-of-onset groups.

## 2 Results

### 2.1 Study population and characteristics

We included individuals from the UK Biobank (UKB) and Biobank Japan (BBJ), encompassing five ancestry groups: European (UKB-EUR), African (UKB-AFR), South Asian (UKB-SAS), East Asian (UKB-EAS), and Japanese (BBJ). The UKB-EUR subset was defined as individuals who self-identified as White British (*N* = 408,963), to reduce potential confounding due to population stratification [20] while retaining high statistical power. In BBJ, 161,280 participants with asthma status and WBC data were included.

For non-European UKB participants, ancestry was assigned using unsupervised clustering via UMAP-HDBSCAN [21] on genetic principal components. This machine-learning approach uses UMAP to process the full principal component space, with HDBSCAN identifying cluster patterns for population structure analysis. This approach allowed for the inclusion of admixed or underrepresented groups often excluded under self-report criteria, thereby increasing statistical power. The final sample included 10,065 UKB-AFR, 9,366 UKB-SAS, and 2,549 UKB-EAS individuals (**Figures S1-S2; Tables S1-S2; Supplementary Materials: Table S1**). The effectiveness of this UMAP-HDBSCAN approach for population subset definition was validated through genomic inflation factors in subsequent association analyses (see **Methods** section in the **Supplementary Materials**; **Figure S3**).

Asthma cases in the UKB were defined based on self-reported doctor-diagnosed asthma. In the UKB-EUR subset, we stratified cases by age-of-onset into three mutually exclusive groups: childhood (*<*18 years; n = 13,419), adult (18-40 years; n = 12,526), and late (≥40 years; n = 18,056). We also defined an overlapping early asthma stratum (*<*40 years) to improve power for detecting genetic signals shared across younger-onset cases. BBJ used physician-diagnosed bronchial asthma from clinical records [22] (**Tables S1-S2**). Age distributions at recruitment and asthma onset across all UKB ancestry groups are shown in **Figure S4**.

In UKB, we evaluated eleven WBC traits as quantitative phenotypes: WBC total count, and absolute counts and percentages of neutrophils, eosinophils, basophils, monocytes, and lymphocytes, denoted NEU, EOS, BAS, MON and LYM, respectively. We used # to denote absolute counts and % for percentages, while only counts were available for BBJ.

Across all populations, EOS#, WBC# and NEU#-white blood cell counts reflecting innate immune activity—were elevated in asthma cases compared to controls (**Tables S3-S4**). WBC trait distributions varied by ancestry, with notably lower WBC# and NEU# in UKB-AFR, consistent with benign ethnic neutropenia [23], and higher EOS# and LYM# in UKB-SAS (**Figure S5**).

### 2.2 Asthma age-of-onset stratified association analysis

To identify independent asthma association signals at 17q12-q21, we performed comprehensive association testing followed by conditional and joint analysis (COJO) [24] across all five ancestry groups. Following quality control filtering, we analyzed 2,047-5,778 SNPs by ancestry group (minor allele frequency *>*1% within each group) across the 17q12-q21 region, which spans 37.2-38.8 Mb on chromosome 17. This region encompasses the canonical asthma locus core region (297 kb) as well as proximal and distal regions, extending a further 550 kb upstream and 750 kb downstream, defined based on linkage disequilibrium structure, base-pair distance, and p-value clustering patterns from age-of-onset stratified analyses in UKB-EUR [5, 25].

We applied COJO to each of the four UKB-EUR age-of-onset strata to identify independent signals while accounting for linkage disequilibrium. A complete overview of all analytical algorithms and their applications across ancestry groups is provided in **Supplementary Materials: Table S2**. The conditional association approach identified eight conditionally independent asthma-associated signals (linkage disequilibrium threshold *r*^2^ *<*0.5; P *<*1.0×10^−4^ [19]) across the four strata (**Figure 1**, **Figure 2a-d**; **Table 1**; **Tables S5-S6**). These eight signals form the foundation for all downstream analyses investigating links with WBC traits and eQTL evidence.

**Figure 1.**
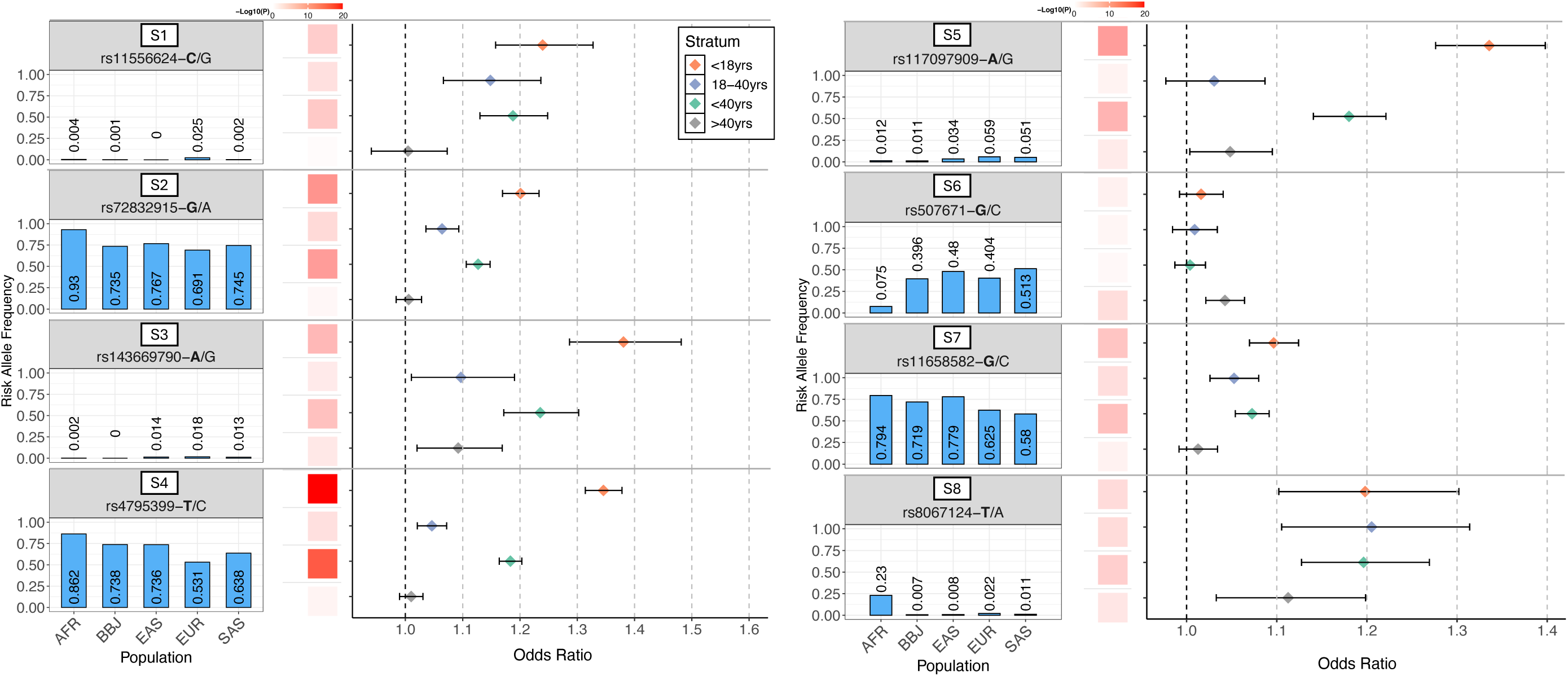
Independent asthma signals: population allele frequencies and odds ratios by age-of-onset strata. Eight conditionally independent asthma-associated signals (S1-S8), identified in the UK Biobank European ancestry subset using GCTA-COJO, are shown in a 4-row by 2-column grid. Each panel corresponds to one signal, represented by the variant displaying the strongest statistical significance across the four asthma strata (i.e., the variant with the lowest unadjusted P-value in GCTA-COJO analysis). Each includes three elements: a bar plot of risk allele frequencies across five global populations (AFR, BBJ, EAS, EUR, SAS), a heatmap of –log_10_(P) across four age-of-onset strata (*<*18 years (childhood), 18-40 years (adult), *>*40 years (late), *<*40 years (early) asthma), and a forest plot of odds ratios (ORs) with 95% confidence intervals by age-of-onset strata. Risk alleles (bolded) are oriented towards asthma risk, and colour intensity in the heatmap reflects statistical significance. –Log10(P) values were square root transformed and rescaled so that all values set at 20 displayed full intensity.

**Figure 2.**
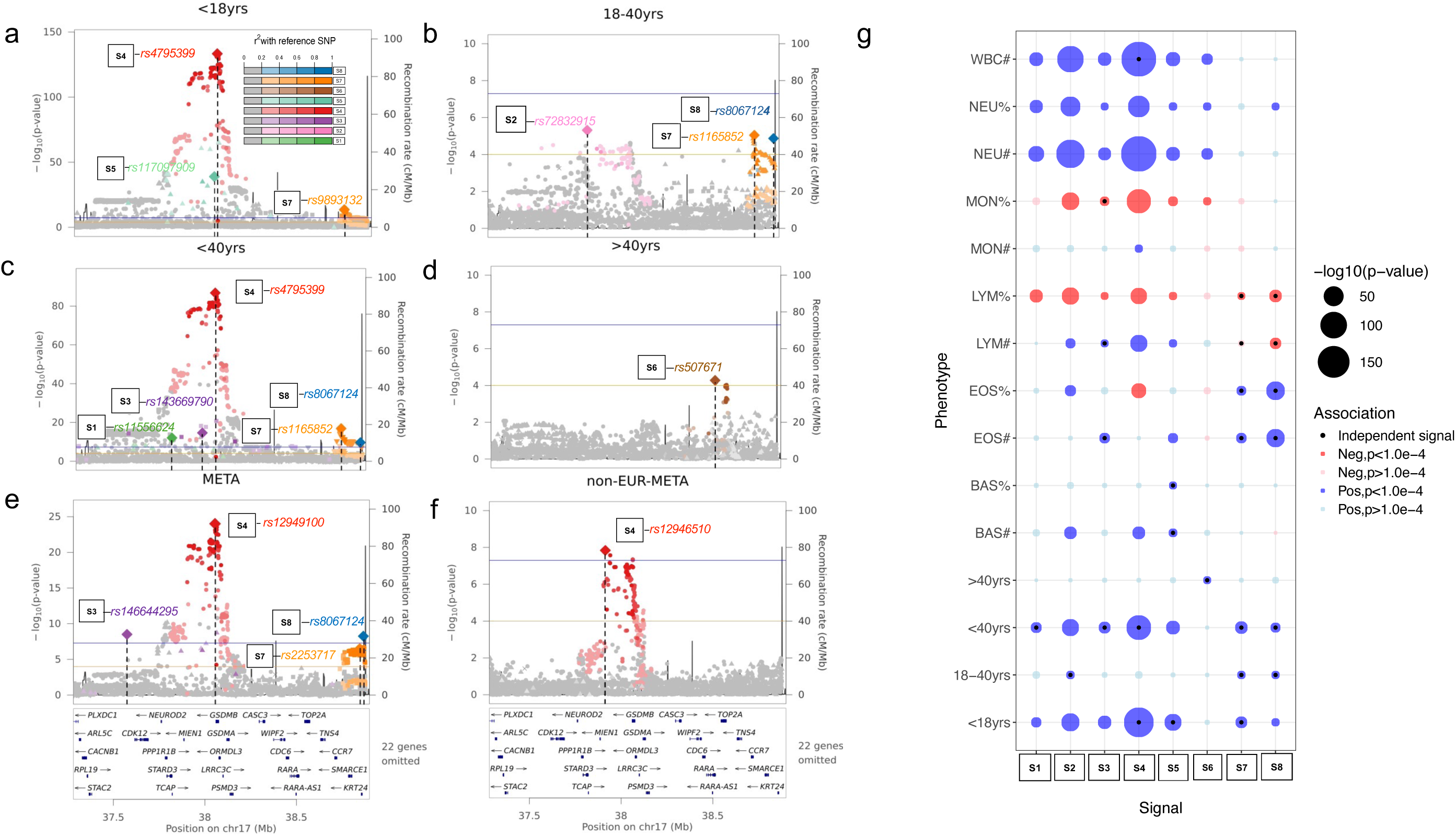
Independent asthma signals and their associations by asthma age-of-onset strata and with white blood cell traits. Regional plots for signals S1-S8, identified using GCTA-COJO in the UK Biobank European ancestry subset (UKB-EUR), are shown by asthma age-of-onset strata (*<*18 years (early), 18-40 years (adult), *<*40 years (early), and *>*40 years (late) asthma) in panels (a-d), and from meta-analyses of asthma adjusted for sex and age (asthma*_all_*) in all populations (EUR+AFR+SAS+EAS+BBJ); e) and in non-Europeans (AFR+SAS+EAS+BBJ); f). For all panels (a-f), each point represents a variant’s –log_10_(P) plotted against its chromosomal position (hg19); lead variants are marked with diamonds and vertical dashed lines, with neighbouring variants shaded by LD (*r*^2^) to the lead SNP (see key in a). Signal colour and shape are consistent across panels. Blue and orange lines indicate genome-wide (P = 5.0×10^−8^) and suggestive (P = 1.0×10^−4^) thresholds; recombination rates and gene annotations are displayed. (g) Bubble plot shows associations between each GCTA-COJO defined asthma signal and six white blood cell traits in UKB-EUR. Bubble size reflects –log_10_(P); colour denotes direction of association with the asthma risk allele (blue = positive, red = negative); black dots indicate independent signals with P *<*1.0×10^−4^.

**Table 1:** Summary of sources of causal evidence.

| Signal | SNP | Signal Resolution | Asthma | Literature |  | Conditional Association |  |  |  | Fine-mapping |  | Colocalization & Mediation |  | SMR |  |
| --- | --- | --- | --- | --- | --- | --- | --- | --- | --- | --- | --- | --- | --- | --- | --- |
|  |  |  | Age-of-onset | Asthma Independent | Pleiotropy only | META-Asthma | non-EUR META-Asthma | EUR WBC | non-EUR WBC | EUR Asthma | EUR WBC | EUR Asthma & WBC | non-EUR Asthma & WBC | Implicated Genes | Other functional |
| 4 | rs4795399 | NR | childhood | ✓ |  | ✓ | ✓ | WBC | LYM, WBC | ✓ | EOS, LYM, MON, NEU, WBC | LYM, MON, NEU, WBC | LYM | †IKZF3, †ZPBP2, †ORMDL3, †GSDMB, MED24, PGAP3, GSDMA, RP11-387H17.4 | ✓ |
| 8* | rs112401631 | R | adult | ✓ |  | ✓ |  | EOS, LYM |  | ✓ | EOS, LYM, NEU | **EOS, LYM** |  |  | ✓ (IKZF2) |
| 5 | rs117097909 | R | childhood |  |  |  |  | BAS |  | ✓ | EOS | EOS |  | PGAP3 |  |
| 3* | rs146644295 | R | early | ✓ |  | ✓ |  | EOS, LYM, MON |  | ✓ |  |  |  |  | ✓ (ZNF362) |
| 7* | rs9893132 | NR | childhood, adult, early | ✓ |  | ✓ |  | EOS, LYM |  | ✓ | EOS |  |  | SMARCE1, KRT24 |  |
| 1 | rs11556624 | R | early |  |  |  |  |  |  |  |  |  |  |  |  |
| 2 | rs72832915 | R | adult |  | ✓ |  |  |  |  |  |  |  |  | PGAP3 |  |
| 6 | rs507671 | R | late |  | ✓ |  |  |  |  |  |  |  |  | RARA,RARA-AS1 |  |

*Note:* Signals are sorted in descending order based on hierarchy of causal evidence from analyses conducted; WBC indicates White blood cell traits; \*Signals with variants showing stronger causal evidence of causality than their LD proxies, as identified using GCTA-COJO are denoted by an asterisk; NR indicates signals where a single causal variant could not be resolved; R designates signals with one variant, which is considered putatively causal based on the summary of its causal evidence over its proxies; EOS highlighted in **bold and underlined** under the colocalization & mediation column, indicates evidence from both analyses for S8–rs112401631, all other WBC traits highlighted under this column are from colocalization analysis only. For age-of-onset, < 18yrs, 18–40yrs & > 40yrs refer to childhood, adult and late-onset respectively; early-onset indicates signals identified in the < 40yrs. Ticks (✓) indicate which asthma signal was implicated in the respective analysis; WBC traits are designated using the first 3 letters. Genes implicated through SMR analysis that pass $P_{GWAS}$ , $P_{eQTL}$ , $P_{SMR}$ and $P_{HEIDI}$ tests are designated by a dagger (†).

We order these from Signal 1 (S1) through 8 (S8) according to their base pair position. Throughout the text, variants involved in a signal referenced with their signal number, or by variant ID alone when representing WBC-specific associations unrelated to asthma. When referencing both signal number and variant ID (e.g., S4—rs4795399), this identifies the lead SNP or top posterior probability SNP from a credible set by ancestry group, analytical method, and asthma definition, while other relevant proxies are referenced in the **Supplementary Tables**. This nomenclature enables rapid distinction of asthma-relevant signals versus WBC-only associations and facilitates comparison of candidate causal variants and accumulation of causal evidence per signal.

The eight signals showed distinct patterns across age-of-onset strata. S4—rs4795399 represented the most statistically significant association overall (P = 2.9×10^−132^ for childhood asthma; P = 7.7×10^−87^ for early asthma), corresponding to the hallmark association first reported at this locus [5]. Five signals were unique to single strata: Signals 1 and 3 emerged only in early asthma, Signal 2 was specific to adult asthma, Signal 5 to childhood asthma, and Signal 6 to late asthma (**Table S5**). Signal 7 appeared in childhood, adult and early asthma, while Signal 8 was detected in adult and early asthma with stronger effects in childhood cases (**Table S6**).

Six of the eight signals overlapped with previously reported asthma associations, validating our stratified analysis approach. The remaining two signals (S2—rs72832915 and S6—rs507671) were novel for asthma but have been reported for other phenotypes (**Table S7**). Analysis of non-European UKB and BBJ cohorts did not identify additional signals at this locus, establishing these eight signals as the reference maximal landscape of statistically independent associations (**Table S8**).

### 2.3 Multi-ancestry asthma meta-analyses

To evaluate the robustness of the eight signals, we conducted multi-ancestry meta-analyses. To combine asthma in UKB-EUR with the four non-EUR ancestry groups, asthma was re-analyzed as a non-stratified phenotype in EUR (denoted EUR-asthma*_all_*). Two meta-analyses were performed: (i) across all populations (META), and (ii) across non-European subsets AFR, SAS, EAS, and BBJ (non-EUR-META), to remove the influence of the high-powered EUR subset.

We first compared conditional analysis on EUR-asthma*_all_* summary statistics with age-of-onset-stratified results. While stratified analyses on EUR individuals identified eight signals, un-stratified analysis identified only four. These were also confirmed in the full multi-ancestry meta-analysis. This included S4—rs12949100 (P = 1.7×10^−18^ in EUR, 2.0×10^−24^ in META; **Figure 2e**; **Tables S8, S9**) and Signals 3, 7, and 8, all via alternate lead SNPs (EUR *r*^2^ ≥ 0.6). In the non-EUR-META analysis, only S4—rs12946510 reached genome-wide significance (P = 1.9×10^−8^; **Figure 2f**; **Figure S6a**), with a consistent direction of effect across all five study populations (**Tables S8, S10**). These findings highlight the robustness of Signal 4 across ancestries. They also underscore the reduced power of the non-EUR subsets to detect additional signals, compounded by the lack of capability for age-of-onset stratification. Consequently, we leveraged WBC traits as immune endophenotypes to enhance signal detection in diverse ancestries. From this foundation of one robustly cross-ancestry validated signal, with some support for three others from the cross-ancestry approach, and strong evidence for all eight signals from the high-powered EUR data, we next investigated WBC trait associations at this locus.

### 2.4 Evidence for WBC subset association within 17q12-q21 compared to the whole genome

Given that asthma and WBC traits together account for more than half of all genome-wide significant associations previously reported at 17q12-q21 (**Table S11**) [26], we tested whether this locus shows unusual enrichment for WBC associations. Using BCX consortium [17] summary statistics from the largest published meta-analysis to date, we compared WBC signal density (accounting for linkage disequilibrium) at 17q12-q21 against 10,000 randomly sampled genome-wide regions of matched size. The 17q12-q21 locus was significantly enriched for WBC associations across ancestries for all major WBC subtypes in EUR (empirical P = 0.0002–0.05), NEU# and WBC# in EAS (empirical P ≤ 0.001), and EOS# and WBC# in AFR (empirical P ≤ 0.02) (**Supplementary Materials: Table S3**). To contextualize these findings, we applied the same enrichment framework to the 5q31-q33 locus, an extended immune-related region spanning 52.3 Mb that includes the IL-4 / IL-13 cytokine gene cluster [27]. This comparison revealed comparable signal densities overall. These results confirm that 17q12-q21 harbors unusually dense WBC trait associations across ancestries. Having established this enrichment, we next systematically investigated WBC traits as potential endophenotypes for asthma across all ancestry groups.

### 2.5 White blood cell trait associations

We identified independent signals by applying WBC trait association scans followed by conditional analysis across 11 WBC traits in all UKB ancestry groups and 6 traits in BBJ. Considering independent signals for each trait, across all traits, the approach yielded 36 independent signals in UKB-EUR and 5 in BBJ, (**Table S12**; see **Methods** for ancestry-specific significance thresholds). We then evaluated whether these WBC signals matched any of the eight asthma signals.

The top WBC association signal within 17q12-q21 was for NEU# at rs2227322 in UKB-EUR (P = 2.8×10^−574^) (**Figure S7**; **Table S12**), and independent of the eight asthma signals (maximum linkage disequilibrium *r*^2^ = 0.15 with S4—rs4795399). The same SNP was also the top NEU# signal in BBJ (P = 1.65×10^−49^), and genome-wide significant in UKB-AFR and UKB-SAS, although neither it nor a proxy marker constituted a distinct signal.

**Table 1** summarizes the overlap between WBC and asthma signals. Five of the eight asthma signals matched WBC trait associations in UKB-EUR. Signals 1, 2 and 6 did not, and were greyed out in **Figure 3** to indicate these signals only displayed accumulated evidence from asthma conditional association testing. Signals 3, 5, and 7 shared the exact lead SNP between asthma and WBC association testing, while Signals 4 and 8 matched through high-linkage disequilibrium proxies (**Table S12**). In BBJ, a match was observed for Signal 4 in WBC# and LYM#. At all eight asthma SNPs, WBC traits demonstrated stronger statistical significance than asthma (**Figure 2g**). For some signals, such as Signal 4 (WBC#) and Signal 8 (EOS and LYM), the lead independent SNP from conditional association matched the lead asthma SNP directly or through a proxy. For others, like Signal 2, no such match was observed (**Figure 2g**). In several instances, the same variant was associated in opposite directions with counts and percentages of a given cell type, compatible with the compositional nature of percentage traits, which are constrained to sum to 100%. As seen in **Figure 2g**, an opposing direction of association does not occur for instances where the SNP is an independent signal for both count and percentage traits of the same WBC subtype.

**Figure 3.**
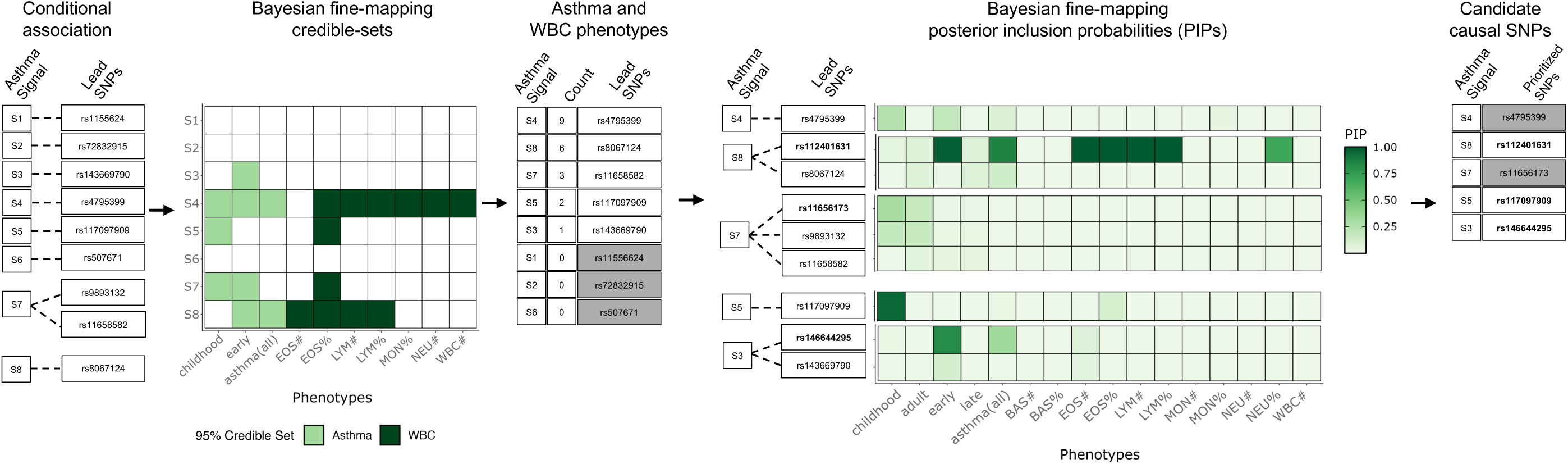
Conditional association and fine-mapping evidence of asthma signals across the 17q12-q21 region and likely causal SNP selection. Conditional association testing: The 17q12-q21 locus contains eight independent asthma signals (S1-S8) identified using GCTA-COJO in the UK Biobank European ancestry subset (UKB-EUR). For each signal, the top conditional association SNP by –log_10_(P) is shown across asthma age-of-onset strata. For Signal 7, two lead SNPs across the age-of-onset strata were identified. Bayesian fine-mapping: Association test statistics across the locus were evaluated through Bayesian fine-mapping using SuSiE across asthma strata and white blood cell (WBC) traits. Signal-phenotype pairs with credible sets are displayed (grid with light green for asthma phenotypes and dark green for WBC traits). Signals were prioritized based on the count of phenotypes with 95% credible sets (CS95). For signals S1, S2, and S6 (greyed out), no CS95s were identified across any phenotype. Within each signal, SNPs were prioritized according to posterior inclusion probability (PIP). For S4 and S7 (greyed out), we could not resolve a single likely causal candidate variant: for S4, multiple SNPs with equal PIPs arising from high pairwise LD; for S7, a higher PIP SNP than the conditional association lead SNPs was identified, but this was modest at 0.3 and similar in magnitude to conditional association lead SNPs.

Additionally, despite the promise of WBC traits as quantitative endophenotypes and the detection of several WBC trait associations, we found no additional matches between WBC and asthma signals in the non-EUR ancestries beyond the single BBJ Signal 4 match. This suggested that our WBC strategy had not yet overcome the power limitations for detecting shared genetic architecture across diverse ancestries. Nevertheless, these findings established a foundation for Bayesian analytical approaches that could reveal evidence for shared causal mechanisms.

### 2.6 Cross-trait ancestry-specific meta-analyses

To further explore ancestry-specific genetic architecture focused on the eight signals for the study populations with lowest sample sizes, UKB-AFR, UKB-SAS, and UKB-EAS, we performed ancestry-specific meta-analyses of asthma and all WBC traits using METAL [28], incorporating a sample overlap correction to adjust for individuals shared across traits within each ancestry. This was followed by conditional analysis using a suggestive significance threshold of *α* = 1.0×10^−4^ [19]. This yielded 13 independent signals (UKB-AFR: 6; UKB-SAS: 6; UKB-EAS: 1) (**Table S13**). Most were distinct from the eight asthma signals, except for a match in UKB-AFR: S4—rs12453507 (P = 1.0×10^−4^), a high-linkage disequilibrium proxy with S4—rs4795399 in EUR (*r*^2^ = 0.84) (but not in AFR, *r*^2^ = 0.19). The genomic segment surrounding this variant was narrowed more than fivefold in UKB-AFR (380.27–380.69 kb) compared to EUR (379.03–381.2 kb) at *r*^2^ *>*0.5. This illustrates the improved signal resolution achievable in African-descent populations. However, this region did not match that identified in infants with asthma of African descent using a combined genetics and transcriptomics approach [10]. This discrepancy makes the finding difficult to interpret, as it may be driven by WBC traits rather than asthma risk. While these ancestry-specific meta-analyses provided some additional signals, they did not substantially advance our understanding of shared causal mechanisms for asthma. We next applied Bayesian fine-mapping to assess strength of evidence across traits at the eight signals and identify likely causal variants within each signal.

### 2.7 Bayesian approaches: fine-mapping across asthma and WBC phenotypes

We performed Bayesian fine-mapping using SuSiE [29] across all ancestry groups, asthma and WBC phenotypes to identify 95% credible sets (CS95) and prioritize causal variants within each signal. In UKB-EUR, fine-mapping identified signals matching five of the eight reference signals (Signals 3, 4, 5, 7, and 8) across asthma phenotypes, with four of these five signals also showing evidence in WBC traits (**Figure 3**,**Figure 4**; **Table 1**; **Figures S8-S9**; **Table S14**).

**Figure 4.**
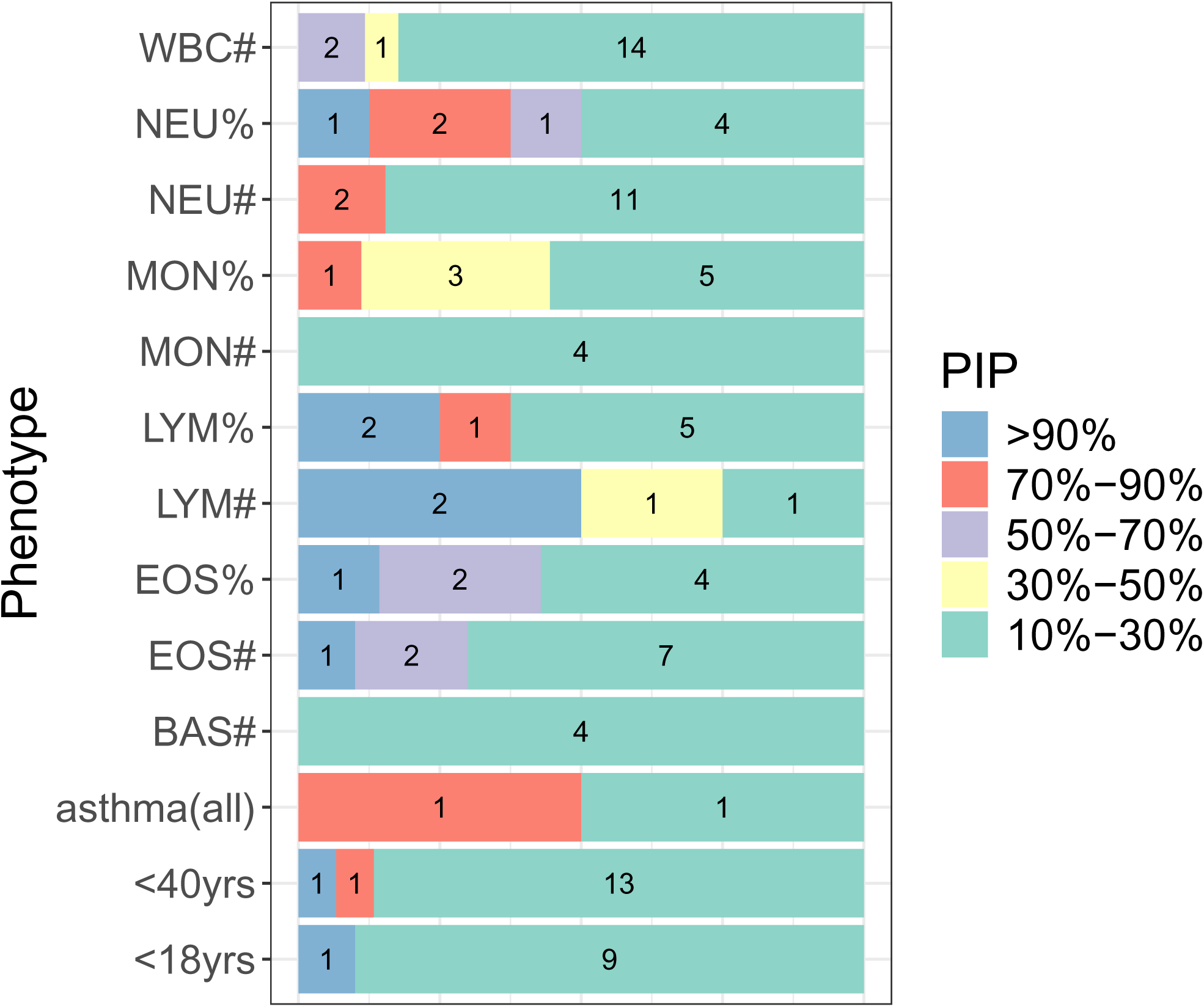
Summary of statistical fine-mapping results for asthma phenotypes and white blood cell traits using SuSiE. Distribution of fine-mapping posterior inclusion probabilities (PIPs) in UKB-EUR. For each phenotype, the plot shows the proportion of variants from all 95% credible sets (CS95s) falling within specified PIP ranges (see legend; variants with PIP *<*10% excluded). No variants from BAS% CS95s met the PIP ≥ 10% threshold. See **Figure S8** for the complete set of CS95s.

Among WBC traits, eosinophil count (EOS#) appeared most frequently across signals, with Signals 4 and 8 showing evidence across multiple WBC traits including EOS#, LYM# and NEU#. Signal 5 and 7 showed evidence only for EOS%. In non-EUR populations, no WBC fine-mapping signals overlapped with the eight reference signals.

Fine-mapping prioritized different candidate SNPs than those identified by conditional association for three signals: Signals 3, 7, and 8. Signal 4 could not be resolved to a single causal variant due to multiple SNPs with equal posterior inclusion probabilities from high linkage disequilibrium. For Signal 5, finemapping confirmed the conditional association lead SNP. Signal 7 demonstrated complex variant resolution: conditional association identified two different lead SNPs across three age-of-onset strata (S7—rs9893132, S7—rs11658582), while fine-mapping prioritized a third variant (S7—rs11656173) with modest PIP (0.30), preventing clear resolution of the causal candidate (**Figure 3**; **Figure S10b**). For Signal 8, fine-mapping identified a different SNP than the original asthma conditional association lead SNP, which exhibited higher PIPs for asthma and even stronger PIPs across multiple WBC traits. For Signal 3, fine-mapping prioritized rs146644295, which had higher PIPs for asthma phenotypes than the conditional association SNP, and corresponded to the lead variant in the cross-ancestry meta-analysis.

Signals 1, 2, and 6 showed no fine-mapping support, consistent with their limited causal evidence—these were specific to single asthma strata and lacked WBC co-associations. FINEMAP [30] corroborated all SuSiE results (**Figure S11**; **Table S15**). To validate these fine-mapping discoveries, we next assessed whether our WBC trait associations were replicated in independent data.

### 2.8 Replication of Bayesian fine-mapping WBC signals

We next assessed whether the WBC trait associations identified by fine-mapping were also observed in the BCX consortium meta-analysis [17](Section 2.4). A subset of our findings were confirmed by BCX across the eight signals: Signal 8 was identified in BCX in EUR for EOS only, and Signal 4 by LYM in both EUR and EAS. In AFR, which benefitted from a large sample size, Signal 4 was supported by EOS while our AFR analysis missed this signal. No other signals showed replication in BCX based on a fine-mapping posterior inclusion probability (PIP) threshold of 0.1 (**Tables S16-S17**). BCX used fine-mapping method MR-MEGA [31], which assumes a single causal variant. In contrast, our fine-mapping analyses allowed multiple causal variants. These findings underscore the limitations of single-variant fine-mapping. They highlight the added value of our multi-trait, multi-population approach for detecting multiple relevant signals at one locus. Beyond identifying variants with causal evidence for both asthma and WBC phenotypes, we sought to determine whether the same causal variant underlies both traits, potentially placing WBC traits in the causal pathway to asthma.

### 2.9 Bayesian approaches: Colocalization

Colocalization was assessed under three complementary sets of assumptions: (i) a pair of traits with a single causal variant using COLOC [32] (**Figure 5a,b**; **Tables S18-S19**), (ii) a pair of traits allowing for multiple causal variants using COLOC-SuSiE (**Figure 5a,b**; **Tables S20-S21**), and (iii) multiple traits with a shared single causal variant using HyPrColoc [33], (**Figure 5c**; **Figures S12-S14**; **Table S22**). All colocalizations detected across pairs from among all asthma and WBC phenotypes and study populations are summarized in **Figure S12**.

**Figure 5.**
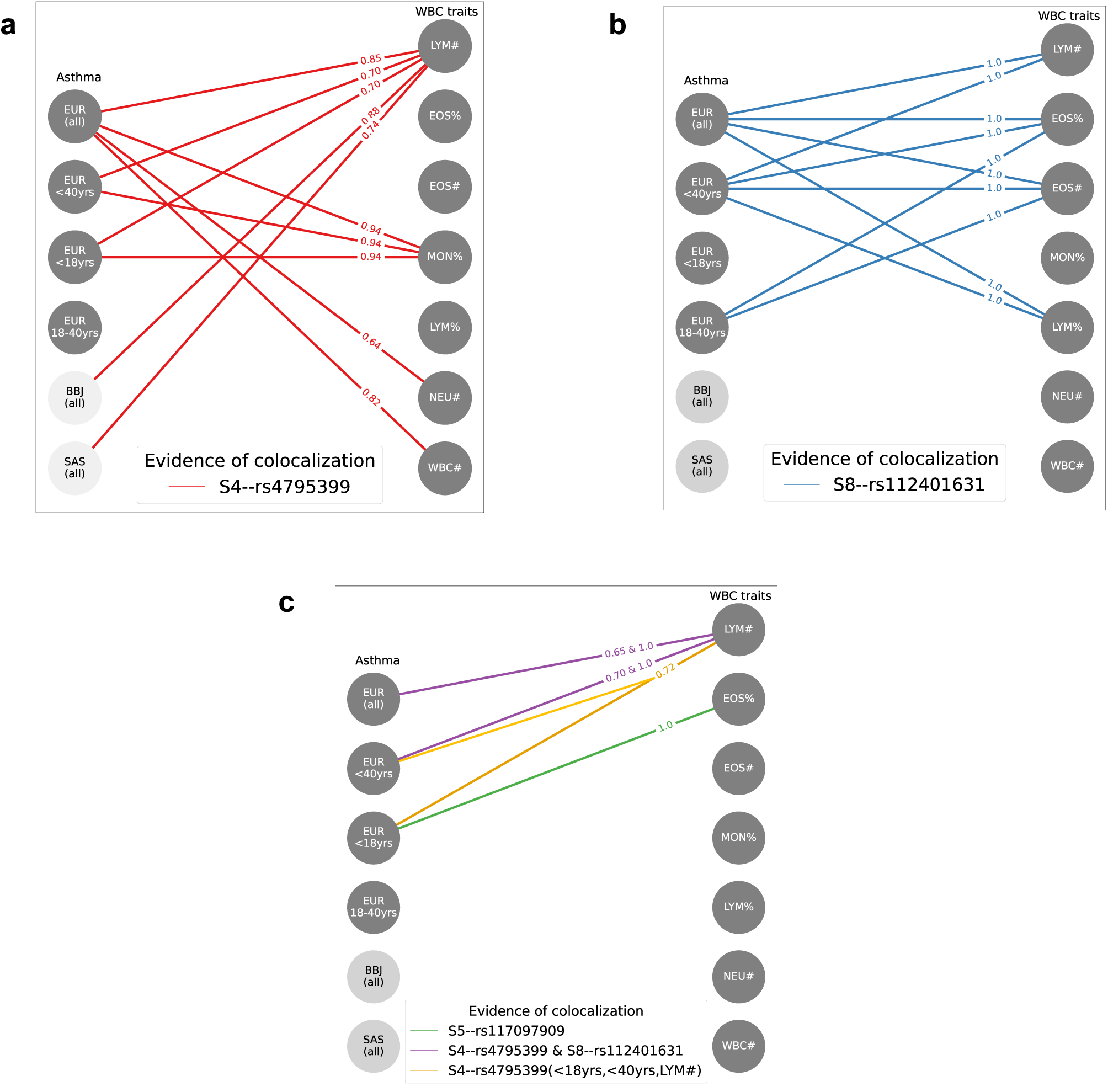
Summary of colocalization analysis between asthma signals and white blood cell traits. Asthma signals with evidence of single and multiple causal variant colocalization between asthma and WBC traits (COLOC-SuSiE H4 PP *>*0.6). Panels (a-c) (left): asthma phenotype nodes, with dark grey for UKB-EUR and light grey for non-EUR populations (BBJ and SAS with black text); (right): WBC trait nodes. Line edges represent evidence of colocalization between the asthma and WBC trait-pair within the study population indicated in the left node (inline-text shows the H4 PP value). Panels (a) and (b) depict single causal variant colocalization, while panel (c) shows multiple causal variant colocalization. In panel (c), the purple line represents multiple causal variant colocalization, with H4 PP values for S4 and S8 shown sequentially. The orange line indicates multi-trait (*>*2 traits) colocalization (from HyPrColoc). For test statistics, see **Tables S18-S22**.

Considering pairs of traits including one asthma and one WBC phenotype, asthma and LYM shared Signal 4 in three study populations: UKB-EUR, UKB-SAS, and BBJ (**Figure 5a**, **Figure 6**, **Figure S13**). Nine variants appeared across credible sets in the three populations. Additional pairs colocalizing at Signal 4 in UKB-EUR included asthma with MON#, NEU#, or WBC# (**Figure S12b-f**). In UKB-EUR only, asthma shared Signal 8 with increased EOS# or decreased LYM# levels, with posterior probabilities at S8—rs112401631 above 0.99 (**Figure 7a-c**).

**Figure 6.**
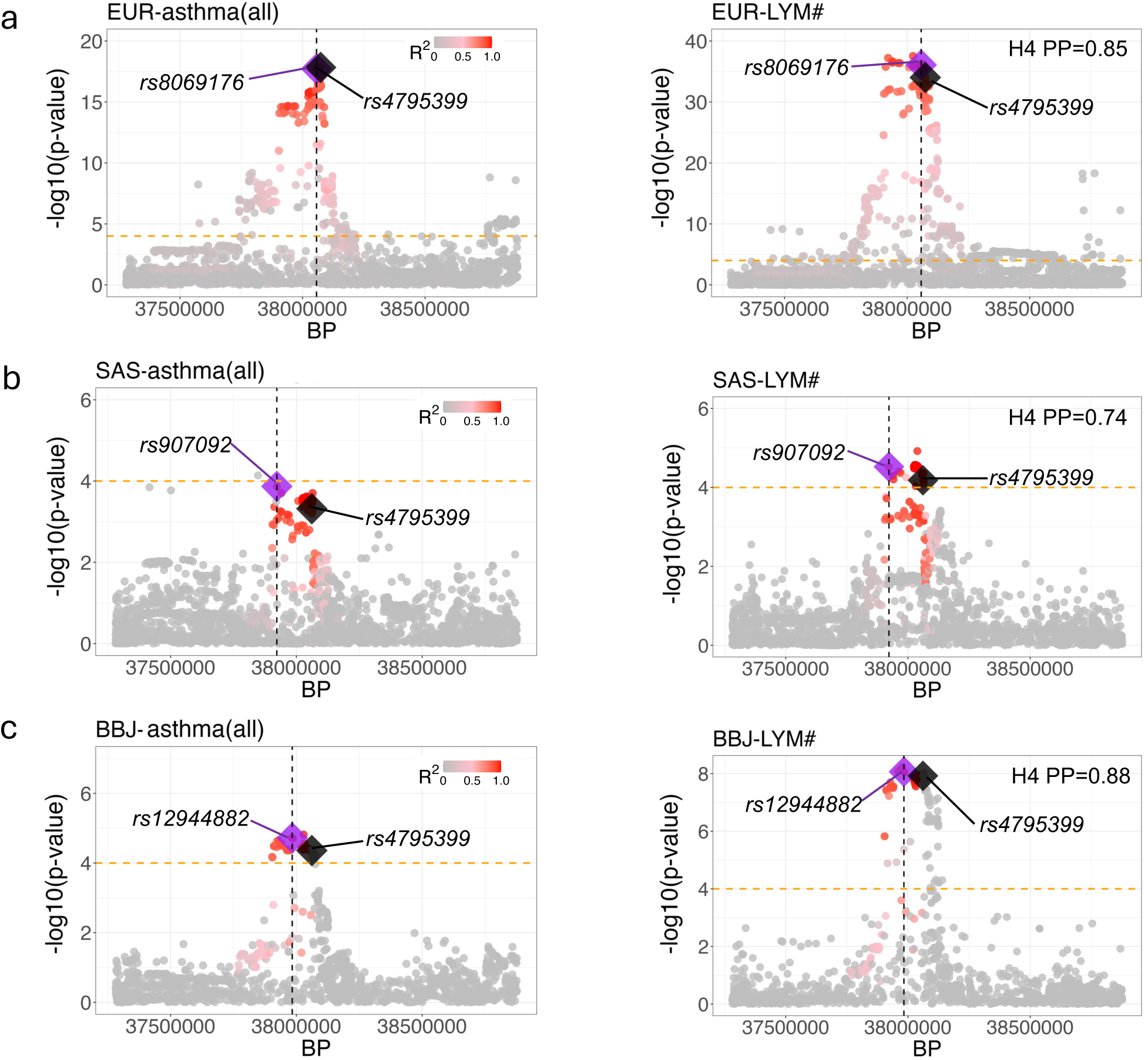
Shared causal variant between asthma and lymphocyte count (LYM#) across three study populations for signal S4. (a–c) Colocalization of S4 with LYM# for asthma*_all_* in UKB-EUR (a), UKB-SAS (b), and BBJ (c). The lead variant from the 95% credible set, representing the highest posterior probability of colocalization, is shown as a purple diamond, with its genomic position marked by a vertical dashed line. Nearby variants are shaded by their linkage disequilibrium (LD, *r*^2^) with the lead SNP (see key). The most significant GCTA-COJO asthma SNP, rs4795399 (S4), is shown as a black diamond to indicate its position and pairwise LD with the colocalization lead SNP. The orange dashed horizontal line denotes the suggestive significance threshold at P = 1.0×10^−4^.

**Figure 7.**
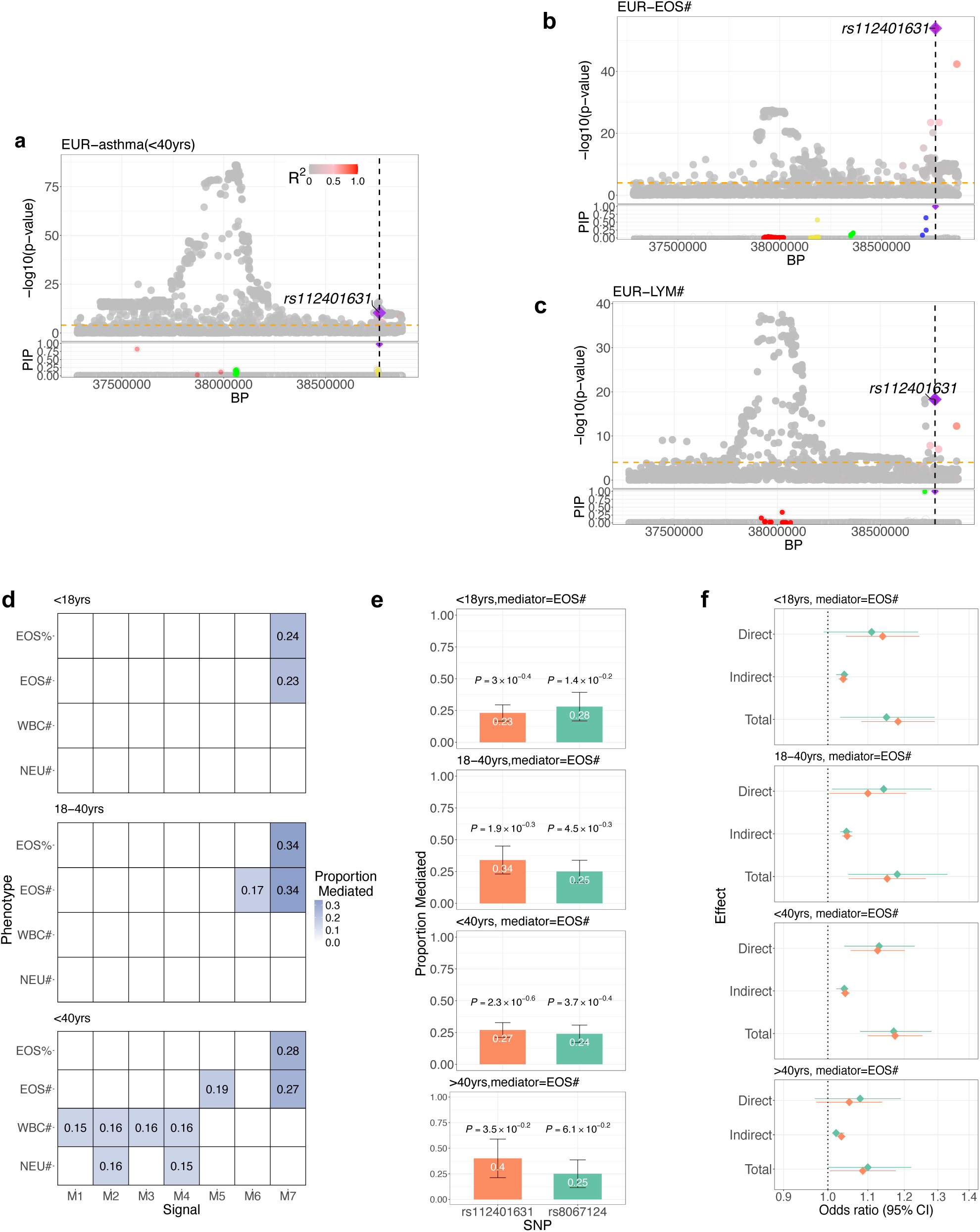
Summary of colocalization and mediation analysis for signal S8. (a-b) Colocalization of S8—rs112401631 with asthma and white blood cell (WBC) traits. Regional plots show colocalization evidence for rs112401631 in asthma with age-of-onset *<*40 years (AO-*<*40yrs) and two WBC traits: eosinophil count (b) and lymphocyte count (c), based on COLOC-SuSiE. (d) Mediation of asthma phenotypes by WBC traits in UKB-EUR. The proportion of the genetic effect of each marker on asthma mediated through individual WBC traits is indicated by the intensity of blue. Variants showing evidence of mediation were filtered and grouped into seven signals (M1-M7) using pairwise LD clumping (*r*^2^ *>*0.5 within 1000 kb). Signal M7 includes two SNPs: S8—rs112401631 and S8—rs8067124 (both part of S8; see **Tables S20,S21**). (e) Mediation of asthma phenotypes by eosinophil count for rs112401631 and rs8067124 (M7). Bar plots show the proportion mediated (white text) across four age-of-onset strata. P-values above each bar correspond to the proportion mediated for each SNP (see **Tables S23,S24**). (f) Odds ratios for the direct, indirect, and total effects of eosinophil count on S8—rs112401631 and S8—rs8067124 across the four asthma strata.

Considering multiple traits and one causal variant, asthma, EOS#, and LYM# were implicated at Signal 8 (**Figure 7a-c**; **Tables S18, S22**). Considering multiple causal variants for trait pairs in UKB-EUR, asthma and LYM# jointly colocalized at Signals 4 and 8 (**Figure 5c**; **Figure S12p–q**). Additionally, asthma colocalized at Signal 5 with EOS% only (**Figure 5c**; **Figure S12g**). The candidate causal SNPs identified across colocalization methods for Signals 5 and 8 matched those from fine-mapping. Linkage disequilibrium patterns across lead SNPs for each signal from conditional association, meta-analyses, fine-mapping, and colocalization analyses are displayed in **Figure S10**. Again, Signal 4 could not be resolved among EUR populations due to extensive linkage disequilibrium.

Crucially, colocalization analysis unlocked the potential of non-EUR ancestry groups where previous methods had not provided much supporting evidence for the eight signals. While association testing and fine-mapping yielded limited signals in UKB-SAS and BBJ, colocalization revealed robust shared causal variants at Signal 4 with posterior probabilities of 0.74-0.88 (**Table S19**). This demonstrated that Bayesian methods could reveal evidence for shared causal pathways across ancestries where classical approaches had failed. Having established these shared causal variants between asthma and WBC traits, we next quantified the extent to which WBC traits might mediate genetic effects on asthma risk.

### 2.10 Mediation through WBC traits

We conducted mediation analyses [34] in UKB-EUR age-of-onset-stratified asthma across all 17q12–21 region SNPs using WBC traits as putative mediators. Among 25 SNPs showing evidence of *>*15% mediation (P ≤ 1.0×10^−4^), the strongest evidence was observed for S8—rs112401631 and S8—rs8067124, with EOS# mediating 23–40% of the asthma effect across age strata (**Figure 7d-f**, **Tables S23-S24**). The highest mediation proportion (40%) was observed for S8—rs112401631 in the late asthma stratum. These variants, which represent both the conditional association candidate and fine-mapping and colocalization lead variants at Signal 8, formed a two-SNP linkage disequilibrium cluster (M7) among seven total mediation clusters identified. No other mediation clusters corresponded to the eight reference signals, and no significant mediation was detected in non-EUR populations.

### 2.11 Gene expression using Summary data-based Mendelian Randomization (SMR) and regulatory elements data from external functional databases

We performed SMR analysis [35] in UKB-EUR across 16 asthma and WBC phenotypes using eQTL evidence from GTEx V8 (49 tissues) [36] and CAGE (peripheral blood) [37] to identify genes with evidence that their expression levels mediate the eight signals. Signal 4 demonstrated the strongest evidence, with multiple genes (*ORMDL3*, *GSDMB*, *IKZF3*, and *ZPBP2*) passing both SMR significance and HEIDI tests across different tissues, indicating shared genetic regulation of gene expression and trait variation (**Figure 8**; **Table 2**; **Figures S15-S17**; **Tables S25-S28**). *ORMDL3* and *IKZF3* showed significant associations in whole blood, *ORMDL3* and *GSDMB* in lung tissue, and *GSDMB* in peripheral blood from CAGE data. Notably, *ZPBP2* in testis was the only gene-tissue combination significant for both asthma phenotypes and a WBC trait (LYM#). Additionally, the CD40-associated lncRNA (*RP11-387H17.4*) at Signal 4 had not been previously linked to asthma. Many different Signal 4 SNPs were identified as the likely causal SNP in this analysis.

**Figure 8.**
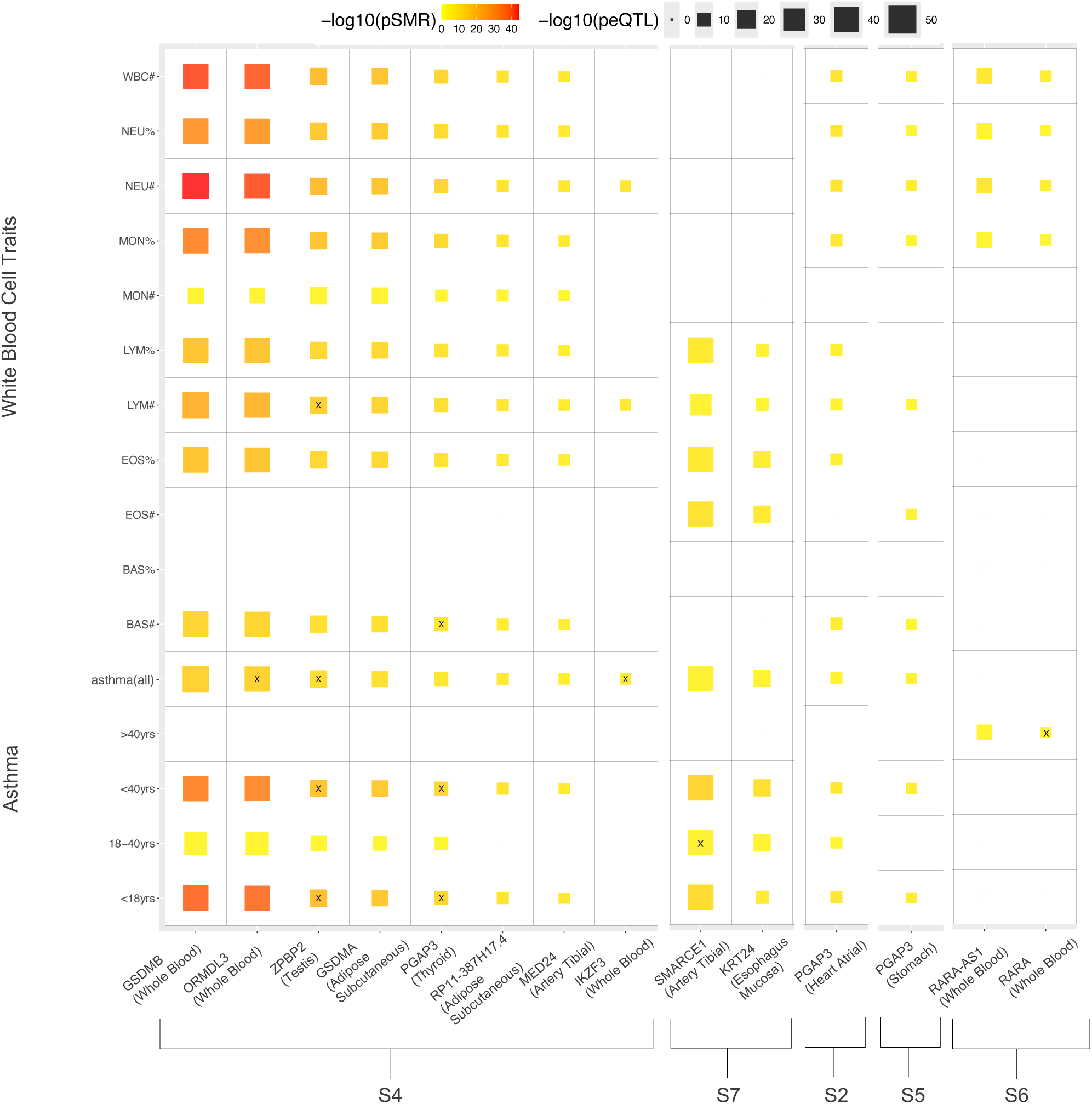
Signal-gene pairs with significant Summary-data-based Mendelian Randomization (SMR) associations across tissues and traits. For each signal-gene pair, tissues with the most significant SMR associations (P *<*0.05/m, where m is the number of probes tested in the study region per tissue; see **Table S23** for tissue-specific correction) are shown, along with SMR results for each phenotype. Coloured squares denote significant associations, with black “X” marks indicating gene–trait pairs that also pass the HEIDI test (P *>*0.05), suggesting shared genetic regulation of both gene expression and trait variation. Specifically, S4—*ORMDL3* and S4—*IKZF3* (asthma*_all_*, Whole Blood), S4—PGAP3 (*<*18yrs, *<*40yrs, BAS#, Thyroid), and S4—*ZPBP2* (asthma*_all_*, *<*18yrs, *<*40yrs, LYM#, Testis) all pass the HEIDI test in the indicated tissues. Among these, only S4—*ORMDL3* and S4—*IKZF3* show significance in Whole Blood. For a full list of signal-gene pairs passing both SMR and HEIDI tests across all tissues, see **Table 2**

**Table 2:**
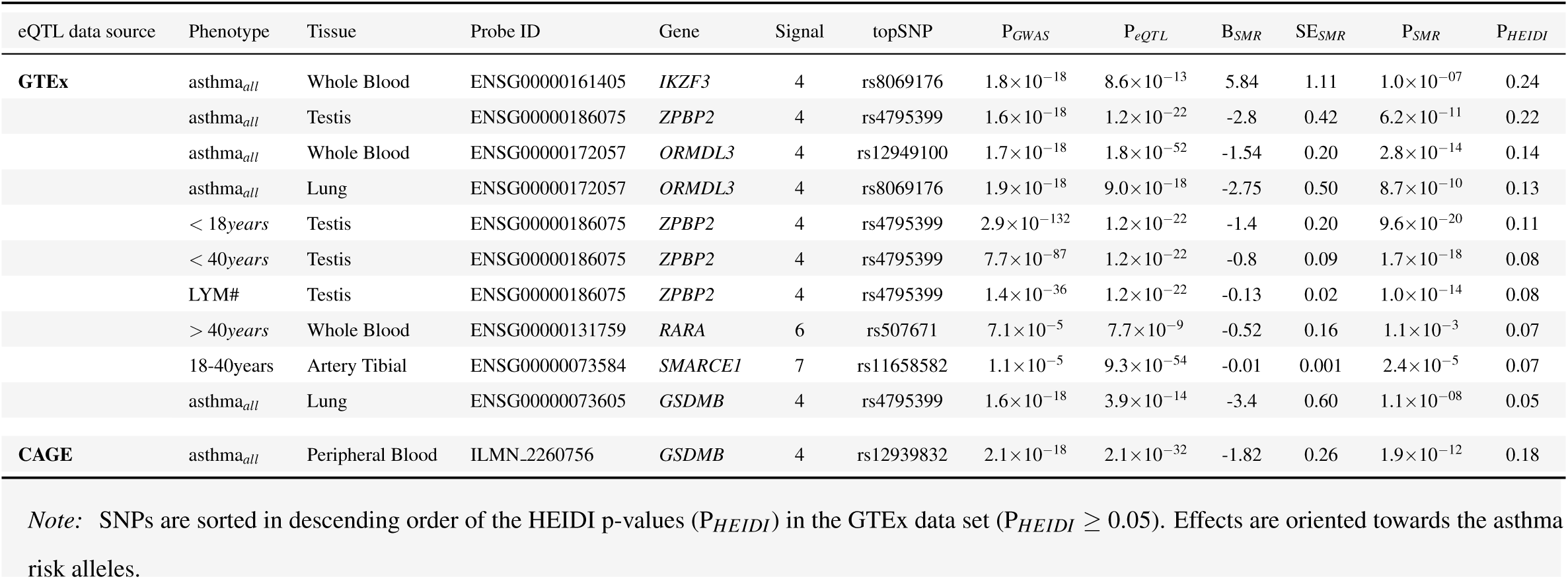
Genes showing significant pleiotropic associations between expression levels and asthma phenotypes or white blood cell traits in the UKB-EUR.

For Signals 2, 5, and 6—which lacked prior WBC co-association, fine-mapping, or colocalization support and were not reported for asthma in the literature (Signals 2 and 6)—SMR provided the first additional causal evidence by identifying convergent gene-tissue associations for the same candidate SNPs identified by conditional association across asthma and WBC traits (**Tables S26-S28**). Signal 6 implicated *RARA* and *RARA-AS1* not previously linked to asthma.

RegulomeDB [38], analysis revealed that Signals 3, 4, and 8 showed the greatest likelihood of gene regulatory involvement supported by ChIP-seq data, with Signals 3 and 8 implicating the most likely causal candidates from fine-mapping and colocalization analyses (**Table S29**).

## 3 Discussion

This study demonstrates that integrating multiple complementary analytical strategies can extract more biologically meaningful causal information than conventional single-approach analyses: (1) age-of-onset heterogeneity through stratified analysis, (2) white blood cell traits as quantitative endophenotypes, (3) inclusion of non-European multi-ancestry groups for enhanced signal resolution and independent sources of signal support, and (4) statistical methods including Bayesian fine-mapping and colocalization, and mediation analysis (5) incorporation of external sources of gene expression data through stringent Mendelian Randomization methodology. Relying on any single strategy would have provided an incomplete picture. Only 1–4 asthma signals were detected in previous UKB-EUR genome-wide studies [39, 40].

The multi-pronged approach yielded three advances: enhanced signal detection through age-of-onset heterogeneity [41, 42], improved mechanistic understanding through WBC trait colocalization, and cross-population validation despite limited power in individual non-European cohorts. Most importantly, we demonstrate that systematic evidence accumulation across multiple analytical categories provides a robust framework for prioritizing variants and genes for functional validation—a critical need given the resource constraints of experimental follow-up studies. While a prior study conducted genome-wide colocalization analyses between asthma and WBC traits in European ancestry individuals [43]—and identified WBC colocalization at our top WBC signal rs2227322—none of their WBC signals showed colocalization with asthma. Our study is the first to report asthma signal-specific colocalization with WBC subtypes using age-of-onset stratification and multi-ancestry data.

Signal 8 emerges as the most mechanistically resolved, with candidate variant rs112401631 acting as a trans-eQTL for *IKZF2* on chromosome 2, and showing convergent evidence across fine-mapping, eosinophil and lymphocyte colocalization, mediation analysis, and functional annotation. The variant lies within a chromatin binding site linked to differential *IKZF2* expression [38], a transcription factor essential for regulatory T-cell development and Th2 cytokine modulation [44]. This suggests a compelling causal pathway: reduced *IKZF2* expression may alter T-cell responsiveness, reducing restraint of eosinophil-promoting responses and increasing asthma susceptibility.

Signal 4 illustrates the complexity of this locus, with SMR analyses implicating *ORMDL3*, *GSDMB*, and *ZPBP2* expression in blood tissues. *ORMDL3* and *GSDMB* align with established functional evidence showing that asthma-risk variants disrupt CTCF-binding and alter chromatin architecture in airway epithelial cells [45]. *ZPBP2*, however, represents a potentially underexplored mechanism. It was the only gene showing coherence between multi-ancestry lymphocyte colocalization and SMR analyses, for which both asthma and lymphocyte count were identified, and functional studies have demonstrated its protective role against airway hyperresponsiveness through reduced CD4+ T-cell infiltration [45]. Signal 4 asthma risk variants are known to downregulate *ZPBP2* expression as part of a broader chromatin remodeling cascade [46]. Additional support for lymphocyte function at this signal comes from CRISPR/Cas9-mediated deletion of a Signal 4 proxy, which reduced *ORMDL3* and *IKZF3* expression in T-helper cells [47]. The colocalization with decreased lymphocyte counts suggests a plausible epithelial-to-immune signaling cascade in which increased *ORMDL3* expression suppresses IL-2 production and impairs CD4+ T-cell proliferation [10, 45]. The CD40-associated lncRNA (*RP11-387H17.4*) was also identified for Signal 4; although not previously linked to asthma through genetic association, its expression has been associated with asthma in transcriptomic studies involving environmental exposures [48]. Together, this evidence supports a model in which Signal 4 asthma-risk variants orchestrate pleiotropic regulatory effects on several genes, influencing epithelial pathways and lymphocyte function [5].

Signal 6 provides the first evidence linking vitamin A metabolism genes to asthma etiology, with the risk allele associated with reduced *RARA* expression in whole blood tissues. This finding aligns with epidemiological data linking vitamin A deficiency to asthma risk [49, 50] supporting biological rationale for supplementation strategies. The restriction to late-onset asthma suggests potential gene–environment interactions contributing to increased penetrance with aging. This supports a mechanistic role for *RARA*-mediated immune regulation, including T-cell differentiation and mucosal immunity, in asthma susceptibility [51]. Signal 2’s candidate variant rs72832915 represents a novel asthma association and was associated with increased PGAP3 expression, implicating GPI-anchor remodeling mechanisms which regulate the surface display of immune and epithelial proteins relevant to airway inflammation [52]. Signal 1’s low-frequency missense variant rs11556624 in STARD3, a cholesterol transfer protein, has been linked to HDL cholesterol [53] and adds to growing evidence for lipid metabolism in asthma pathogenesis [54]. Signal 5’s rs117097909 demonstrated high posterior probability in fine-mapping and eosinophil-specific colocalization, potentially affecting epithelial-eosinophil interactions. Signal 7’s associations with *SMARCE1* and *KRT24* expression suggest roles in chromatin remodeling and epithelial differentiation, respectively [55, 56].

Our WBC endophenotype approach complements rather than conflicts with established tissue-specific findings. While Ober et al. showed that genetic regulation of *GSDMB* expression in airway epithelial cells is central to childhood-onset asthma [10], our colocalization with systemic immune cell counts adds a complementary immunobiological layer. Epithelial-derived cytokines promote eosinophil activation and Th2/ILC2 responses [10, 57, 58] linking airway gene regulation to changes in peripheral immune cell profiles. Similar to tissue-specific eQTL colocalization, our results suggest a tractable pathway connecting epithelial variation to systemic immune effects.

Several limitations constrain our interpretations. This chromosomal region is analytically challenging; for example, the strongest WBC association signal (rs2227322, P = 2.8×10^−574^) shows no asthma association but creates interference patterns through linkage disequilibrium for WBC traits, potentially obscuring weaker true signals. While we successfully identified colocalization signals for eosinophils and lymphocytes that inform pathophysiology, consistent with shared genetic architecture across asthma and WBC traits, the WBC endophenotype strategy did not fully overcome power limitations in non-European populations [59]. The UKB-AFR cohort yielded no genome-wide significant asthma signals and fewer WBC associations than other populations, underscoring persistent challenges in studying diverse populations [60]. This reinforces the need for substantial expansion of African-ancestry genomic datasets [61, 62, 63] to enable meaningful contributions to fine-mapping efforts.

Methodologically, WBC traits were measured at recruitment [20, 22] rather than during active disease, limiting direct inference about acute asthma pathophysiology. However, this timing supports causal modeling by capturing baseline immune phenotypes rather than disease consequences. The time lapse between asthma diagnosis and recruitment may affect case classification, though similar age-at-recruitment profiles across strata suggest non-differential bias. Our WBC-focused approach may have missed causal pathways involving other cell types, particularly airway epithelial cells, which are increasingly recognized as central in asthma pathogenesis[10, 57, 58]. Integration of eQTL data from healthy individuals [36] was partially consistent with fine-mapping but also implicated non-relevant tissues, highlighting the need for disease-contextualized functional data. Furthermore, the absence of gene identification for some signals through SMR is not surprising, as disease-associated variants are often located in regulatory regions of genes under purifying selection and with complex regulatory architectures, limiting their detectability in conventional eQTL studies [64]. Finally, while we provide strong evidence for specific causal variants and pathways, functional validation will be essential to confirm mechanisms and therapeutic potential.

Therapeutic implications vary across signals. Signal 8 is the most tractable target given its tight causal resolution and immune-specific mechanism via *IKZF2*. Signal 4, though complex due to high LD and multiple genes, showed consistent lymphocyte associations across populations. Among Signal 4 genes, *ZPBP2* presents an intriguing candidate through SMR support for lymphocyte levels, though its highest expression in testis rather than asthma-relevant tissues may limit its therapeutic relevance. Signal 5’s eosinophil-specific colocalization complements existing anti-IL-5 strategies used in eosinophilic asthma [65], while Signal 6’s vitamin A metabolism findings suggest new supplementation-based interventions. Given the roles of lymphocytes and eosinophils in orchestrating and executing asthma inflammation [14], Signals 4 and 8 highlight promising anti-inflammatory targets upstream of disease initiation. This fine-mapping and endophenotype colocalization framework extends beyond asthma to other immune-related diseases with quantitative traits, such as cardiovascular disease or autoimmune conditions. The demonstration that systematic, multi-strategy analysis can reveal novel causal information provides a generalizable template for dissecting complex traits. Genetic support for drug targets, especially from multiple causal lines of evidence, markedly increases the probability of successful drug development [66].

Immediate priorities include functional validation of high-priority variants, particularly rs112401631 (*IKZF2*) and the *ZPBP2* pathway, via CRISPR-based studies in relevant cell types. Long-term goals include translating these insights into therapeutic interventions, particularly targeting vitamin A metabolism and immune tolerance pathways. The evidence accumulation framework presented here—integrating fine-mapping, colocalization, mediation analysis, and external validation—provides a practical roadmap for identifying and prioritizing candidate variants and genes for mechanistic studies. As genomic datasets continue to expand, such systematic workflows will be critical for efficiently translating genetic associations into experimentally testable hypotheses.

## 4 Conclusion

This study demonstrates that integrating age-of-onset stratification to address etiological heterogeneity, white blood cell quantitative traits as putative endophenotypes, and multi-ancestry analyses along with deploying Bayesian fine-mapping and colocalization algorithms can substantially advance understanding of complex loci such as 17q12-q21. In asthma, this approach proved particularly informative and offers a generalizable strategy for dissecting diseases with diverse clinical subtypes and components that may involve different sets of relevant endophenotypes. The evidence supporting Signal 4 exemplifies how cross-ancestry convergence and integrative functional genomics can establish specific features of causal pathways involving immune mechanisms. In other instances, as exemplified by Signal 8, combining fine-mapping, colocalization with multiple immune traits, and mediation analysis allowed us to hone in on a specific candidate causal variant. Overall, five signals showed evidence of co-association with white blood cell traits, while among the others, Signal 1 was the most clearly non-immune in nature. This can help classify signals for specific follow-up studies and highlights that the absence of immune involvement can itself be informative, underscoring the mechanistic heterogeneity of asthma. The signals varied in the amount and type of supporting evidence: Signal 8 showed strong evidence of implication of WBC traits without gene expression support, while Signals 2 and 6, although lacking WBC trait co-association and fine-mapping, pointed to novel gene expression pathways plausibly relevant to asthma. Additional validation of new asthma Signals 2 and 6 will be important to clarify their biological and clinical relevance. Notably, our findings showed that combining endophenotype integration with Bayesian fine-mapping made it possible to leverage non-European ancestry groups with much smaller sample sizes; rather than discarding these datasets for reasons of low power, they contributed meaningful evidence that strengthened our results. Together, these insights illustrate how endophenotype-informed, multi-ancestry strategies can move beyond statistical associations toward biologically grounded models of disease risk. This framework provides a scalable approach for dissecting complex diseases with both immune and non-immune components and can be broadly applied to other disease contexts where quantitative endophenotypes inform causal pathway model construction.

## 5 Methods

### 5.1 Study participants

We drew on UK Biobank study subjects [20] with asthma status and WBC measures. For the European (EUR) ancestry subset, we included individuals who self-identified as White British (*N* = 408,963), aligning with prior genetic studies to reduce population stratification. For non-European populations, we assigned ancestry using UMAP-HDBSCAN [67] clustering on genetic principal components (see **Methods** section in the **Supplementary Materials**) resulting in three non-EUR groups from UKB: African (AFR; *N* = 10,065); South-Asian (SAS; *N* = 9,366) and East-Asian (EAS; *N* = 2,549). This clustering approach enabled us to incorporate individuals from admixed or underrepresented groups who are often excluded from analyses based solely on self-reported ethnicity. We also drew on individuals from The Biobank Japan Project (BBJ) with asthma and WBC data (*N* = 161, 280)[22].

### 5.2 Phenotype definition

Asthma cases in the UKB were defined as participants who self-reported a doctor-diagnosed asthma condition in both data field 6152 and data field 20002. Controls were participants who answered “no” to asthma in field 6152 and did not report asthma in field 20002. Age of asthma onset was obtained from field 3786. In BBJ, cases were defined based on physician-diagnosed bronchial asthma, and controls were individuals without a physician-documented asthma diagnosis, following the protocol by Nagai et al.[22]. Full details, including exclusion criteria and quality control, are provided in the **Methods** section in the **Supplementary Materials**. We adopted two approaches to defining asthma as a phenotype for association testing.: (I) asthma adjusted for sex, age at recruitment, genotyping array, and the top 10 principal components in each study population (asthma*_all_*); (II) age-of-onset (AO) stratified analysis in the UKB-EUR, with strata defined by onset age but still adjusted for the same covariates as in (I). The AO strata included: onset *<*18 years (childhood asthma; 13,419 cases); 18–40 years (adult asthma; 12,526); *<*40 years (early-onset asthma; 25,945); and ≥40 years (late-onset asthma;18,052), using 361,244 shared controls (see **Supplementary Materials: Table S2**). The divisions into the three categories match a recent previous study [2] providing clinical relevance to the cut-offs at ages 18 and 40 years based on comorbidities, asthma control and demographic factors and these divisions were also used in a recent asthma genetics study using the UK Biobank [68].

We modelled white blood cell (WBC) traits across UKB populations by transforming residuals to the standard normal distribution following adjustment of raw values for PCs and technical covariates [69] (see **Methods** section in the **Supplementary Materials**) using rank-based inverse normal transformation in R. WBC traits in the BBJ were modelled using methods described by Kanai et al. [70], adjusted for using fewer covariates (age, sex, top 10 genetic PCs) and similarly normalized. Counts and percentages of WBC traits were designated with “#” and “%” respectively.

### 5.3 Study region

We defined the study fine-mapping region on 17q12-q21 using the “SNP2GENE” function from FUMA [25], applied to summary statistics from the four age-of-onset asthma related strata (childhood-, adult-, early– and late-onset) to determine the region boundaries. The region defined using the early-onset stratum (*<*40yrs) was the most inclusive, encompassing all boundaries identified in the other strata 37,281,157-38,876,409bp (hg19; 1.6Mb). This definition was obtained using the parameters: P ≤ 1.0×10^−5^, LD *r*^2^ *<*0.5 and 10,000 unrelated UKB, White-British participants for LD reference.

### 5.4 Simulations using BCX consortium data

To explore whether 17q12-q21 contained an excess of independent signals across the genome, we simulated a null hypothesis distribution for the count of signals by randomly drawing similarly sized regions (1.6 Mb; 10,000 segments) across autosomes using association test summary statistics for the counts of basophils, eosinophils, lymphocytes, monocytes, neutrophils and total white blood cells from participants of European (EUR, *N* = 563,946), African (AFR, *N* = 15,171), East-Asian (EAS, *N* = 151,807) and Hispanic (HIS, *N* = 9,368) ancestry from the BCX consortium [17] using the GenomicRanges R-package. Independent signals were identified using SNPCLIP-based [71] pruning for each WBC trait and population (MAF ≥ 0.02, LD *r*^2^ *<*0.2; EUR: P *<*5.0×10^−8^; non-EUR: P *<*1.0×10^−4^). We excluded the HLA region (chr6:29.7–33.1 Mb, hg19) and the Duffy/DARC region for AFR and HIS (chr1:158.7–159.6 Mb). Empirical p-values were computed based on the observed counts within chr17q12–21 (see **Methods** section in the **Supplementary Materials**). We repeated this analysis for a second well-known asthma-related locus, 5q31–q33 (chr5:122.06–174.37 Mb; 52.3 Mb), chosen for its established role in immune regulation and inclusion of multiple interleukin genes (e.g., IL3, IL4, IL5, IL13).

## 6 Association analysis

We ran quality control (QC) filters on imputed dosages (MAF ≥ 0.01, HWE p-value ≥ 1.0×10^−6^, imputation info score R^2^ ≥ 0.3) for bi-allelic autosomal markers using PLINK [72] (EUR-8,056; AFR-13,730; SAS-8,395; EAS-6,875; BBJ-6,483 variants) and retained 3,381 variants in the EUR; 5,778-AFR; 2,162-EAS; 3,050-SAS and 2,047-BBJ. We conducted association scans for asthma (see **Phenotype definition** section in **Supplementary Materials**): (I) Population-based and meta-analysis scans for asthma*_all_* using SAIGE [73], which accounts for case–control imbalance and models relatedness using a sparse genetic relationship matrix (GRM). SAIGE was applied to each ancestry group independently to ensure consistent modeling across populations. (II) Age-of-onset (AO)-stratified analysis in UKB-EUR using BOLT-LMM [74]; both algorithms model fixed and random effects under a linear mixed model to account for sample relatedness and population structure. We also analysed WBC traits across study populations using BOLT-LMM. LD scores for calibrating BOLT-LMM statistics were computed using LDSC [75] for the non-EUR UKB populations; pre-computed LD scores provided in the package were used for UKB-EUR and BBJ.

### 6.1 Fixed effects multi-ancestry meta-analysis

We aggregated summary data across study populations using fixed-effect inverse variance weighted meta-analysis implemented in METAL [28]. We performed two meta-analysis scans for asthma*_all_*: (I) across all study populations (META), including variants in the UKB-EUR and at least one non-EUR population; (II) across non-EUR populations (non-EUR-META), including variants in the BBJ and at least one other non-EUR population. We accounted for sample overlap using the development version of METAL for the cross-trait analysis of asthma*_all_*-WBC traits in the non-EUR populations and assessed heterogeneity in effect estimates across datasets using Cochran’s Q test and I^2^ statistics.

### 6.2 Multiple testing correction for WBC traits

To define population-specific p-value thresholds, we estimated the effective number of independent tests in the study region (*Me f f_G_*) as the number of eigenvalues that explained 99.5% of the variation in SNP data for each study population [76]: EUR-2,858; AFR-5,656; SAS-2,782; EAS-1,756. Similarly, we estimated the effective number for 11 WBC traits (*Me f f_WBC_*), finding: EUR-4; AFR-4, SAS-4; EAS-4. We determined the adjusted p-value threshold as 0.05/(*Me f f_G_*× *Me f f_WBC_*) [76], yielding EUR-7.0×10^−5^; AFR-3.5×10^−5^; SAS-7.2×10^−5^; EAS-1.0×10^−4^.

#### 6.2.1 Conditional association analysis

To identify distinct, independent association signals, we performed conditional analysis using GCTA-COJO [24]. We invoked the “–cojo-slct”, “–cojo-wind 1000” and “–cojo-p 1e-4” arguments to perform stepwise model selection using unrelated asthma controls as LD reference panels (EUR, *N* = 50,000; AFR, *N* = 8,000; SAS, *N* = 7,000; EAS, *N* = 2,000; BBJ, *N* = 20,000), using the PLINK [72] argument “–king-cutoff 0.0884” to prune out first and second-degree relatives [77]. For the meta-analysis scans, the largest populations were used as reference panels for conditional analysis (i.e., EUR for META and BBJ for non-EUR-META) as previously recommended [24].

### 6.3 Bayesian statistical fine-mapping analysis

SuSiE [29] and FINEMAP [30] were used to prioritize causal variants across traits and study populations. We computed in-sample LD matrices in unrelated asthma controls (see approximate and conditional analysis section) using LDstore [78]. The maximum number of causal variants was set to 10 for both algorithms using the default uniform prior probabilities of causality. Both algorithms computed posterior inclusion probabilities (PIP) for each variant being the true putative causal variant. Variants were then ranked in descending order of PIPs for each signal until the cumulative sum of their PIPs ≥ 0.95 to obtain 95% credible (CS95s).

### 6.4 Bayesian colocalization analysis

We identified shared causal variants between all possible asthma-WBC trait-pair or WBC trait-WBC trait-pair combinations across study populations using the COLOC R-package (COLOC-SuSiE)[32]. We obtained CS95s of variants that reported the highest posterior probabilities of being causal across each trait-pair analysed (H4 PP *>*0.6). We also performed multi-trait colocalization in each population using HyPrColoc [33]; which allowed for the assessment of colocalization across multiple traits simultaneously accounting for sample overlap and correlation between traits (see **Methods** section in the **Supplementary Materials**)

### 6.5 Mediation analysis

We assessed whether WBC traits mediated the effects of variants displaying strong asthma associations using the Regmedint R-package[79]. Using the counterfactual framework, we screened for mediation for each WBC trait under a logistic outcome model for asthma allowing for the incorporation of exposure-mediator interactions [34, 80]. SNP genotypes were considered as the exposure variables under a dominant genetic model (i.e., X = 0,1,1) for the asthma risk alleles across study populations (see **Methods** section in the **Supplementary Materials**). The additive model was also evaluated as a sensitivity analysis.

### 6.6 Summary data-based Mendelian Randomization analysis and prioritizing variants using RegulomeDB

We assessed whether gene expression mediated the effect of asthma independent signals on asthma and WBC traits using the SMR package [35]. We used cis-eQTL variants as the instrumental variables, gene expression levels as the exposure and asthma phenotypes and WBC traits as the outcome (UKB-EUR). We restricted our analyses to the study region and extracted results for the asthma independent signals and their high LD proxies (listed in **Table S27**) using the V8 release of the GTEx cis-eQTL summary data [36] across 49 tissues and in peripheral blood tissues only from the CAGE dataset [37] (except for GTEx: S8—rs8067124; CAGE: S8—rs112401631; S4—rs11078926 and S4—rs2305480 in both datasets that were not available). Tissues with the most significant variant-gene pairs were prioritized (P*_eQTL_* ≤ 5.0×10^−8^; P*_SMR_ <*0.05/m; m is the number of probes tested in the study region per tissue; see **Table S23**).

To differentiate between pleiotropy and linkage in the significant SMR associations, we performed the HEIDI test [35]. The test was carried out to test the hypothesis that a single variant affects both gene expression and trait variation, with the alternative hypothesis suggesting significant SMR findings were influenced by linkage-related associations. We used the conservative unadjusted P *>*0.05 from HEIDI to declare that pleiotropy likely influences the main SMR finding (rejecting gene-probes P *<*0.05 for linkage related associations). We implemented the default settings; removing SNPs in very strong LD (*r*^2^ *>*0.9) with the top associated eQTL, and SNPs in low LD (*r*^2^ *<*0.05) with the top associated eQTLs; PeQTL *<*1.5×10^−3^; number of cis-SNPs ≥ 3 for the HEIDI as the HEIDI test loses power if cis-SNPs *<*3 [35]; with the maximum number of eQTLs for the HEIDI test set at = 20. We also used a false discovery rate (FDR = 1.0×10^−3^) to adjust for multiple testing.

We prioritized asthma signals using the RegulomeDB ranking and scoring scheme [38]. Ranks were assigned based on the availability of supporting regulatory data types for a specific variant (i.e., ChIP-seq or eQTL data); with lower ranks indicating more evidence of a variant being located in a functional region, while scoring was determined using a tissue-specific prediction algorithm. The algorithm utilizes experimental data from variants located within biochemically characterized regions, along with information from published literature, to predict the likelihood of a variant being regulatory. The scoring scheme ranged from 0 to 1, with a score of 1 indicating a higher probability of the variant influencing gene expression.

## Declarations

### Ethics approval and consent to participate

Not applicable

### Competing interests

All authors declare no competing interests.

### Funding

This work was financially supported by the Queen Elizabeth II Diamond Jubilee Scholarship program (QEII) and the McGill Genome Centre

### Contributions

A.V.G. conceived the study. G.M.L. participated in an initial and a final discussion. A.V.G. and C.B.E. developed the methodological analysis plan. S.G. and A.D.P. provided input on assigning population labels for non-EUR UKB participants using UMAP-HDBSCAN. A.D.P. performed the clustering analysis and generated plots for the UMAP-HDBSCAN clustering. M.M. helped in data curation for statistical analysis. C.T. provided input and advice on statistical analysing using data from the BBJ. C.B.E. implemented the computational software and performed statistical analyses. A.V.G. and C.B.E. analyzed the results. C.B.E. wrote a preliminary draft and A.V.G. and wrote the draft of the manuscript. All authors critically reviewed the final version of the manuscript.

## Data Availability

All data produced in the present study are available upon reasonable request to the authors

## Acknowledgments

Many thanks to Celia Greenwood for critically reviewing the manuscript. We are grateful to all participants of the UK Biobank and the Biobank Japan project for providing their genetic data, their funding agencies and the teams who generated and assembled the datasets. This research was conducted with the UK Biobank Resource under application 6728. We would also like to thank Ioannis Ragoussis for the useful feedback during discussions about this project; Shadi Zabad, Melissa Spear, Luke Anderson-Trocmé and the members of the Gravel lab for useful discussions on programming and data access; Yoichiro Kamatani, Masaru Koido and the members of the Laboratory for Statistical and Translational Genetics-Riken IMS for their technical support and constructive feedback at various stages of this project.

## Data and Code Availability

The analysis code is available at the manuscript’s GitHub Repository.

## Resources

SAIGE (v0.35.8.3): https://github.com/weizhouUMICH/SAIGE

BOLT-LMM (v2.3.5): https://alkesgroup.broadinstitute.org/BOLT-LMM/BOLT-LMM_manual.html

LDSC (v1.0.1): https://github.com/bulik/ldsc

GCTA-COJO (v1.93.3beta2): https://yanglab.westlake.edu.cn/software/gcta/#COJO

COLOC (v5.1.0): https://chr1swallace.github.io/coloc/articles/a06_SuSiE.html

Regmedint (v1.0.0): https://kaz-yos.github.io/regmedint/

FINEMAP (v1.4): http://www.christianbenner.com/

LDstore (v2.0): http://www.christianbenner.com/#

SuSiE (v0.11.93): https://stephenslab.github.io/susieR/index.html

HyPrCOLOC (v1.0): https://github.com/jrs95/hyprcoloc

METAL: http://csg.sph.umich.edu/abecasis/Metal/download/

METAL development version: https://github.com/statgen/METAL

FUMA: https://fuma.ctglab.nl

SNPCLIP: https://ldlink.nci.nih.gov/?tab=apiaccess

PLINK: https://www.cog-genomics.org/plink/

UMAP: https://github.com/lmcinnes/umap

HDBSCAN: https://github.com/scikit-learn-contrib/hdbscan

LocusZoom: https://my.locuszoom.org

LDLINK: https://ldlink.nci.nih.gov/?tab=home

SMR: https://yanglab.westlake.edu.cn/software/smr/

## Notes

### Competing Interest Statement

The authors have declared no competing interest.

### Author Declarations

The data used in this study were accessed from the UK Biobank under approved application number 6728

