## Supplementary_Materials for "Multi-ancestry fine-mapping of the chromosome 17q12-q21 asthma locus identifies independent associations implicating lymphocyte and eosinophil levels in the causal pathway"

Chief Ben-Eghan^1,2^, Markus Münter^2^, Chikashi Terao^3^, Alex Diaz-Papkovich^2,4^, Simon Gravel^1,2^, G. Mark Lathrop^1,2^, Audrey V. Grant* ^1,5^

^1^Department of Human Genetics, McGill University; Montreal,
QC, H3A 0G1, Canada.

^2^McGill University and Genome Quebec Innovation Centre; Montreal,

QC, H3A 0G1, Canada.

^3^Laboratory for Statistical and Translational Genetics, RIKEN Center for Integrative Medical

Sciences;

Yokohama, Japan.1-7-22 Suehiro-cho, Tsurumi-ku, Yokohama, Kanagawa,

230-0045, Japan.

^4^Quantitative Life Sciences, McGill University; Montreal,

QC, H3A 0G1, Canada.

^5^Faculty of Dental Medicine and Oral Health Sciences, Department of Anesthesia, Alan Edwards Centre for Research on Pain, McGill University; Montreal,

QC, H3A 1G1, Canada.

*Corresponding author

Methods

#### **Assigning population labels using UMAP-HDBSCAN clustering algorithm**

We obtained information on the selected ethnicity of participants using the UK biobank (UKB) data-field “21000” which organises participant ethnicity into a hierarchical tree structured dictionary for all 488,377 genotyped participants. The dataset is subsequently grouped into 4 main categories based on selected ethnicity, with the majority being White (88.3% British, 2.6% Irish, 3.4% Any other white background), with large populations identifying as Black (1.6% either African, Caribbean, or Any other Black background), Asian (1.9% either Indian, Pakistani, Bangladeshi, or Any other Asian background), mixed ethnicity (0.6%), Chinese (0.3%), other ethnic group (0.8%), or an unavailable response (0.5%) (See Additional file1: Table S1 for the breakdown of UKB participants by selected ethnicity). It is worth mentioning that in the UKB, the Asian category is assigned to participants of South-East Asian descent.

We assigned the “EUR” population label to participants that selected White-British ethnicity in the UKB (*N* = 431, 110). Despite having a smaller proportion of non-EUR participants, the UKB is still one of the largest, open access biobanks for individuals of non-EUR ancestry. Individuals of admixed ancestry or those who select “Mixed” or “Other” ethnicity from the UKB are seldom included in analyses to reduce confounding due to population structure and because they are often statistically underpowered for discovery. There are reasonable justifications for excluding participant data from analysis pipelines [1], however, to fully exploit this data resource, we sought to incorporate these often-discarded populations in our analyses focusing on the non-EUR populations from the UKB.

We applied Uniform Manifold Approximation and Projection (UMAP)[2, 3] in conjunction with the clustering algorithm Hierarchical Density-Based Spatial Clustering of Applications with Noise (HDBSCAN)[4] on the full complement of UKB data. While dimensionality reduction methods like PCA aim to preserve large-scale structure by identifying the maximal variance in a given data set, ignoring variation along other directions, UMAP focusses on preserving local neighbourhoods within a data set by grouping individuals according to similar genetic distances[5, 6]. The initial step we performed was dimension reduction using UMAP, where we reduced the top 20 genetic principal components of the full UKB genotype data to 3 dimensions, specifying the 150 nearest neighbours and a minimum distance of 0.0001 to ensure dense cluster formation. HDBSCAN was parameterized with a minimum cluster size of 50 individuals and an epsilon value of 0.5, resulting in 16 mutually exclusive clusters across the entire UKB, with six individuals remaining un-clustered.

Using this data-driven agnostic approach, we were able include non-EUR individuals that are often filtered out or excluded from analyses and re-assign them based on the clusters formed to their continental groups i.e., African (AFR), South-Asian (SAS) and East-Asian (EAS). Compared to methods using principal component analysis or using selected ethnicity only (Additional file1: Table S1), UMAP-HDBSCAN led to an increase in sample size by 2,416 for AFR (↑ 32%), with a slight decrease in SAS by 138 samples (↓ 1.4%) (See Additional file1: Table S1, Additional file1: Figs. S1-S3 for a summary of the clustering process described above). After applying the clustering algorithm, we were able to track individuals that clustered closer to their continental groups from the non-EUR populations using the selected ethnicity variable.

In the UKB-AFR where we experienced the most significant increase in sample size, we used the genomic inflation factor and Q-Q plots to assess the genome-wide distribution of the test statistic compared to the expected null distribution in the newly assigned AFR cluster. To do this, we performed genome-wide scans for the counts of white blood cell traits in UKB-AFR defined by 1) AFR-selected ethnicity only and 2) UMAP-HDBSCAN assigned AFR cluster, using the same covariates (See phenotype definition section below for how blood cell traits were modelled). The increase in sample size in the UMAP-HDBSCAN assigned AFR cluster did not result in inflation of the genomic inflation factor statistic and exhibited approximately equal values when compared to AFR based on selected ethnicity only (Additional file1: Fig. S4).

#### **Phenotype definition**

We selected participants with asthma from the UKB using information from two data fields: data field 6152 (“has a doctor ever told you that you have had any of the following conditions?” asked on a touchscreen questionnaire) and data field 20002 (verbal interview, self-reported non-cancer illness). Affected individuals reported “asthma” to data field 6152 (coding variable 8) with an assigned code for asthma (coding variable 1111) in data field 20002 (*N*=50,264 cases). Self-reported information on the age of asthma onset was obtained using the data-field 3786 (“What was your age when the asthma was first diagnosed?” asked on a touchscreen questionnaire). Participants who did not report a diagnosis of asthma in data field 6152 (8), were designated as controls (*N*=380, 581), while those who selected “prefer not to answer” were excluded and marked as missing. We subsequently stratified asthma cases and controls for the UKB subpopulations, using the UMAP-HDBSCAN defined clusters presented in the previous section, to facilitate downstream analysis. A full breakdown of the counts of cases and controls can be found in the Additional file 2: Table S1. In the BBJ asthma cases and controls were defined using protocol presented by Nagai *et al*.[7]. Briefly, all patients with bronchial asthma were diagnosed by physicians at cooperating institutions throughout Japan (66 hospitals and 12 medical institutes). Exclusion of participants who were not of East-Asian descent or had received a bone-marrow transplant was carried out prior to harmonising datasets across medical centres. Following sample level qc filters such as excluding individuals with greater than 10% missing genotype data, a total of 7,456 asthma cases were available for downstream analysis. Non-asthmatic controls were individuals without a doctor diagnosed asthma record, drawn from participants across the contributing institutions (*N*=153,654)

Data for white blood cell (WBC) traits in the UKB were obtained from the following data-fields: basophil count (30160-0.0), basophil percentage (30220-0.0); eosinophil count (30150-0.0), eosinophil percentage (30210-0.0); lymphocyte count (30120-0.0), lymphocyte percentage (30180-0.0); monocyte count (30130-0.0), monocyte percentage (30190-0.0); neutrophil count (30140-0.0), neutrophil percentage (30200-0.0); total white blood cell count (30000-0.0). Differential white blood cell percentages (%) and the total WBC count (WBC#) were measured directly with a haematology analyser. The absolute counts (#) of the other white blood cells are derived from the percentages and WBC#; e.g. to calculate LYM#, we use (LYM % x WBC#)/100% (see additional details here [8]; see also Table S1 of Astle et al [9]). We modelled WBC traits following the approach used by Mousas *et al*.[10]. Briefly, within each UKB study population, we adjusted for the following covariates: age, sex, smoking status, assessment center where the blood samples were collected, machine counter that processed the blood samples, month, day of the week, time of day samples were collected, selected ethnicity of participants, height, weight, and top 10 PCs. Regression residuals were normalized using rank-based inverse normal transformation (INT) to the standard normal distribution for each UKB population. WBC traits in the BBJ were modelled using methods described by Kanai *et al*.[11]. Data was available for only the counts of WBC traits in BBJ participants (basophil, eosinophil, lymphocyte, monocyte, neutrophil and total white blood cell counts). We adjusted for blood cell indices using fewer covariates (age, sex, top 10 PCs) as there was not as much granular information to use for covariate adjustment as was present in the UKB. We normalized residuals after adjustment similarly using INT. Counts and percentages of WBC traits were designated with “#” and “%” respectively.

#### **Simulations using data from BCX consortium**

To explore whether the asthma-17:37.2-38.8Mb region contained an excess of independent signals compared to the total set of GW signals present for each WBC trait, we simulated a null hypothesis distribution for the count of signals in similarly sized regions (1.6Mb). We first obtained meta-analysis summary statistics for the counts of basophils (BAS#), eosinophils (EOS#), lymphocytes (LYM#), monocytes (MON#), neutrophils (NEU#) and total white blood cells (WBC#) for individuals of European (EUR, *N* = 563,946), African (AFR, *N* = 15,171), East-Asian (EAS, *N* = 151,807) and Hispanic (HIS, *N* = 9,368) ancestry from the BCX consortium, which published the largest trans-ethnic study on blood cell phenotypes to date (http://www.mhi-humangenetics.org/en/resources)[12]. We first retained variants that passed a p-value filter (EUR, *P* < 5 × 10^−8^; non-EUR, *P* < 1 × 10^−4^) for all WBC traits. We obtained independent signals using the R version of SNPCLIP [13] by pruning out variants with MAF < 0.02 for each ancestry (pairwise LD *r*^2^ > 0.2) using the corresponding 1000 Genomes reference population (Phase 3). We excluded the HLA region (chr6:29,691,116–33,054,976, hg19) across all populations because of the extensive LD structure in this region. In the AFR and HIS populations, we excluded the Duffy/DARC region in chr1 known to be enriched for GWS signals associated with benign ethnic neutropenia (chr1:158,724,683- 159,624,683, hg19)[12, 14].

We then generated 10,000 random overlapping segments (~454 segments per chromosome) each 1.6Mb in size across all autosomes using the GenomicRanges R-package[15]. We populated each bp interval of the 10,000 segments generated using the count of independent GW significant signals from the pruned list (EUR, *P* < 5 × 10^−8^; non-EUR, *P* < 1 × 10^−4^) to serve as our null distribution. For each population, we calculated empirical p-values per WBC trait by first obtaining the count of GW significant variants in the asthma-17:37.2-38.8Mb region and then sorted the counts of GW significant variants in each bp interval in our null dataset in descending order. Empirical p-values were obtained by dividing the number of observations before obtaining the count of GW significant variants in the in asthma-17:37.2-38.8Mb region in the null dataset by 10,000.

To provide a benchmark for comparison, we applied the same signal counting and empirical p-value framework to a larger region spanning 5q31-q33 (chr5:122.06-174.37 Mb; 52.3 Mb), selected for its established role in immune regulation and inclusion of multiple interleukin genes (e.g., *IL3*, *IL4*, *IL5*, *IL13*). Despite its larger genomic span, this region yielded fewer and less consistent enrichments than 17q12-q21 across WBC traits.

#### **Bayesian colocalization analysis**

We ran colocalization analysis using the latest version of COLOC (COLOC-SuSiE; version 5.1.0)[16] on all possible asthma-WBC trait-pair or WBC trait-WBC trait-pair combinations across study populations. We used the global colocalization hypothesis to determine trait-pairs showing evidence of colocalization: *H0*: neither trait had a genetic association in the region; *H1/H2*: only trait 1/trait 2 had a genetic association in the region; *H3:* both traits were associated with different causal variants and *H4:* both traits were associated and shared a single causal variant. We reported trait-pair-causal variant combinations that favoured H4 (PP > 0.6). We used the following as input parameters: effect estimates (BETAs), variance of the effect estimates (SE^2^), proportion of case samples for asthma phenotypes (s), sample size (*N*) for WBC traits, MAF and population-specific in-sample LD. For each trait-pair analysed, we obtained the 95% credible sets (CS95s) of variants that reported the highest posterior probabilities of being causal across the trait-pair to support the H4 shared causal variant hypothesis (H4 PP > 0.6). In instances where COLOC-SuSiE did not generate CS95s in both traits in the trait-pair, we reverted to the single causal variant assumption implementation as suggested by Wallace *et al*.[16]. We also performed multi-trait colocalization using HyPrColoc (version 1.0)[17]; which implements the single causal variant assumption but allows for the assessment of colocalization across multiple traits simultaneously. HyPrColoc also accounts for sample overlap and correlation between traits. We analysed all asthma and WBC traits together in each population using the following as input parameters: effect estimates (BETAs) and their standard errors (SE) in addition to the in-sample LD. We distinguished between the asthma phenotypes and the WBC traits using the “binary.outcomes” and the “trait.names” arguments respectively and specified the “sample overlap” and “snp.scores” arguments to estimate the posterior probability of colocalization explained by each SNP.

#### **Mediation analysis**

We conducted mediation analysis to assess whether variants in the study region had their effects mediated through a WBC trait via the Regmedint R-package (version 1.0)[18]. Using the counterfactual framework, we screened for mediation using a single mediator (WBC trait) under a linear mediator, logistic outcome model which allowed for the incorporation of exposure-mediator interactions and binary outcomes[19, 20]. We screened for mediation in 1,187 variants across the four AO-related strata with p-values ≤1 × 10^-4^ in the UKB-EUR. We used the AO-related asthma phenotypes as outcome variables and SNP genotypes as exposure variables under a dominant genetic model for the asthma risk allele in each AO-related stratum. (i.e., X=0,1,1). For the non-EUR populations, we screened for mediation in all variants that passed QC, using the asthma_all_ as the outcome variable. We set the exposure-mediator interactions argument to “true” and obtained confidence intervals and estimates for the statistical significance of the direct, indirect and total effects, together with the proportion of the total effect of each SNP mediated through a WBC trait (proportion mediated). We filtered out variants where the direct and indirect effects were inconsistent (in opposite directions) and retained variants with indirect effect p-values ≤ 7 × 10^-5^ (See multiple testing correction in the methods for the adjusted significance levels for the UKB populations). Finally, we performed pairwise LD clumping (*r*^2^ > 0.5, within a 1000kb window) via PLINK[21] on variants retained after the filters and assigned them into mediation signal groups (M1 to M7) ordered by base-pair position in the UKB-EUR.

### **Supplementary Tables**

Tables S1-S29 are provided in a separate Excel workbook.

### Supplementary Information Tables

#### Table S1: Selected ethnicity of participants in the UK biobank.

| Selected Ethnicity | *N* | Category |
| --- | --- | --- |
| White | 546 | **White** |
| British | 431,110 | **(460,238)** |
| Irish | 12,760 |  |
| Any other White background | 15,822 |  |
| Black or Black British | 26 | **Black** |
| African | 3,206 | **(7,649)** |
| Caribbean | 4,299 |  |
| Any other Black background | 118 |  |
| Asian or Asian British | 42 | **Asian** |
| Indian | 5,716 | **(9,474)** |
| Pakistani | 1,748 |  |
| Bangladeshi | 2,221 |  |
| Any other Asian background | 1,747 |  |
| Mixed | 46 | **Mixed** |
| White and Black African | 402 | **(2,843)** |
| White and Black Caribbean | 597 |  |
| White and Asian | 802 |  |
| Any other Mixed background | 996 |  |
| Chinese | 1,504 | **Other** |
| Other Ethnic group | 4,357 | **(8,173)** |
| Do not know | 204 |  |
| Prefer not to answer | 1,586 |  |
| No Ethnicity | 522 |  |
| Total |  | **488,377** |

#### Table S2: Algorithms used and the analyses strategy.

| Algorithm | Use | Strategy |
| --- | --- | --- |
| SAIGE | Genetic association testing for binary phenotypes accounting for case-control imbalances. | asthma_all_ across all study populations |
| BOLT-LMM | Genetic association testing using linear mixed models accounting for fixed and random effects. | AO-related strata in UKB-EUR, WBC traits across all study populations |
| METAL | Fixed-effects inverse variance weighted meta-analysis. | asthma_all_ across all study populations |
| GCTA-COJO | Iterative conditional stepwise analysis to identify independent association signals using LD estimates between variants. | All asthma phenotypes & WBC traits, across all study populations (All) |
| FINEMAP | Statistical fine-mapping using shotgun-stochastic search for prioritizing putative causal variants using in-sample LD. | All |
| SuSIE | Bayesian statistical fine-mapping using summary statistics and LD information under the multiple causal variant assumption. | All |
| COLOC-SuSIE | Bayesian colocalization analysis between two trait-pairs under the multiple shared causal variant assumption. | All |
| HyPrColoc | Bayesian colocalization analysis across multiple traits simultaneously (>2traits), under the single shared causal variant assumption. | All |
| Regmedint | Regression based causal mediation analysis accounting for exposure–mediator interactions. | All |
| SMR | Summary-data-based Mendelian Randomisation to test for pleiotropic associations between the expression level of a gene and a complex trait. | All |

#### Table S3: Counts of genome-wide significant variants in the chr17q12-q21 & chr5q31-33 regions for WBC traits.

| Population | Basophil | | Eosinophil | | Lymphocyte | | Monocyte | | Neutrophil | | Total WBC | |
| --- | --- | --- | --- | --- | --- | --- | --- | --- | --- | --- | --- | --- |
| EUR | **24** | 21 | **17** | 156 | **17** | 128 | **12** | 73 | **72** | 48 | **79** | 123 |
|  | **(3.3x10^-03*^)** | (1.3x10^-01^) | **(2.9x10^-02^)** | (7.0x10^-03*^) | **(2.0x10^-02^)** | (1.7x10^-01^) | **(4.6x10^-02^)** | (2.9x10^-01^) | **(5.0x10^-04**^)** | (1.6x10^-01^) | **(2.0x10^-04**^)** | (5.5x10^-02^) |
| EAS | **-** | 5 | **3** | 32 | **2** | 7 | **1** | 5 | **25** | 1 | **22** | 11 |
|  | **-** | (5.3x10^-01^) | **(2.6x10^-02^)** | (2.3x10^-02^) | **(2.5x10^-02^)** | (4.1x10^-01^) | **(6.0x10^-02^)** | (6.3x10^-01^) | **(1.0x10^-04**^)** | (8.9x10^-01^) | **(1.0x10^-03*^)** | (5.8x10^-01^) |
| AFR | **-** | 2 | **3** | 7 | **1** | 3 | **-** | 9 | **2** | 12 | **3** | 16 |
|  | **-** | (8.5X10^-01^) | **(1.1x10^-02^)** | (4.1x10^-01^) | **(1.7x10^-01^)** | (7.5x10^-01^) | **-** | (3.3x10^-01^) | **(4.8x10^-02^)** | (7.6x10^-02^) | **(2.3x10^-02^)** | (5.1x10^-02^) |
| HIS | **-** | 2 | **1** | 2 | **1** | 2 | **-** | 2 | **3** | 1 | **3** | 4 |
|  | **-** | (7.1x10^-01^) | **(1.4x10^-02^)** | (7.4x10^-02^) | **(1.0x10^-02^)** | (6.8x10^-01^) | **-** | (7.4x10^-01^) | **(3.2x10^-03*^)** | (8.1x10^-01^) | **(4.2x10^-03^)** | (3.9x10^-01^) |

For each phenotype, the table shows the number of genome-wide significant variants and corresponding empirical p-values for the 17q12-q21 and 5q31-q33 regions. Empirical p-values appear in parentheses below each count. Dashes indicate no significant associations. Red font highlights cases where associations with greater significance were observed in 5q31-q33 than in 17q12-q21. Population abbreviations: EUR - European (N = 563,946), EAS – East Asian (N = 151,807), AFR – African (N = 15,171), HIS – Hispanic (N = 9,368). Significance thresholds: EUR = 5×10⁻⁸; non-EUR = 1×10⁻⁴.

*p < 0.001; **p < 0.0001

### **Supplementary Figures**

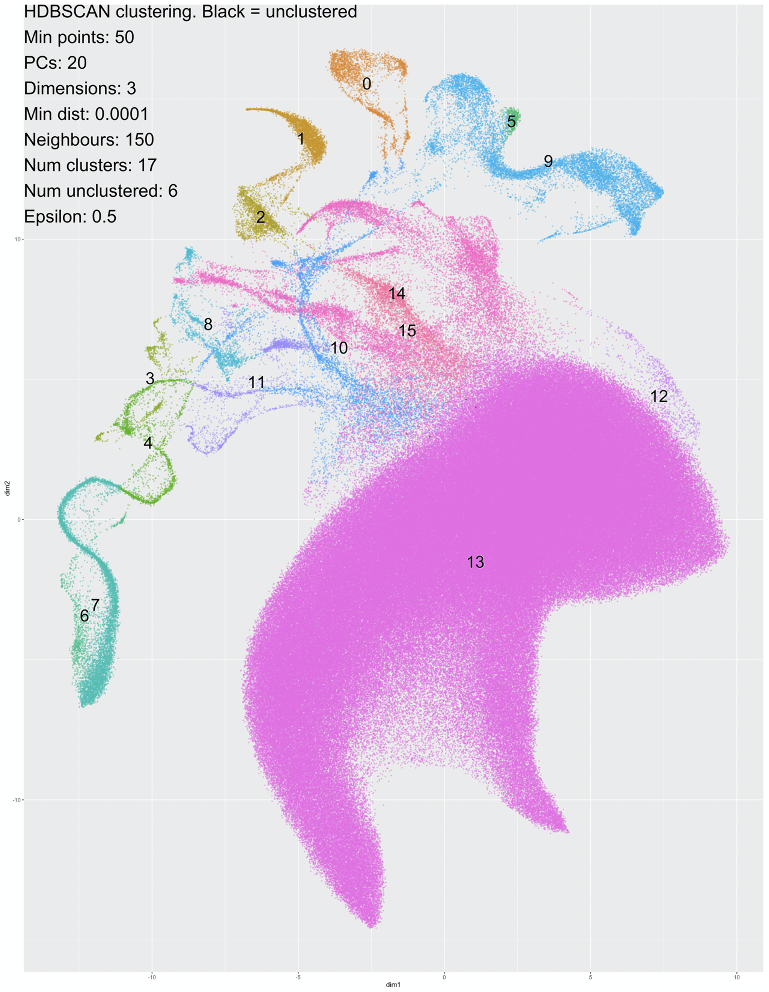

dim2

dim1

**Figure S1. Uniform Manifold Approximation and Projection and Hierarchical Density-Based Spatial Clustering of Applications with Noise (UMAP-HDBSCAN) clustering in the full UKB cohort**. Each study participant is represented as a point on the 2D UMAP plot, coloured by cluster assignment. Genotype data were pre-processed using 20 global principal components (PCA), followed by clustering on higher-dimensional UMAP embeddings using HDBSCAN, with parameters shown in the top right corner of the plot. The 17 resulting clusters are colour-coded, with clusters 3, 4, 6, and 7 representing UKB participants of African ancestry (UKB-AFR), clusters 5 and 9 representing UKB participants of South-Asian ancestry (UKB-SAS), cluster 0 representing UKB participants of East-Asian ancestry (UKB-EAS), and the remaining clusters representing populations of European ancestry.

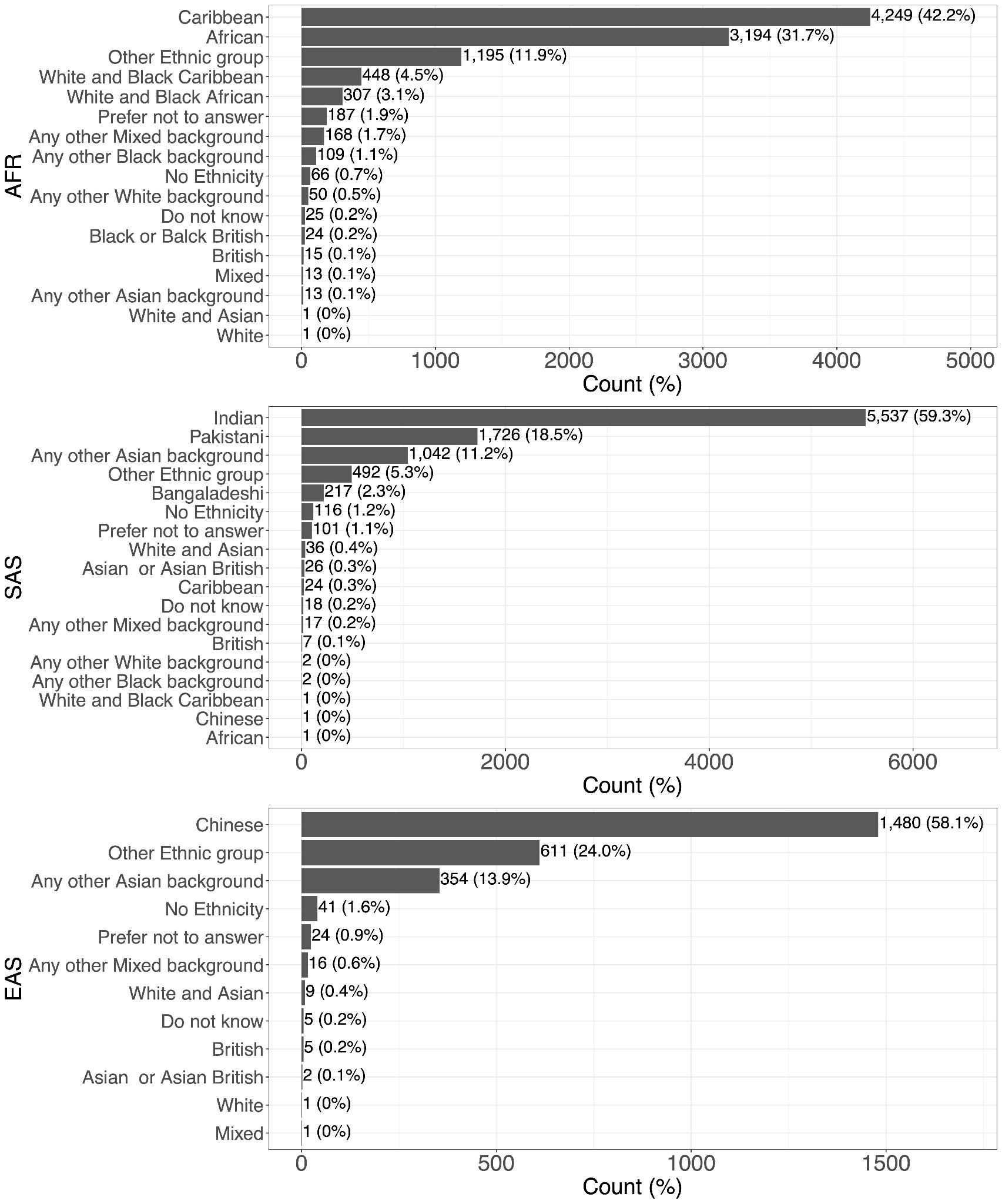

**Figure S2. Uniform Manifold Approximation and Projection and Hierarchical Density-Based Spatial Clustering of Applications with Noise (UMAP-HDBSCAN) improves sample size among UKB non-European ancestry participants**. The figure shows the number of participants assigned to non-European (non-EUR) ancestry groups following UMAP-HDBSCAN clustering. Individuals who initially selected “Mixed,” “Other,” “Prefer not to answer,” “Do not know,” or “No Ethnicity” were re-assigned to continental ancestry groups based on clustering results. Final sample sizes were: African (UKB-AFR), 10,065 participants; East Asian (UKB-EAS), 2,546 participants; and South Asian (UKB-SAS), 9,336 participants.

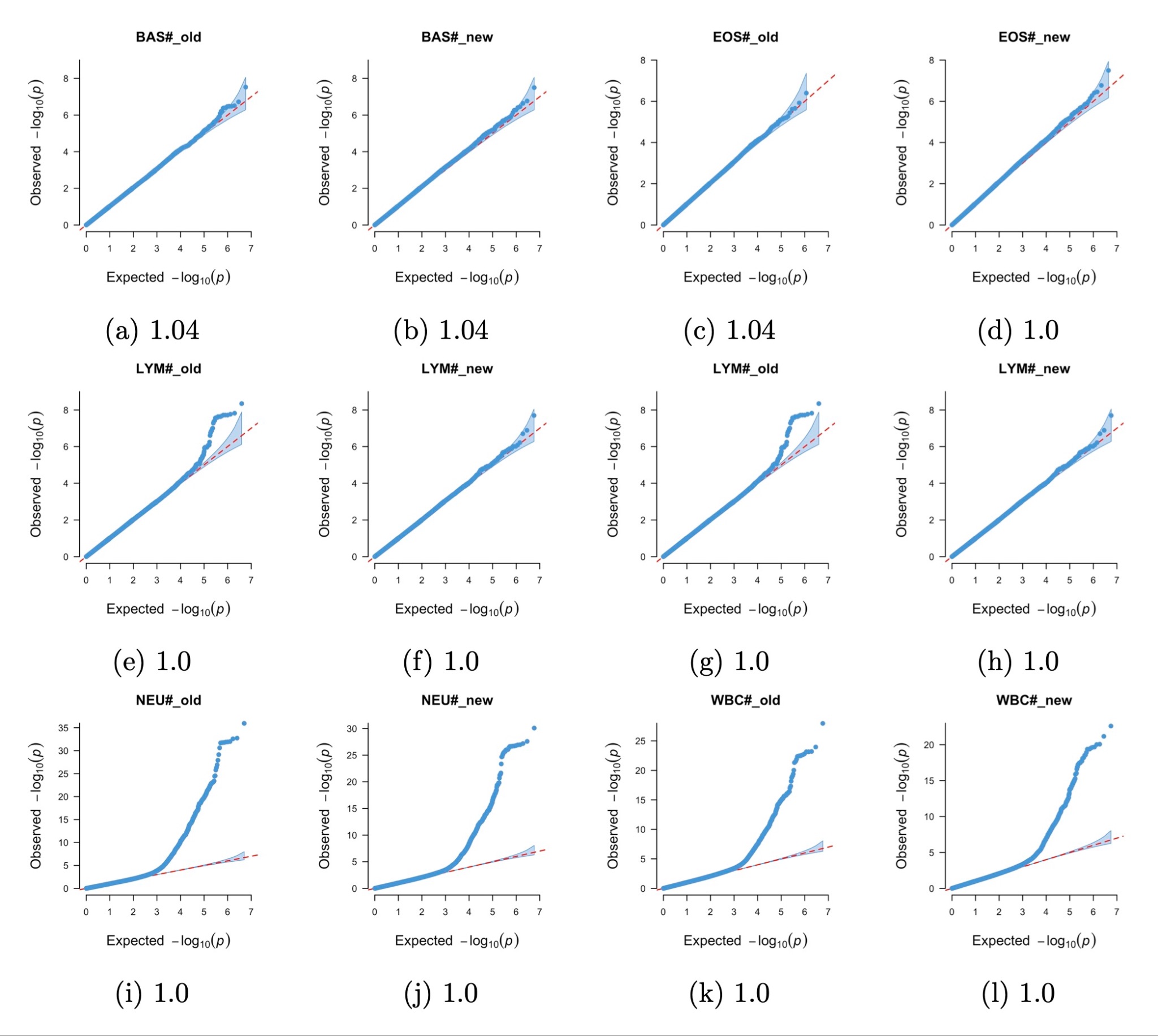

**Figure S3. Quantile-quantile (Q-Q) plots for the UKB-African (AFR) subset before and after UMAP-HDBSCAN clustering.** Q-Q plots show genome-wide association results for WBC trait counts (#) in the UKB-African (AFR) subset before ("old") and after ("new") reclassification using UMAP-HDBSCAN clustering. Genomic inflation factors (λ) for each WBC trait are displayed below each plot (**a**–**l**). "Old" refers to participants based on self-reported African ethnicity, while "new" represents individuals assigned to the AFR cluster after unsupervised clustering.

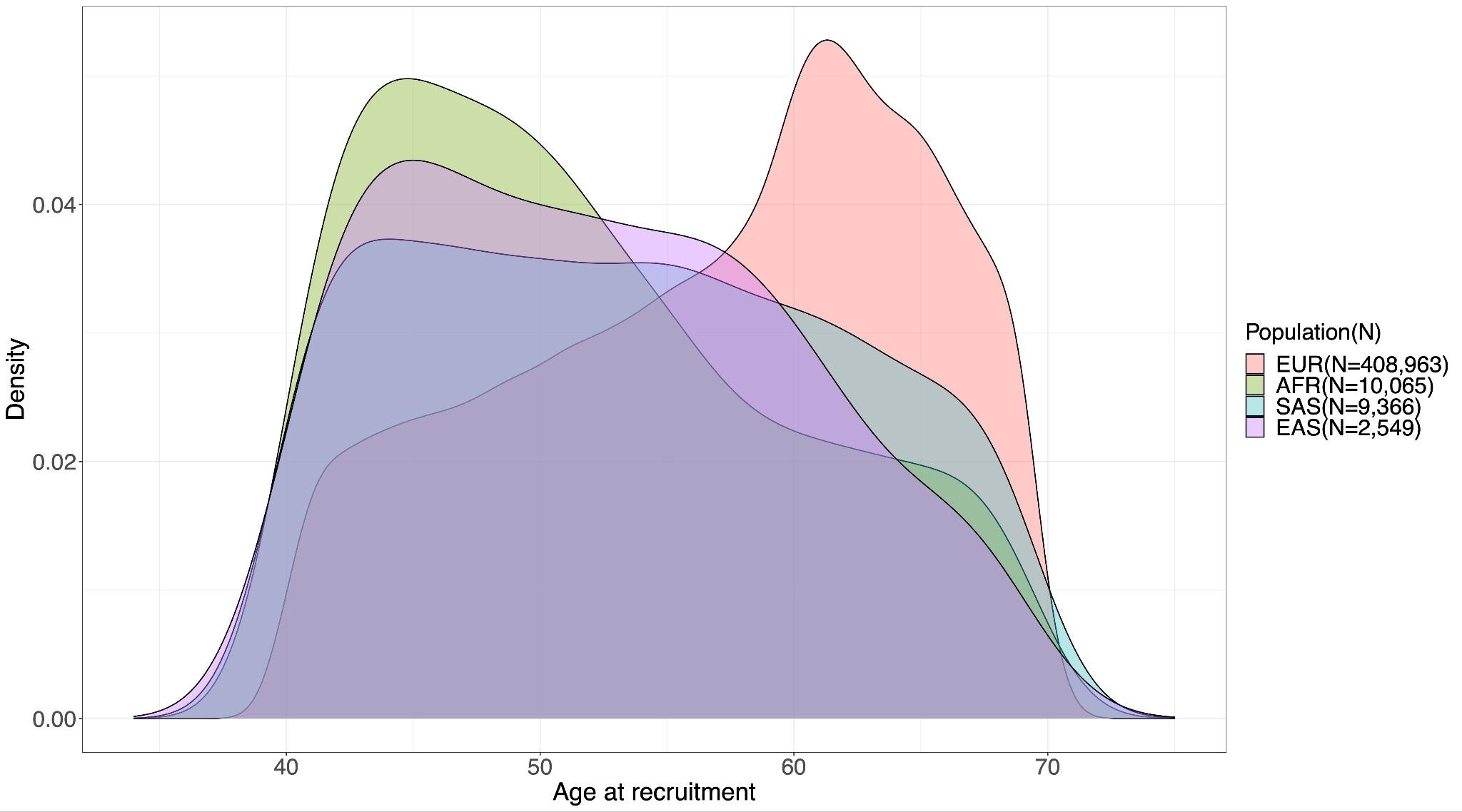

**a**

**b**

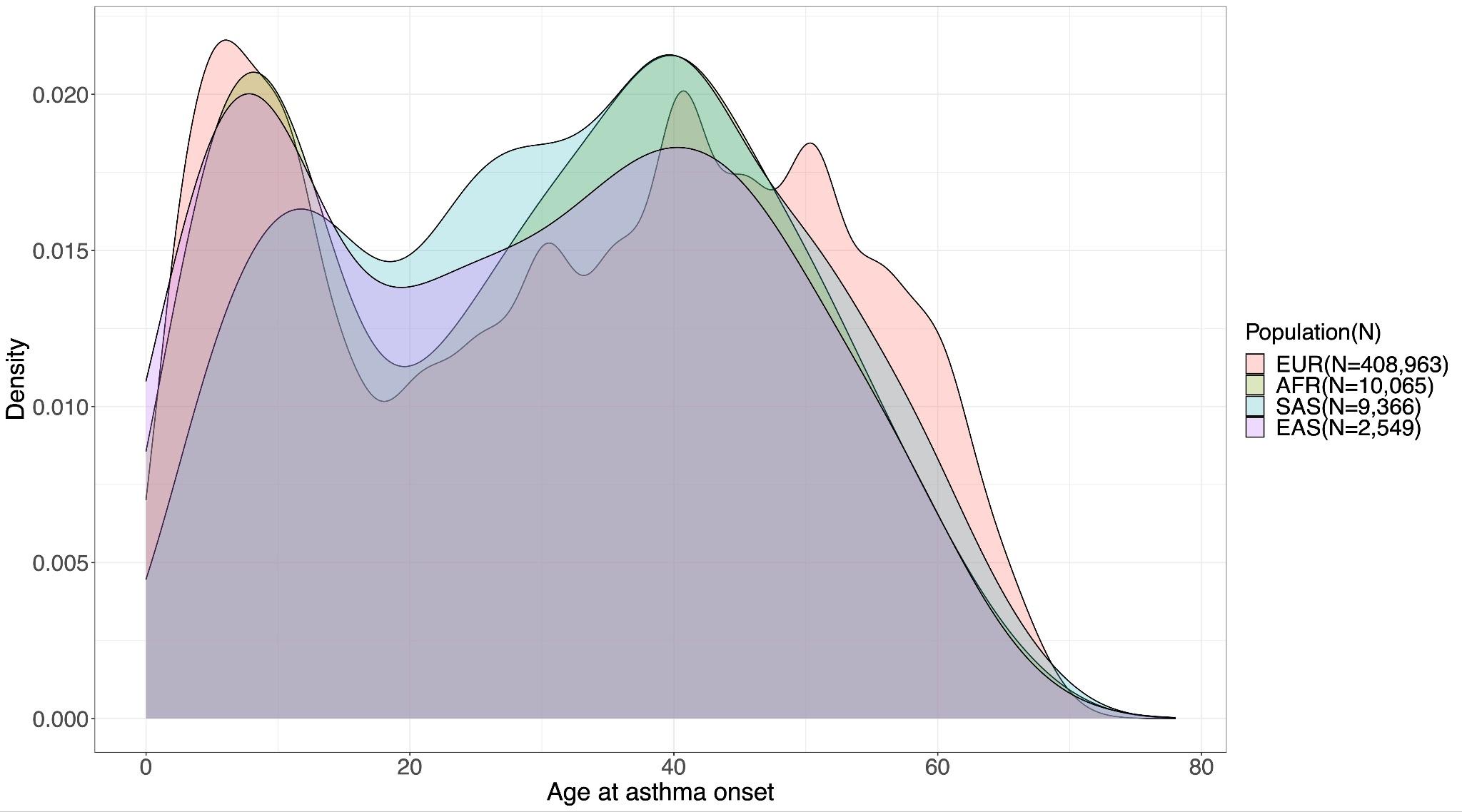

**Figure S4. Age distribution across UKB subpopulations.** (**a**) Density plot of age at recruitment (baseline) for each UKB population subset, with corresponding sample sizes indicated. The UKB subpopulations are designated by ancestry as follows: African (AFR), South-Asian (SAS), European (EUR), and East-Asian (EAS). (**b**) Density plot of age at asthma onset across UKB subpopulations.

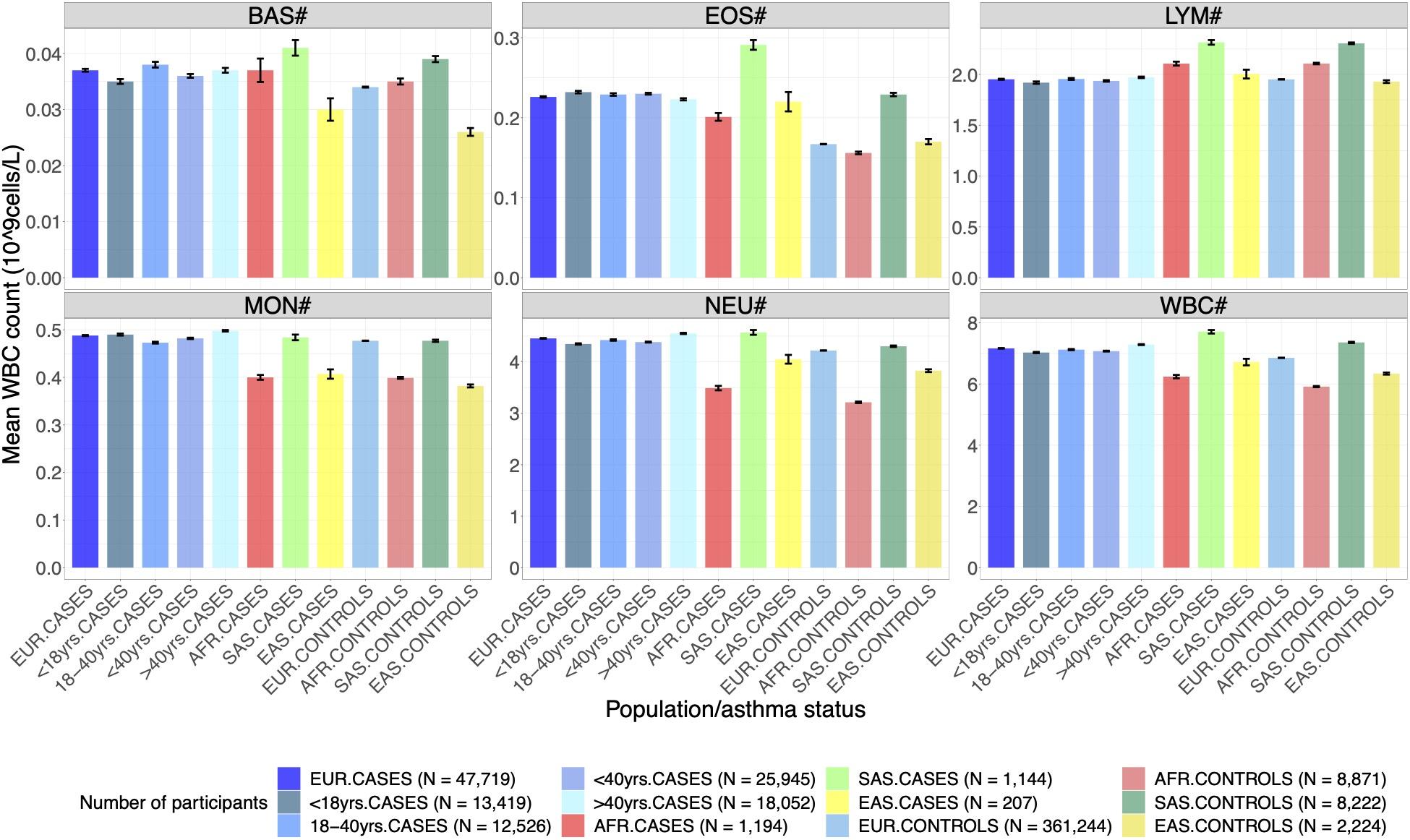

**Figure S5. Mean white blood cell (WBC) counts by asthma case-control status across UKB population subsets.** The cases for each population are plotted first, followed by the controls. The standard errors of the means are represented by the black error bars.  The figure shows mean unadjusted WBC counts stratified by asthma case-control status across UKB ancestry groups. For each population, asthma cases are plotted first, followed by controls. Black error bars indicate the standard error of the mean for each group. The counts (#) and percentages (%) of white blood cell (WBC) traits were represented as: basophils (BAS), eosinophils (EOS), lymphocytes (LYM), monocytes (MON), neutrophils (NEU), and total WBC count (WBC#)

**a**

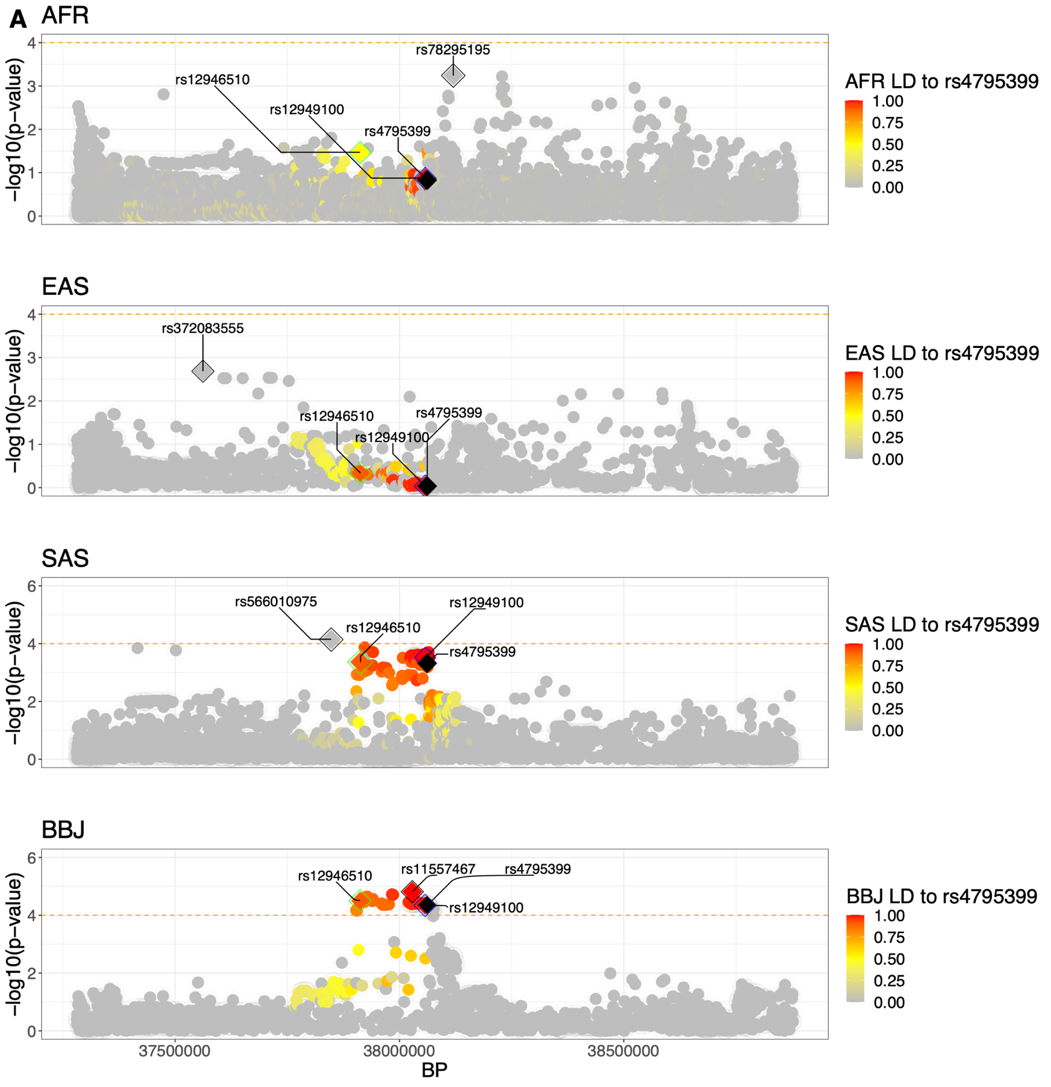

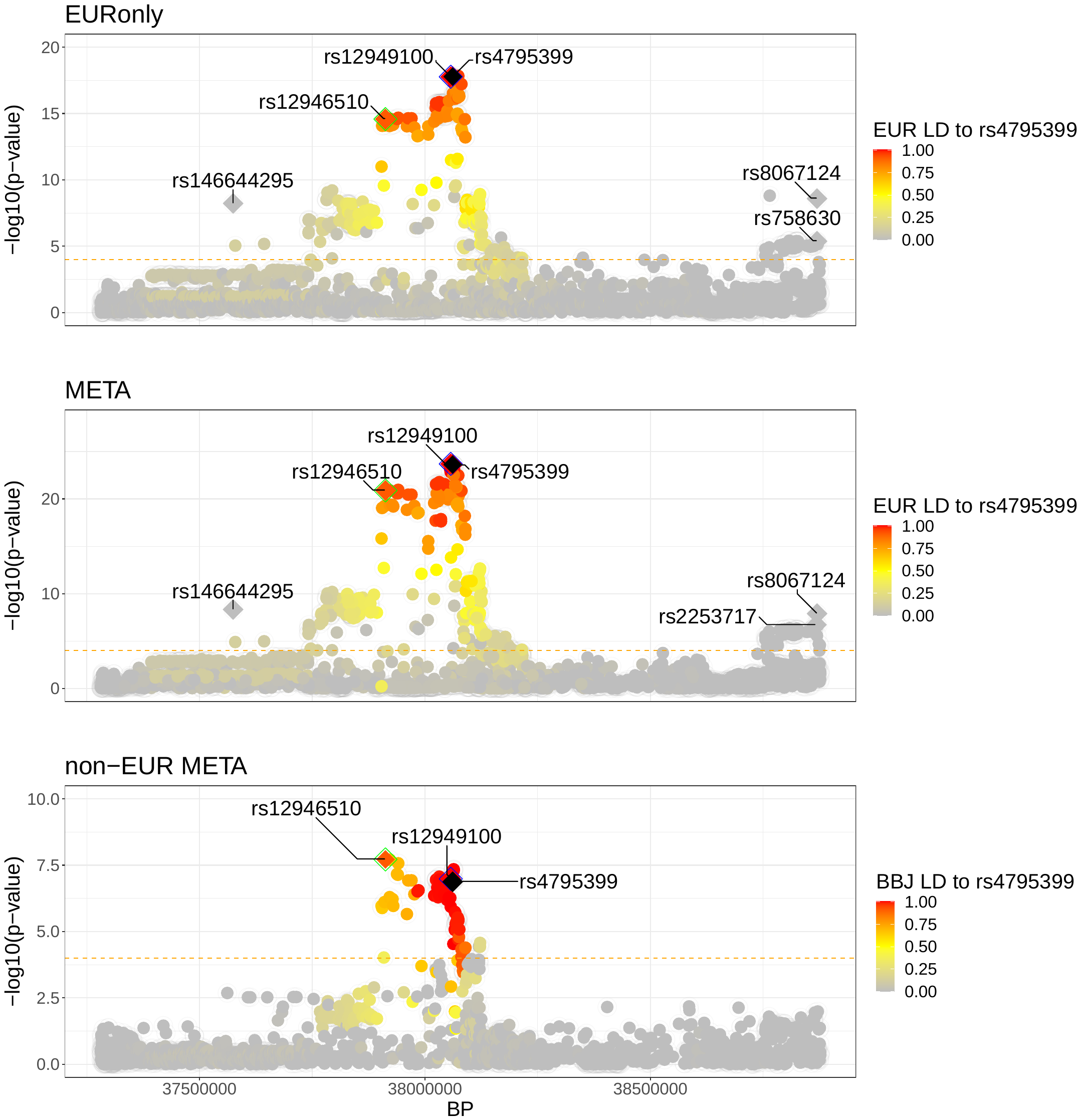

**b**

**Figure S6.** **Regional association plots for asthma adjusted for sex and age (asthma_all_) in EUR-only (UKB European ancestry), META (all study populations), and non-EUR META (all non-European study populations).** (**a**) Regional association plots showing asthma signals identified using GCTA-COJO. In the UKB-European (EUR) subset, the asthma_all_ phenotype was analysed, with signal S4--rs4795399 shown as a black diamond. Signal S4--rs12949100, the top signal from the full meta-analysis (EUR-META: European + African + South Asian + East Asian + BBJ), is shown as a diamond with a blue outline, and S4–rs12946510, the top signal from the non-European meta-analysis (non-EUR META: African + South Asian + East Asian + BBJ), is shown as a diamond with a green outline. The most genome-wide significant variant in each scan is represented by a diamond with a black outline. The dashed orange line indicates the suggestive significance threshold at P = 1 × 10⁻⁴.Variant shading corresponds to pairwise linkage disequilibrium (LD, r²) with S4--rs4795399 (see key). In the African, South Asian, and East Asian scans, the most genome-wide significant variants were not in high LD (r² < 0.2) with S4--rs4795399, S4--rs12949100, or S4--rs12946510. In contrast, in the BBJ scan, the most significant variant (rs11557467) was in high LD with S4--rs4795399. (**b**) Tracking of the S4 signals across the UKB-European-onlyasthma_all_ scan (EUR), full meta-analysis (EUR-META), and non-European meta-analysis (non-EUR META). Additional signals (S3--rs146644295, S7--rs2253717, and S8--rs8067124) are also shown; however, these did not meet the suggestive threshold in the non-EUR META analysis.

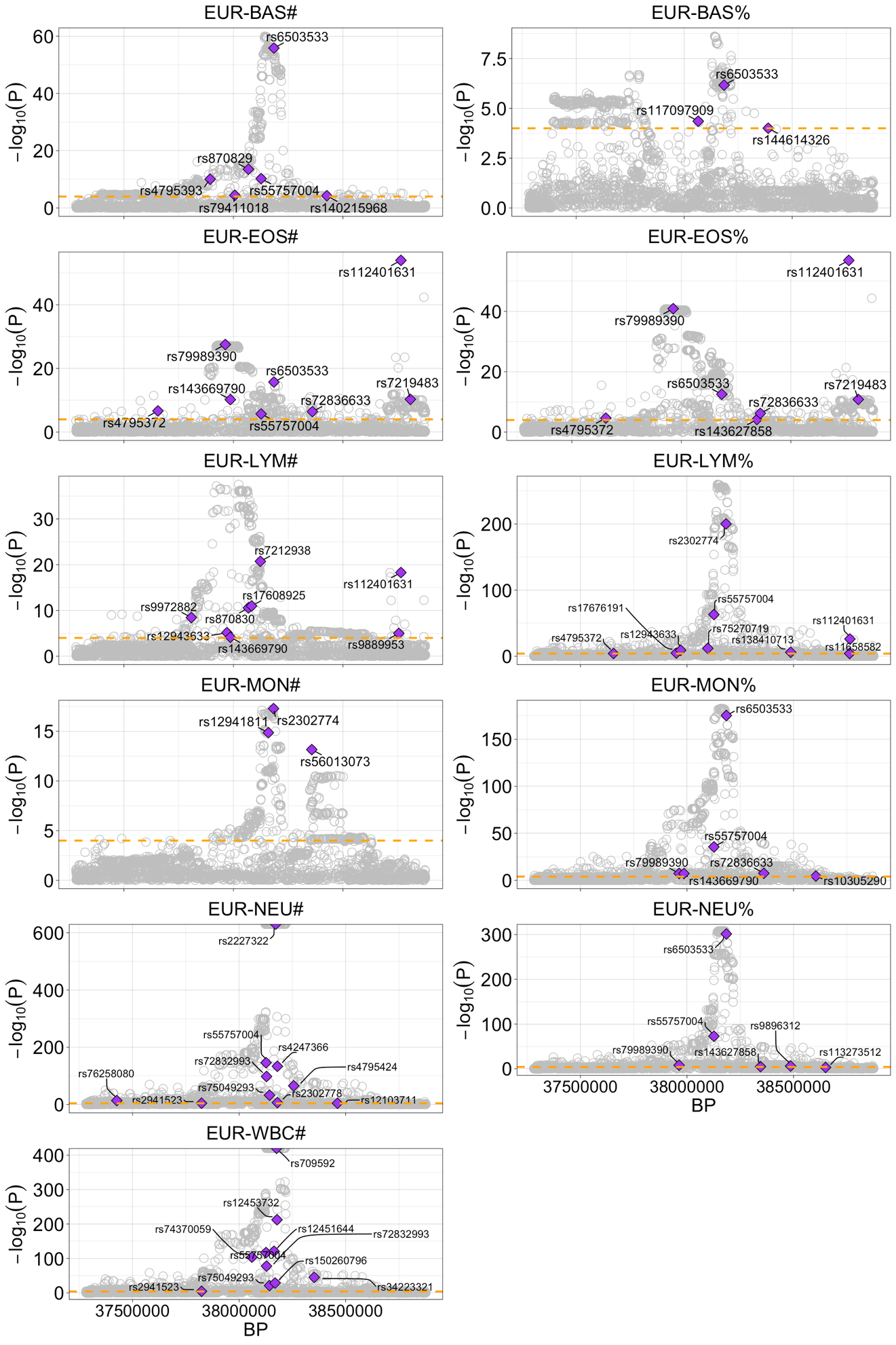

**a**

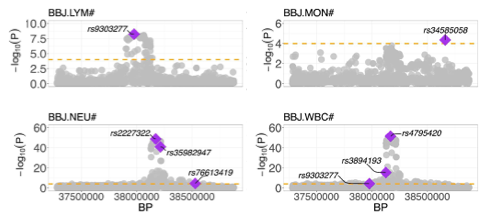

**b**

#### **Figure S7. Independent association signals in white blood cell traits.**

Regional association plots for white blood cell (WBC) traits, highlighting independent signals identified using GCTA-COJO. Purple diamonds represent independent lead variants for WBC traits in the UKB-European (UKB-EUR) subset (**a**) and the participants of East-Asian ancestry from the Biobank Japan (BBJ) cohort (**b**). The dashed orange line indicates the suggestive significance threshold at P = 1 × 10⁻⁴.

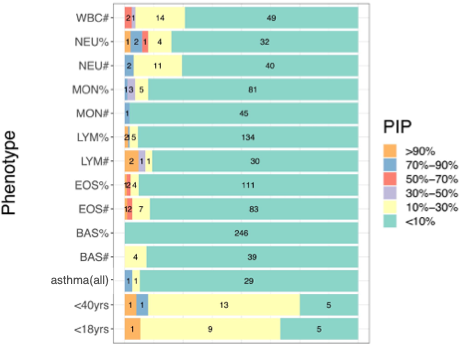

**Figure S8. Full summary of statistical fine-mapping results in UKB-European (UKB-EUR) participants using the SuSiE algorithm**. The figure shows the distribution of posterior inclusion probabilities (PIPs) for variants included in all 95% credible sets (CS95s) across phenotypes. For each phenotype, the proportion of variants falling within specified PIP ranges (see key) is displayed.

EUR SuSIE Credible Sets

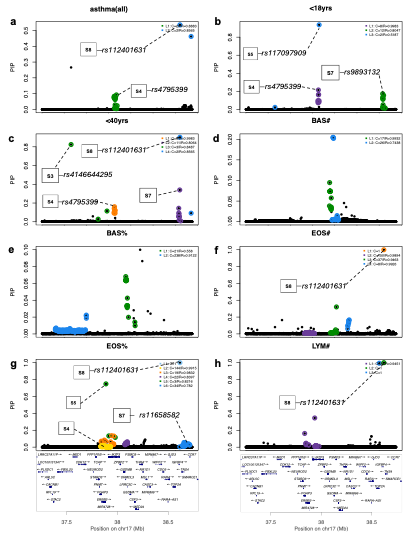

EUR SuSIE Credible Sets continued

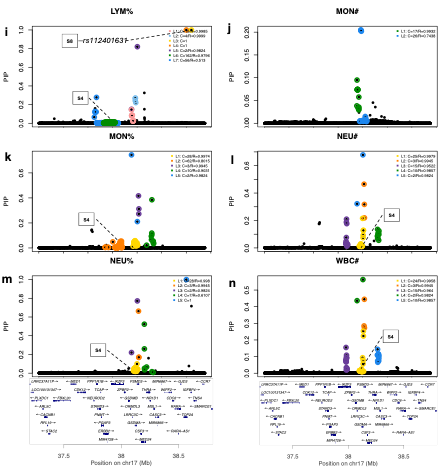

Non-EUR SuSIE Credible Sets

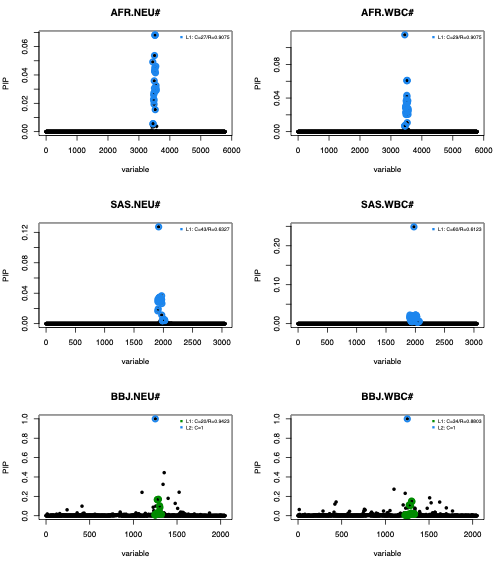

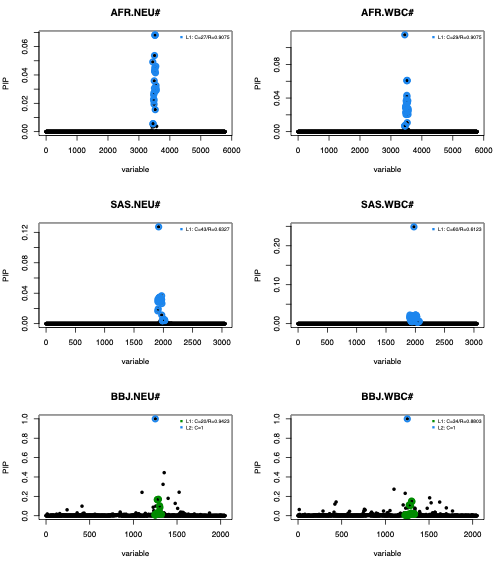

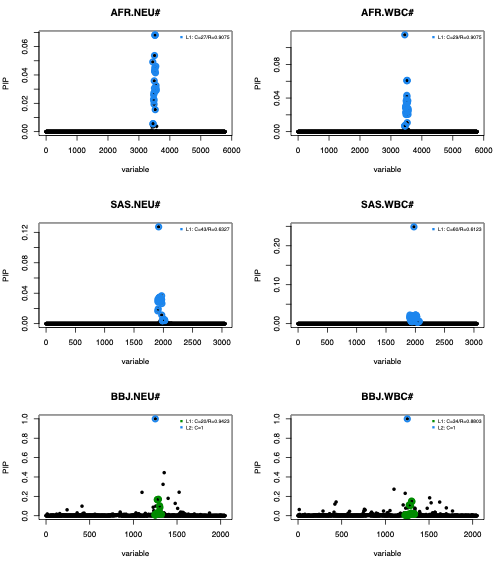

**t**

**s**

**r**

**q**

**p**

**o**

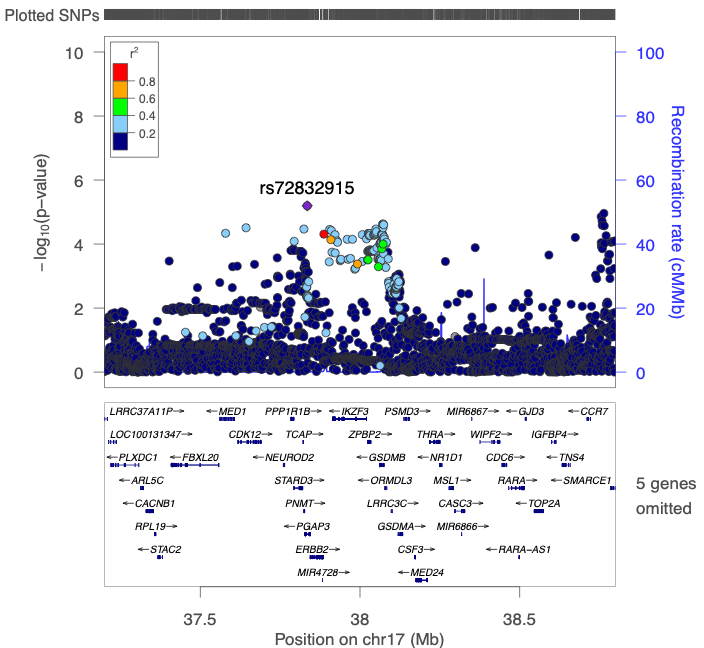

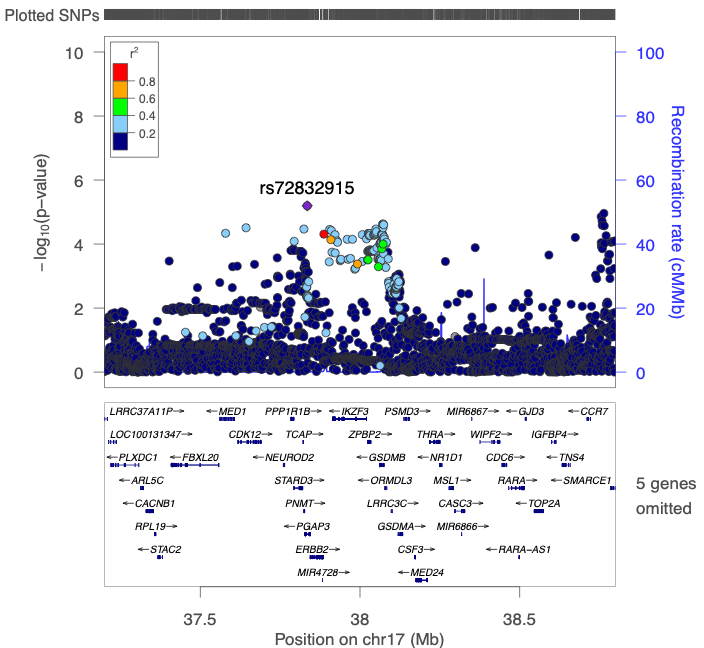

**Figure S9. Regional plots of asthma phenotypes and white blood cell traits in UKB-European and non-European populations showing 95% credible sets identified using the SuSiE algorithm**. Each plot displays variants within the study region, with chromosomal position on the x-axis and posterior inclusion probability (PIP) for causality on the y-axis. Panels (**a**-**n**) correspond to asthma phenotypes or white blood cell (WBC) traits in UKB-European (UKB-EUR) participants, while panels (**o**-**t**) represent WBC traits in non-European (non-EUR) populations. Each colour represents a distinct 95% credible set (CS95). Variants shaded in black without a coloured outline were not assigned to any CS95. Credible sets are numbered L1 to Ln, where n is the total number of credible sets identified. For each CS95, the number of variants (C) and the mean pairwise LD (r²) among variants are shown in the top right corner. Credible sets containing asthma signals are annotated with rsID’s if the GCTA-COJO variant is the lead (has the highest PIP in the credible set); otherwise, only the signal number is shown, marked with dashed lines.

**a**	Signal4--rs4795399 LD proxies.

EUR

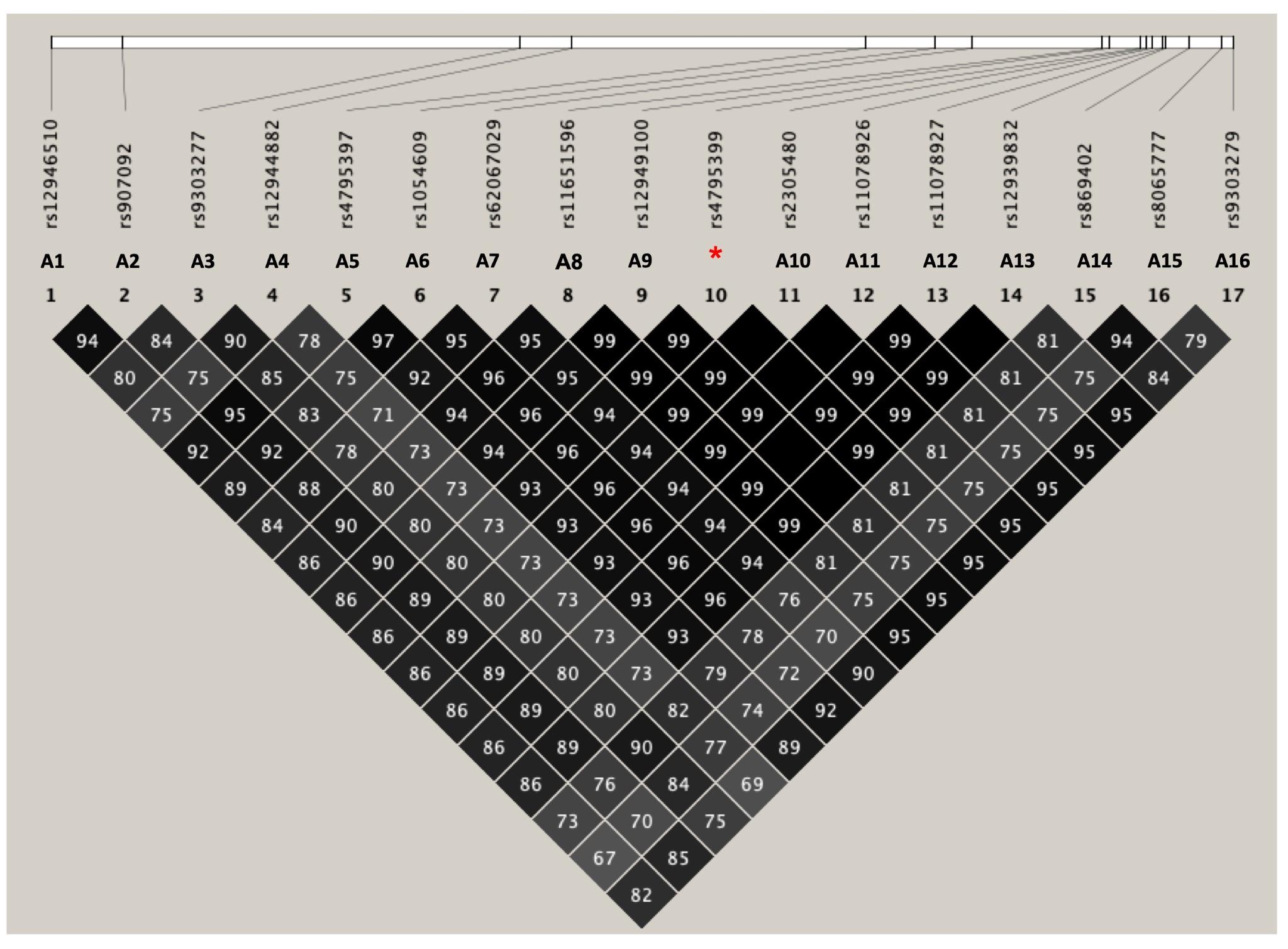

AFR	SAS	EAS

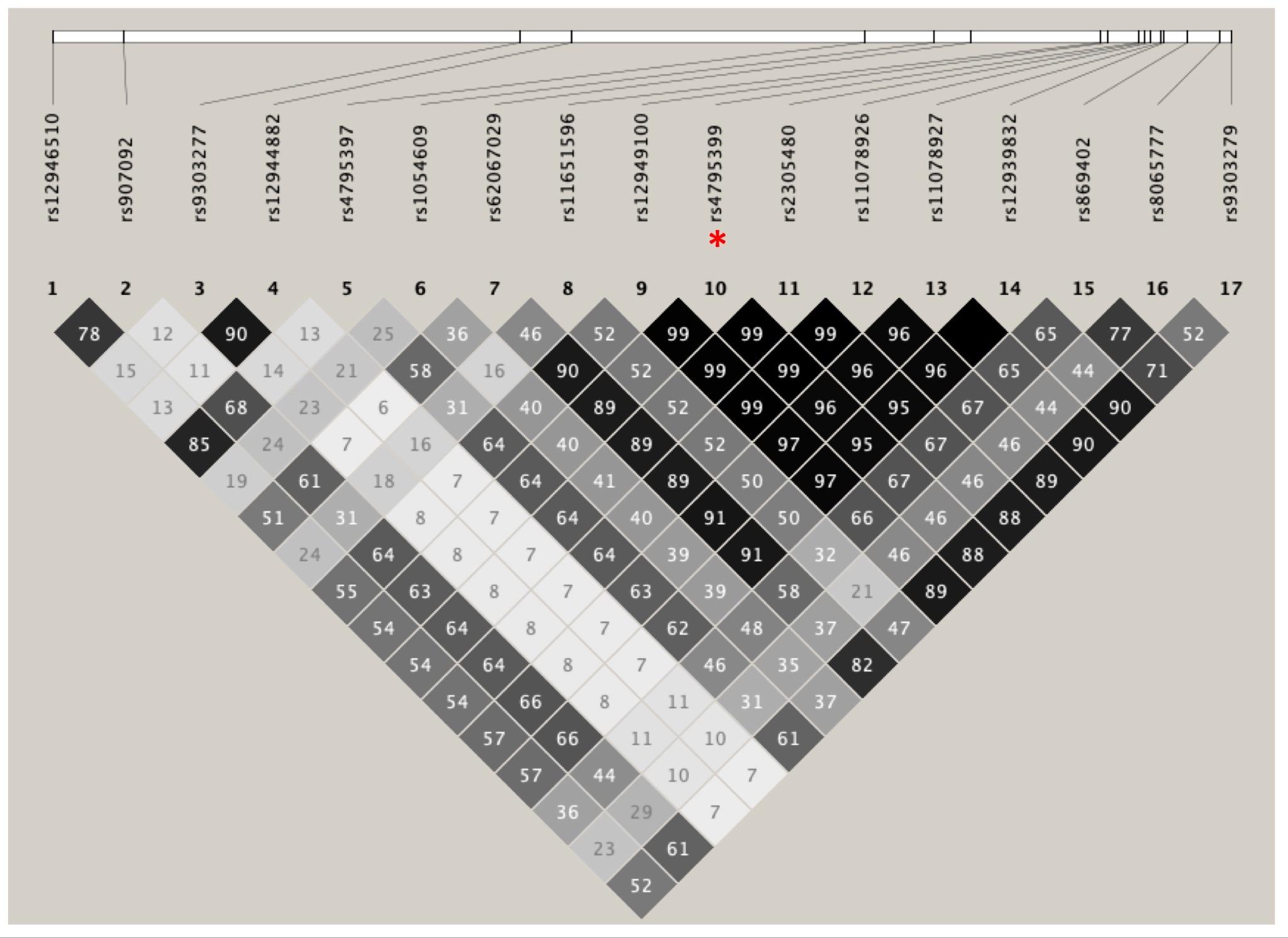

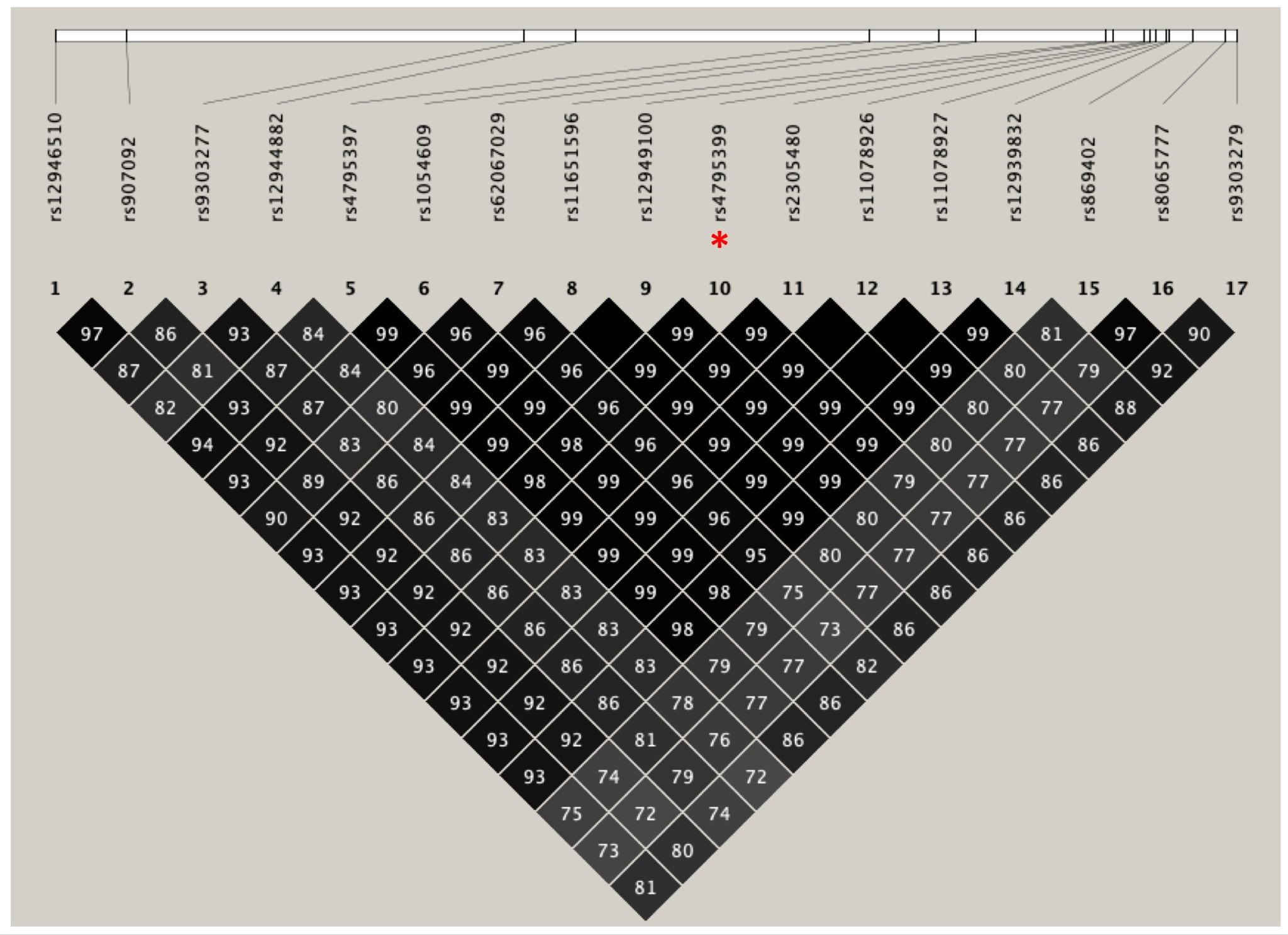

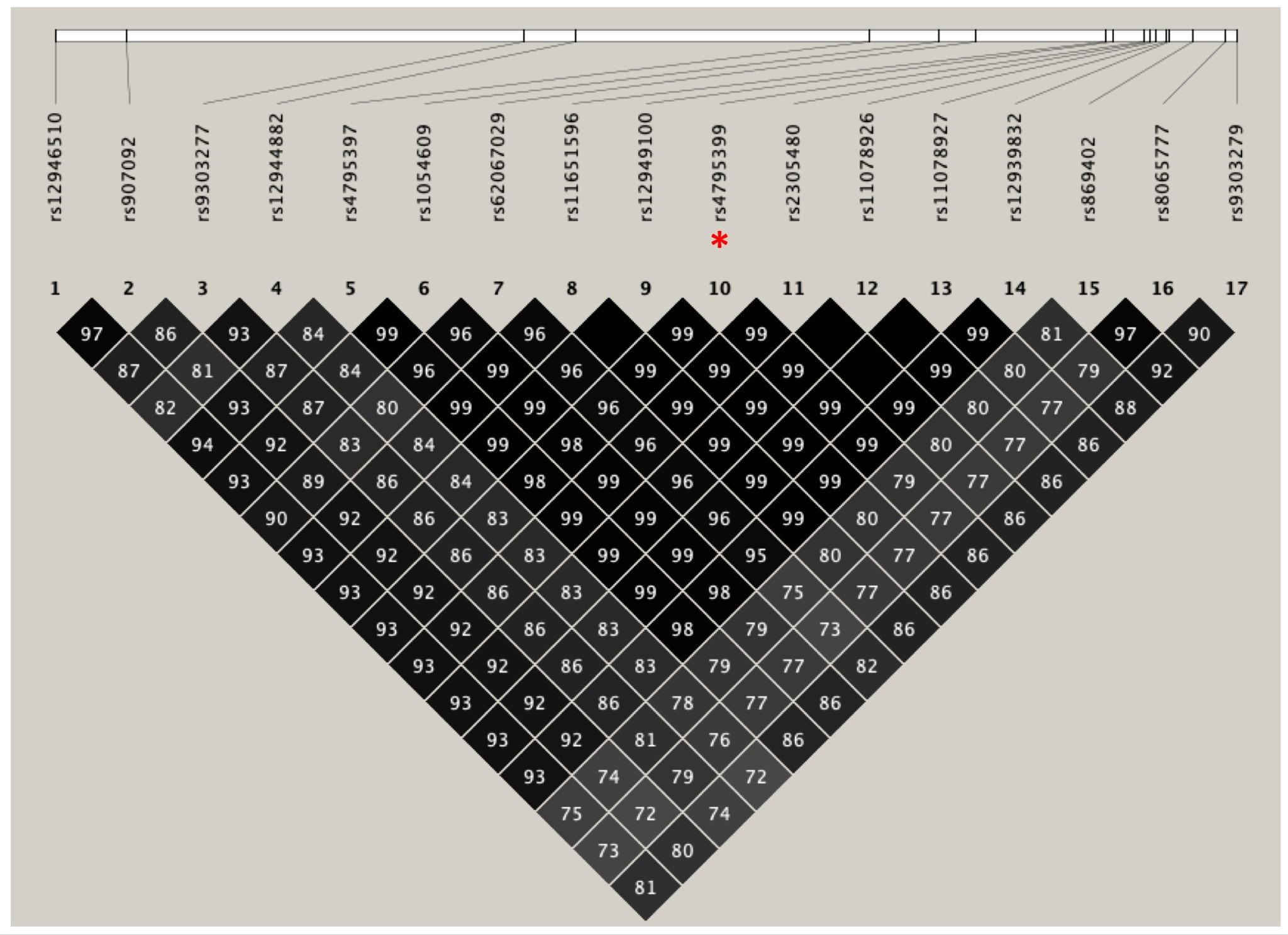

**b** Signal7--rs11658582 LD proxies

EUR	AFR

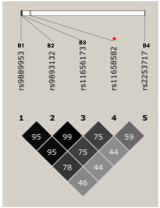

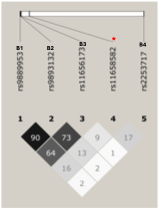

SAS	EAS

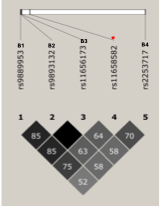

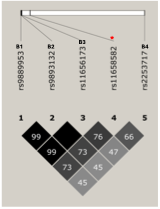

**Figure S10. Pairwise linkage disequilibrium (LD) between asthma signals and their LD proxies across study populations**. Asthma signals with more than two identified LD proxies in a given analysis are shown. Panels (**a**) and (**b**) display LD proxies for S4—rs4795399 and S7—rs11658582, respectively (both indicated with an asterisk in their respective plots). Proxies are ordered by base pair position (hg19). In panel (**a**), for S4–rs4795399: A1 and A9 were identified as independent signals by GCTA-COJO in the non-European meta-analysis (non-EUR META) and the full meta-analysis (META), respectively (see Additional file2: Tables S9 and S10). A3 and A7 were identified as independent signals for lymphocyte count (LYM#) and white blood cell count (WBC#) in BBJ, and for WBC# in UKB-European (UKB-EUR), respectively (Additional file2: Table S12). A2 and A4 were identified as top signals in the colocalization analysis for asthma adjusted for sex and age at baseline (asthma_all_) and LYM# in the SAS and BBJ populations, respectively (Table S19). A8–A13 were consistently grouped within the same 95% credible sets (CS95s) with S4--rs4795399 in SuSiE analyses for <18 years, <40 years, asthma_all_, and monocyte percentage (MON%) in UKB-EUR (Table S14); and for multi-trait colocalization (HyPrColoc) involving <18 years, <40 years, and LYM# (Additional file2: Table S22). A5, A14, A15, and A16 were identified through colocalization analysis for asthma–WBC trait pairs via COLOC-SuSiE for LYM#, MON%, NEU#, and WBC# in UKB-EUR (Table S20). In panel (b), for S7--rs11658582: B1, B2, and B4 were identified as independent signals in UKB-EUR using GCTA-COJO for LYM#, <18 years, and in the META analysis, respectively (see Additional file2: Tables S12, S5, and S9 respectively). B3 was prioritized by SuSiE as the proxy with the highest posterior inclusion probability (0.30) in the <18 years CS95 for signal 7 (see Fig. 3 and Additional file 2: Table S14).In panel (**b**), for S7—rs11658582: B1, B2, and B4 were identified as independent signals in UKB-EUR using GCTA-COJO for LYM#, <18 years, and in the META analysis, respectively (see Additional file2: Tables S12, S5, and S9 respectively). B3 was prioritized by SuSiE as the proxy with the highest posterior inclusion probability (0.30) in the <18 years CS95 for signal 7 (see Fig. 3 and Additional file 2: Table S14).

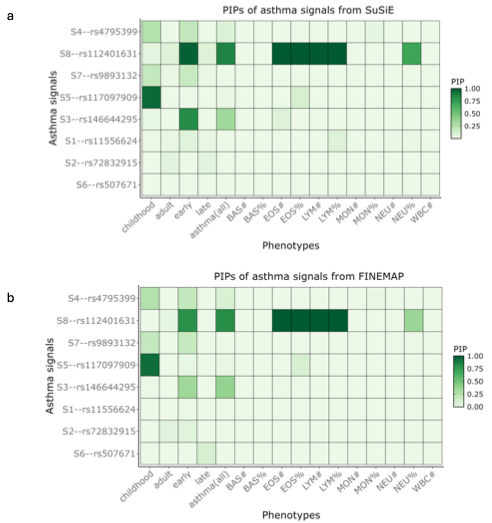

**Figure S11. Comparison of the Posterior Inclusion Probabilities (PIPs) for asthma signals between SuSiE and FINEMAP algorithms.** Heatmaps displaying the PIPs of asthma signals from statistical fine-mapping across asthma phenotypes and WBC traits. Signals are ordered according to the prioritization strategy outlined in Figure 3. Panel (**a**) above, shows PIPs obtained using SuSiE, while panel (**b**) below displays corresponding PIPs from FINEMAP. The comparison highlights agreement between the two fine-mapping approaches.

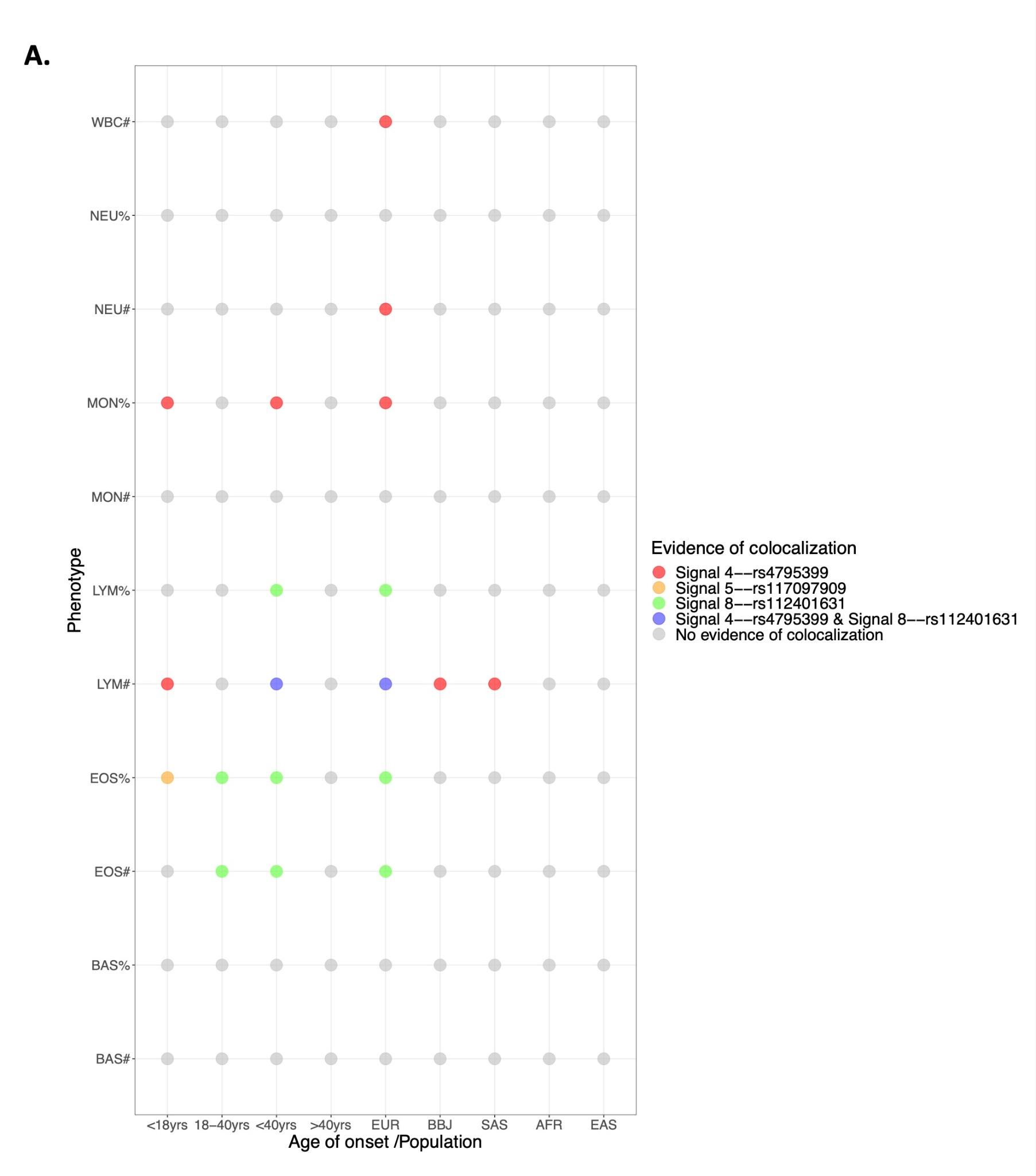

**a**

Signal 4--rs4795399

**b**

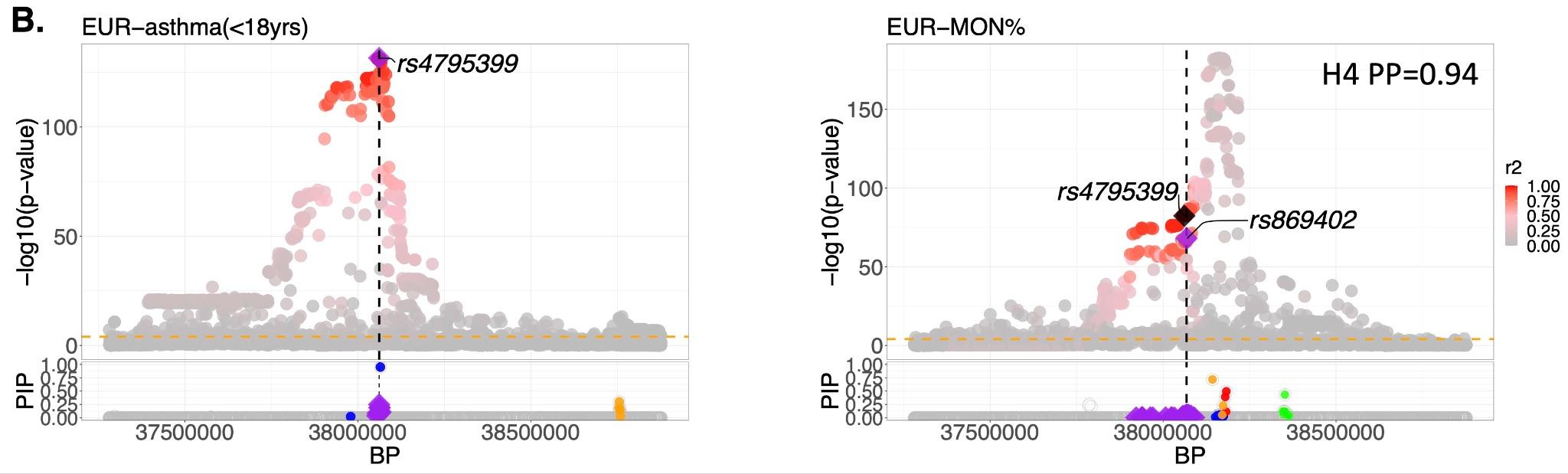

**c**

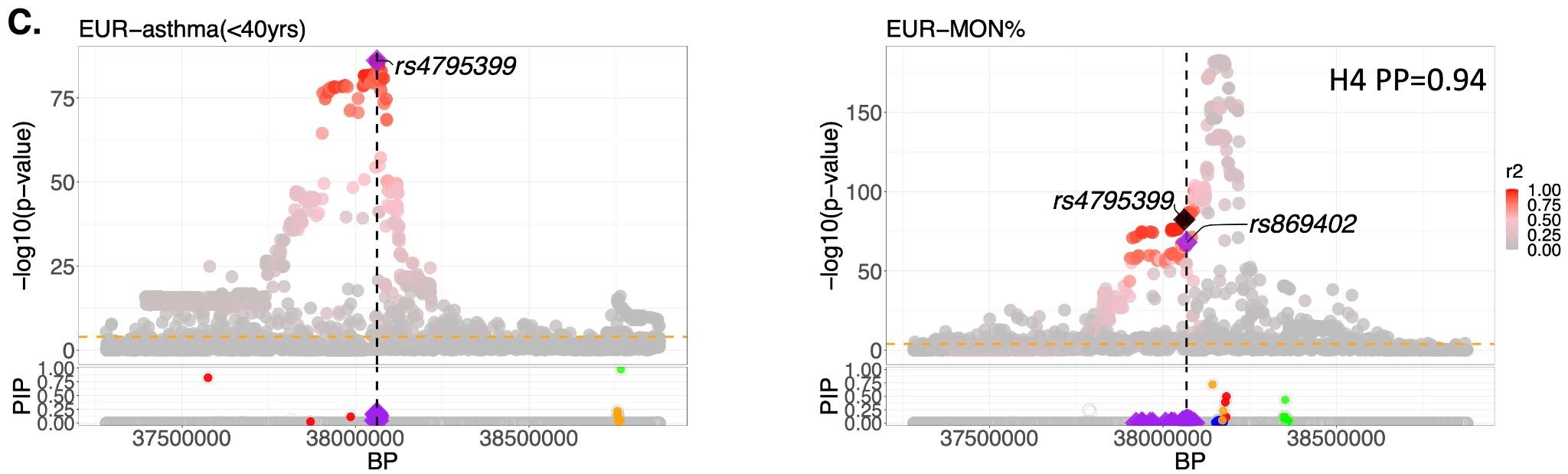

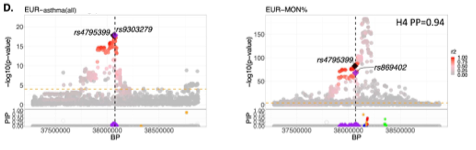

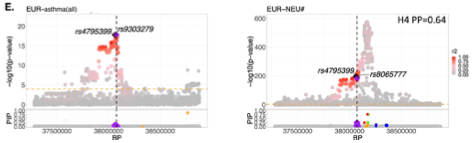

**e**

**d**

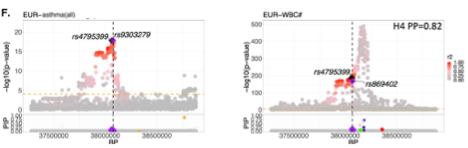

**f**

Signal 5--rs117097909

**g**

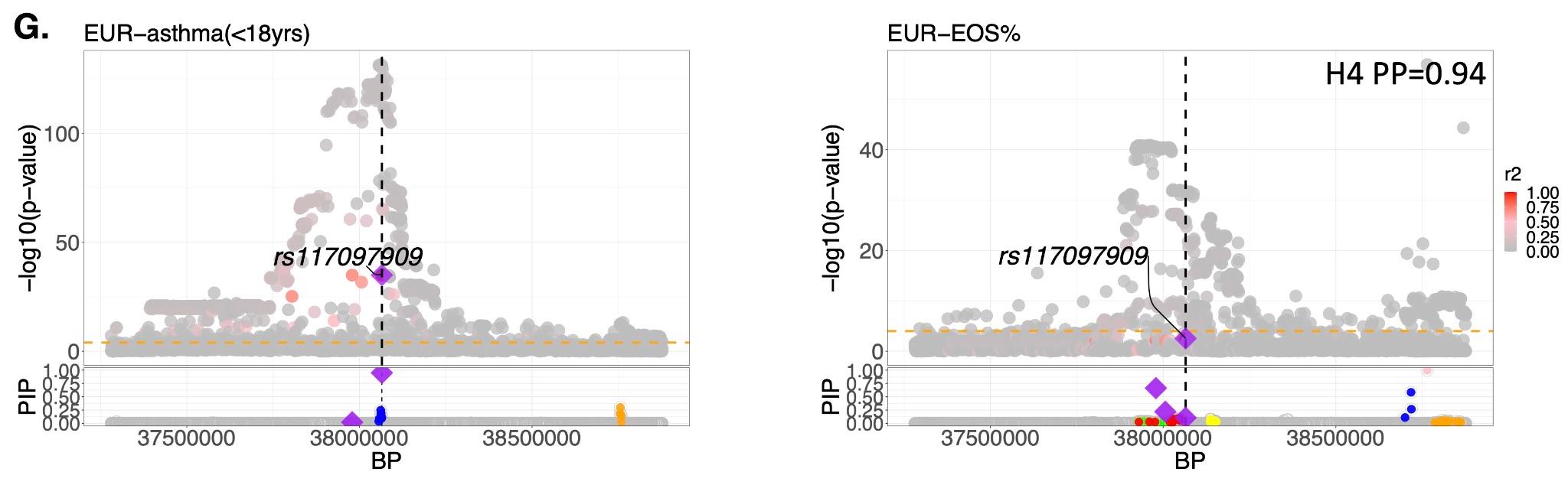

Signal 8--rs112401631

**h**

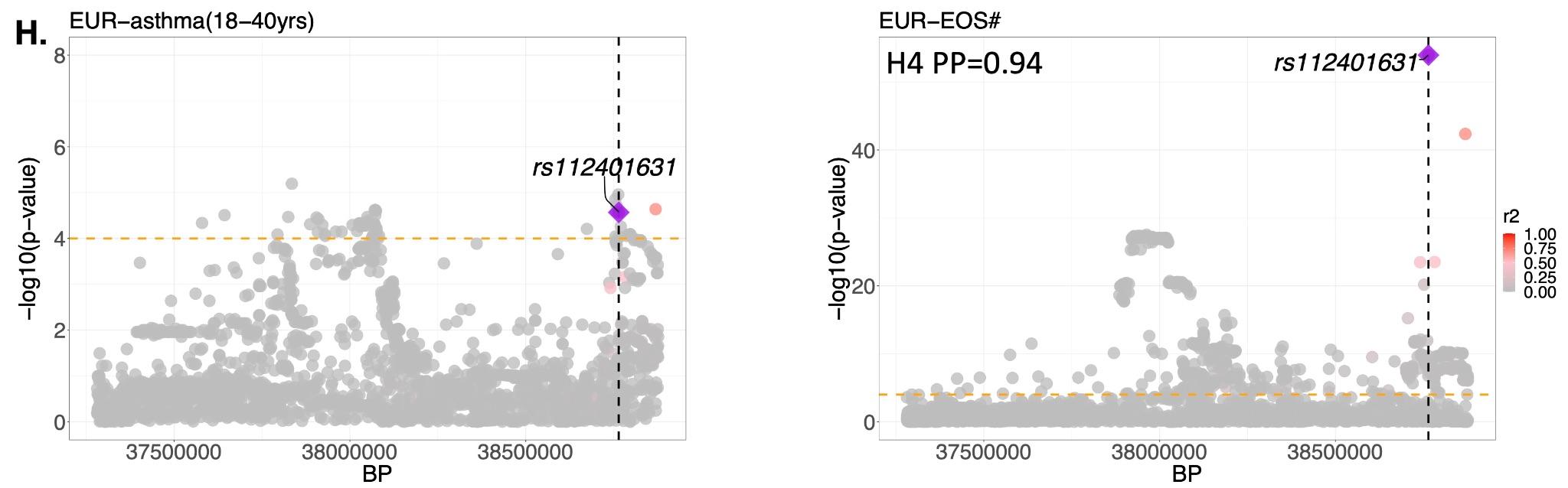

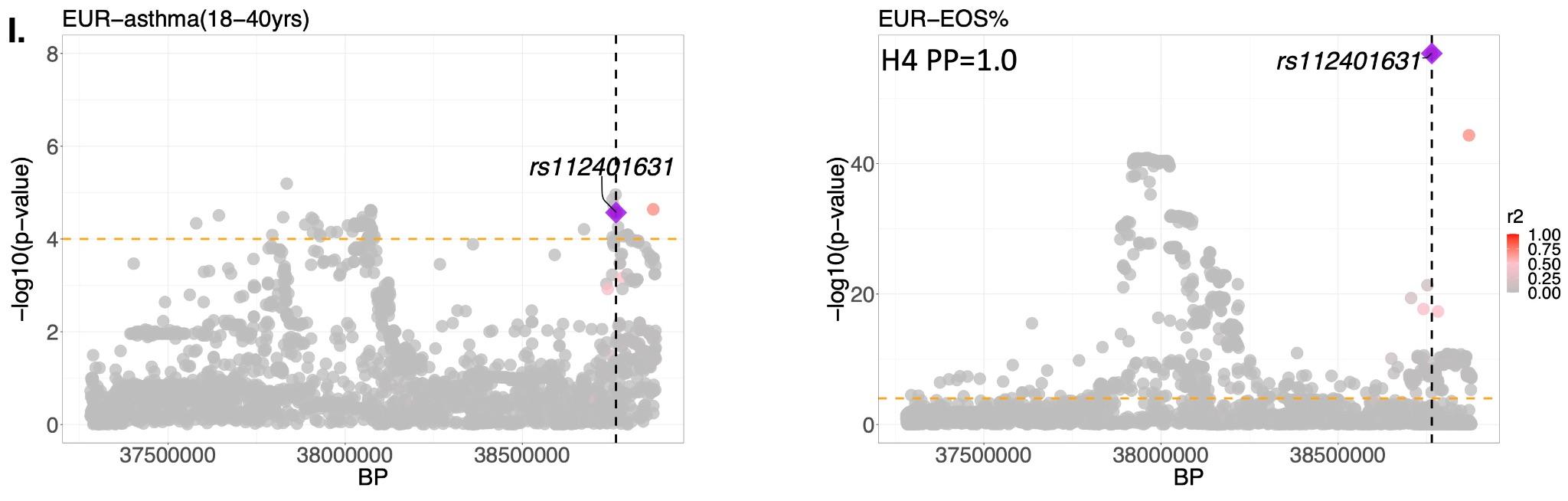

**i**

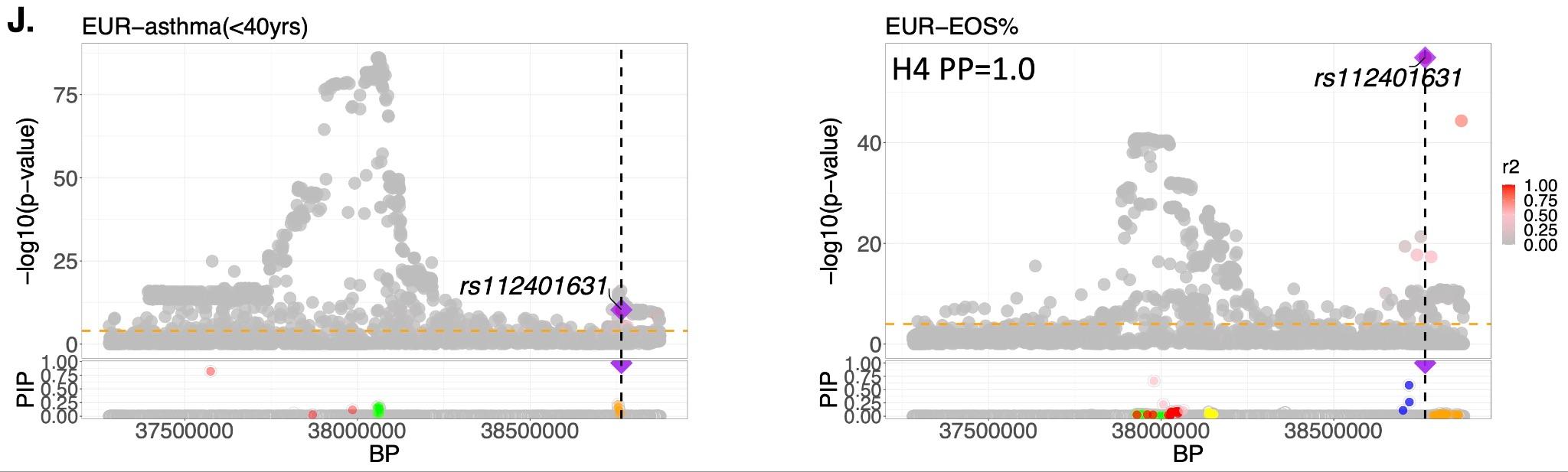

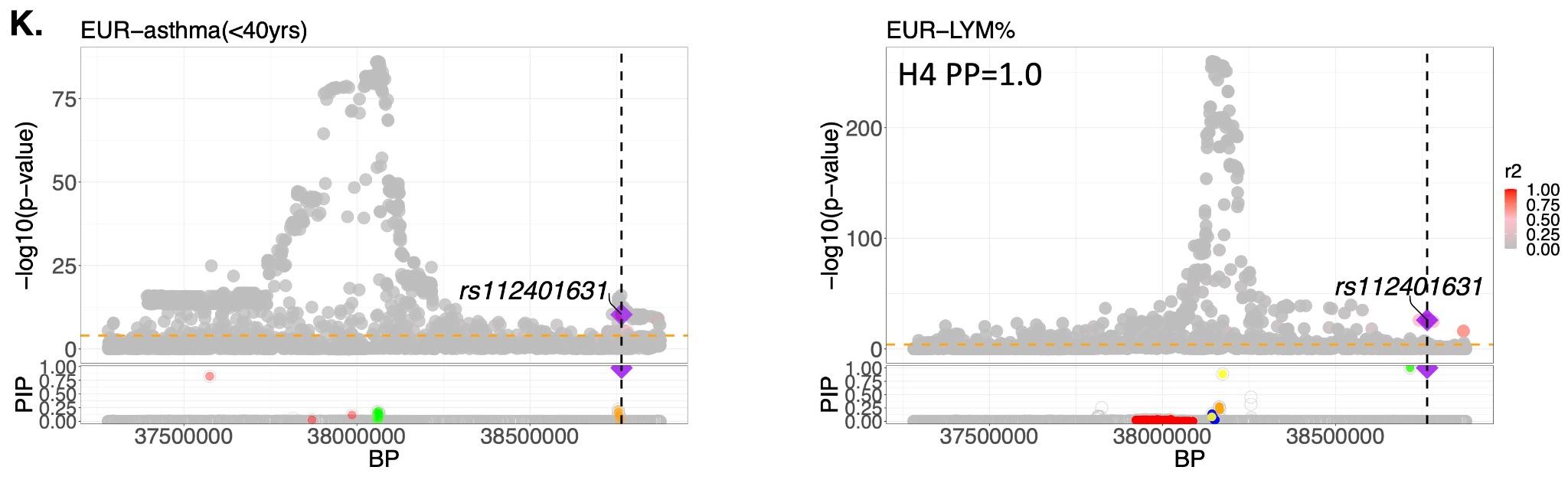

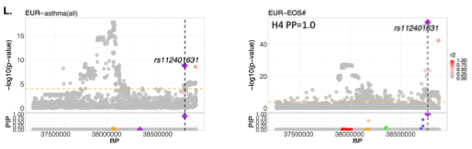

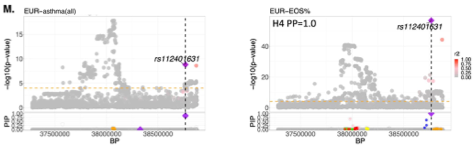

**m**

**l**

**j**

**k**

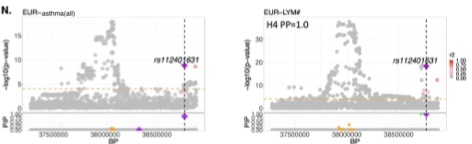

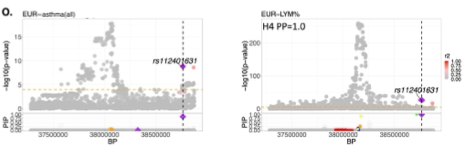

**o**

**n**

Signal 4--rs479539 & Signal 8--rs112401631

**p**

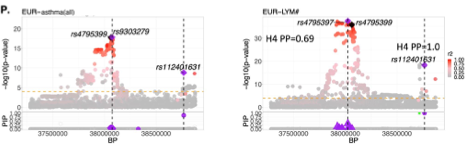

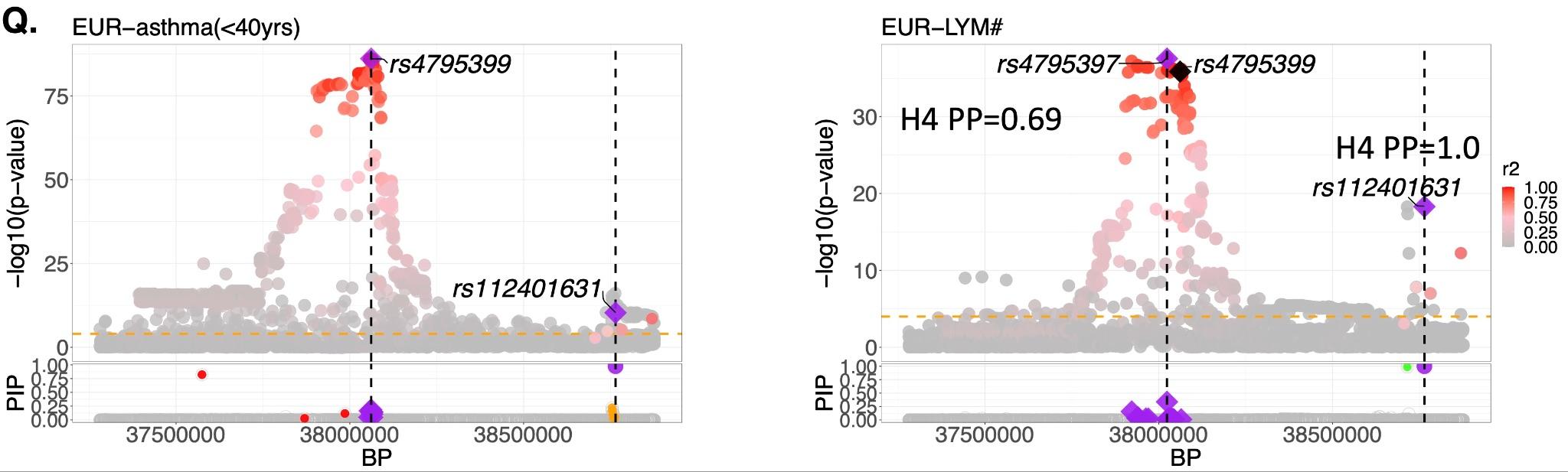

**q**

#### **Figure S12. Colocalization of causal variants between asthma and white blood cell traits.**

(**a**) Summary and (**b**-**q**) regional plots for variant–trait pairs showing colocalization evidence, based on COLOC-SuSiE analyses. Independent signals per trait were defined, and colocalization between SuSiE-generated 95% credible sets across asthma and WBC trait-pairs was assessed. Trait-pair combinations favouring shared causal variants (H4 PP > 0.6) are shown. In each regional plot, the H4 PP value is displayed on the WBC trait panel. The lead variant from the credible set (highest colocalization probability) is marked by a purple diamond and dashed vertical line; nearby variants are shaded by linkage disequilibrium (r²). In plots (**p**-**q**), purple diamonds and circles represent credible set variants for Signal 4 and Signal 8, respectively. The dashed orange line denotes the suggestive significance threshold (P = 1 × 10⁻⁴). Summary colocalization results across WBC traits and asthma age-of-onset strata are shown in (**a**). Regional plots (**b**-**q**) show asthma–WBC trait pairs for three asthma signals: S4--rs4795399 (**b**-**f**), S5--rs117097909 (**g**), and S8--rs112401631 (**h**-**o**), with S4 and S8 jointly colocalizing for lymphocyte count (**p**-**q**). For Signal 4, evidence of colocalization was observed without a distinct lead variant due to strong linkage disequilibrium among associated markers; S4--rs4795399 is depicted as a black diamond in the regional plots. Posterior inclusion probability (PIP) plots are shown for cases with credible sets identified for both traits.

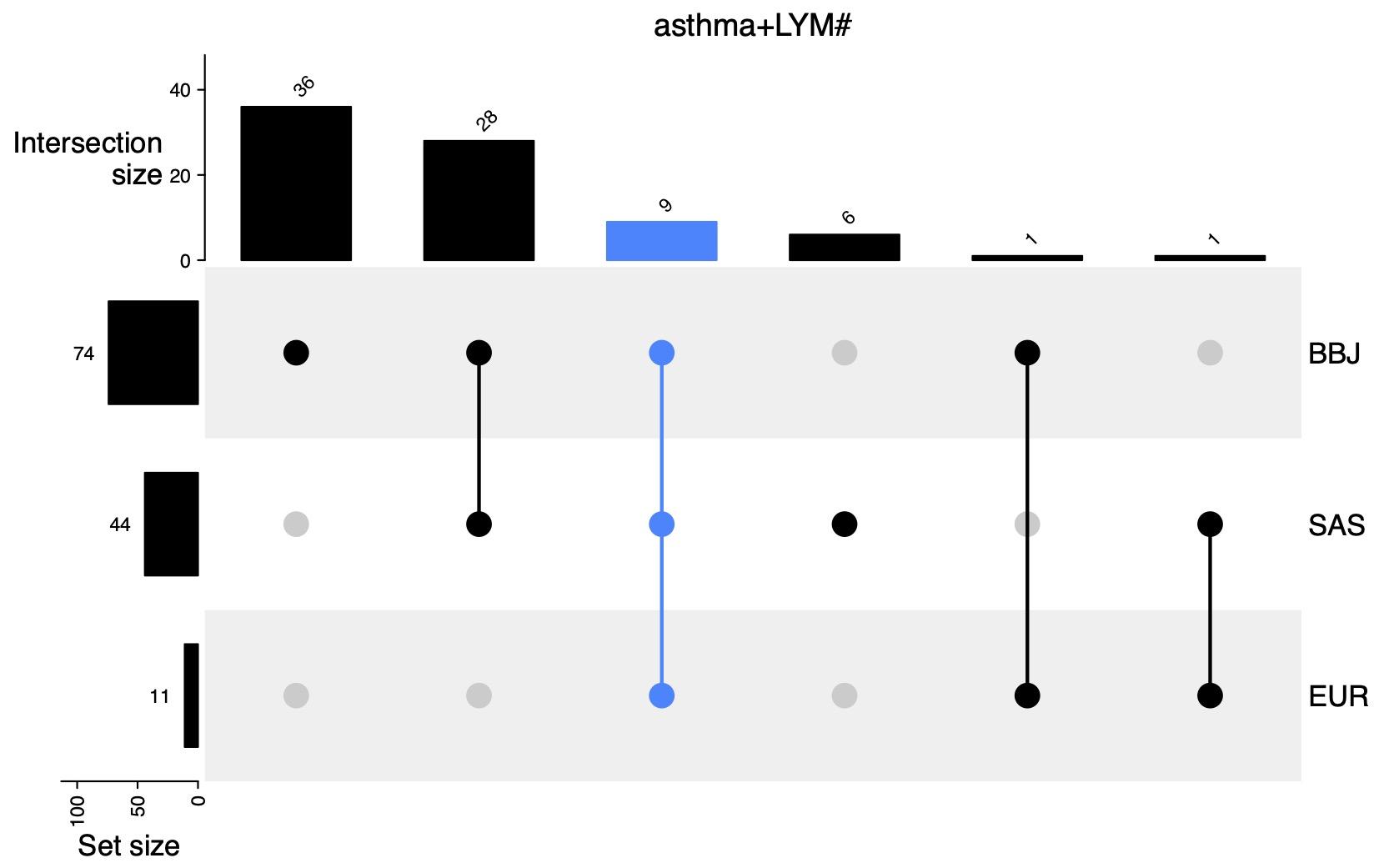

**Figure S13. Signal 4 proxies for asthma-lymphocyte count (LYM#) colocalization under the single causal variant assumption (COLOC).** In the EUR, the union of variants (11) across three CS95s for: <18yrs+LYM#, <40yrs+LYM# and asthma_all_ are shown (H4 PP > 0.6), while variants in the CS95s for asthma_all_ in the SAS (44) and BBJ (74) are shown. Presence of overlapping variant sets across two or more populations, or unique sets to a particular population, are displayed as black circles with vertical line connectors across populations in the bottom panel. Frequency bins on top (intersection size) and to the left (set size) show the total counts of variants for each overlapping CS95s, and per population, respectively. The 9 variants (Signal 4 proxies) that occur in the CS95s of all 3 populations are as follows: rs4795399, rs8069176, rs11078926, rs12939832, rs2305480, rs11078927, rs11651596, rs11078928, rs12949100. For test statistics see Additional file2: Tables S18 and S19.

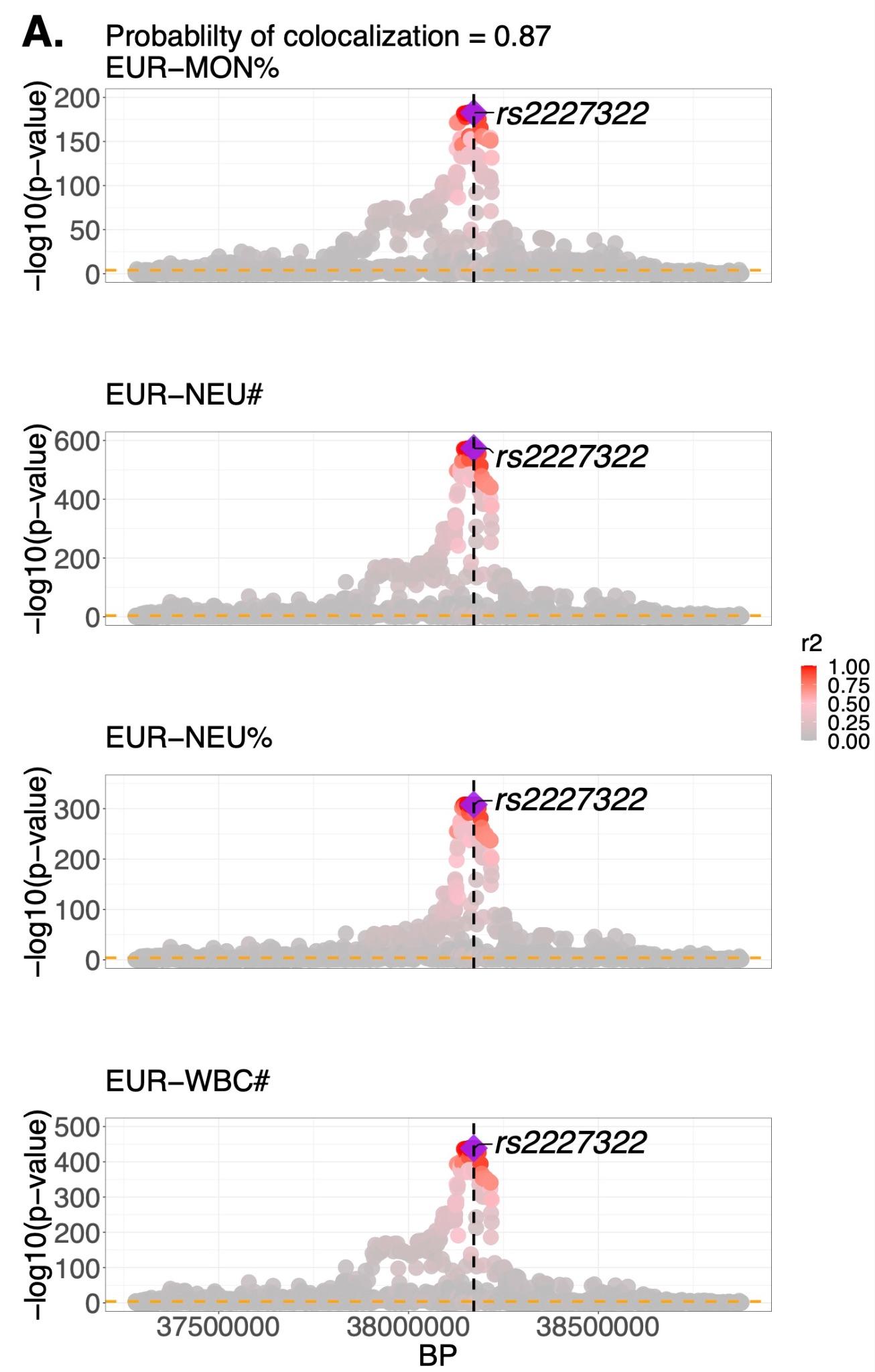

**Figure S14. Evidence of colocalization across multiple white blood cell traits in the UKB-European population.** Colocalization analysis across four white blood cell (WBC) traits: monocyte percentage (MON%), neutrophil count (NEU#), neutrophil percentage (NEU%), and total white blood cell count (WBC#)-was performed using HyPrColoc in UKB-European (UKB-EUR) participants. The probability of colocalization was 0.87, with lead variant rs2227322 identified. Variants are shaded according to linkage disequilibrium (LD, r²) with the lead variant (see key). The dashed orange line indicates the suggestive significance threshold at P = 1 × 10⁻⁴. See Additional file2: Table S22 for full multi-trait colocalization results.

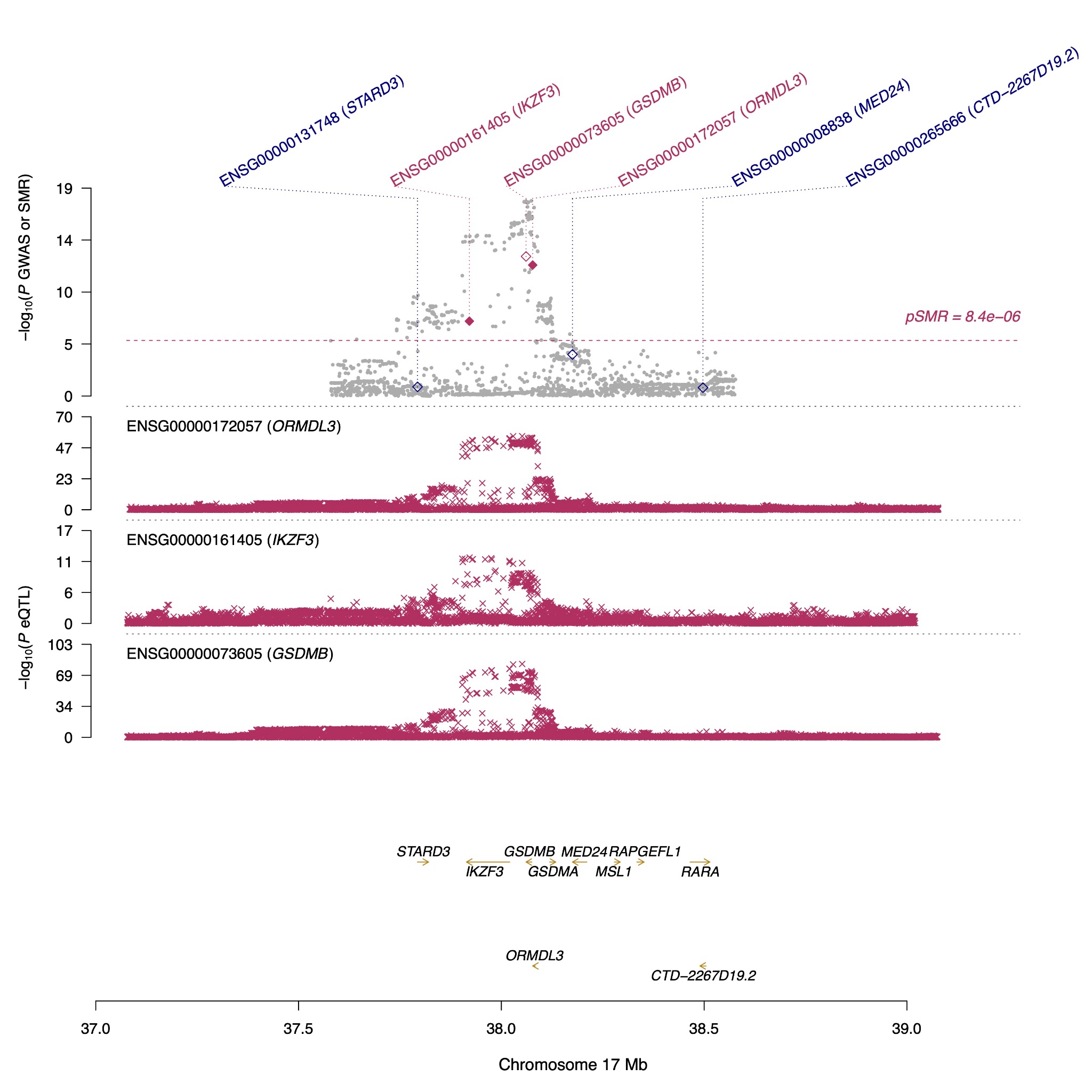

**Figure S15. Significant genes identified from Summary-data-based Mendelian Randomization (SMR) and Heterogeneity in Dependent Instruments (HEIDI) analysis in whole blood.** The plot highlights significant genes (*ORMDL3* and *IKZF3*) mapping to Signal 4 variants, demonstrating pleiotropic associations with asthma adjusted for sex and age at baseline (asthma_all_) in UKB-European (UKB-EUR) participants, using Genotype Tissue Expression Project (GTEx) eQTL data. In the top plot, grey dots represent -log₁₀(P-values) for SNPs from the GWAS of asthma_all_. Rhombuses represent -log₁₀(P-values) for probes from the SMR test in whole blood: solid maroon rhombuses indicate probes that pass the HEIDI test (*P*_HEIDI_ ≥ 0.05), hollow maroon rhombuses indicate probes that fail the HEIDI test, and blue hollow rhombuses indicate probes that were not significant in SMR analysis. The middle plot shows eQTL association results. The bottom plot indicates the genomic locations of genes tagged by the SMR probes. The SMR P-values for *ORMDL3* and *IKZF3* are 2.8 × 10⁻¹⁴ and 1.5 × 10⁻⁷, respectively.

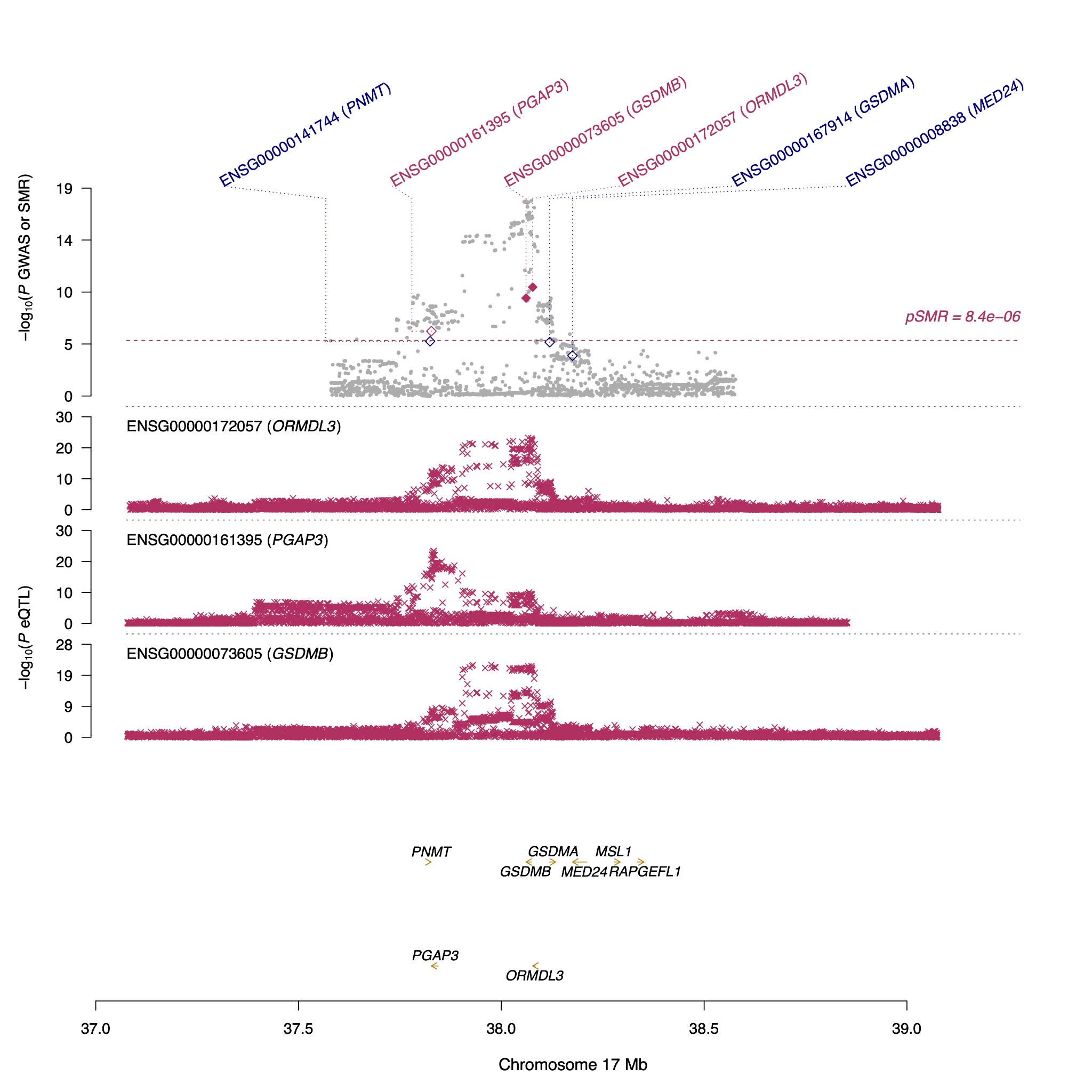

**Fig. S16. Significant genes identified from Summary-data-based Mendelian (SMR) Randomization and Heterogeneity in Dependent Instruments (HEIDI) analysis in the lung.** The plot highlights significant genes (*ORMDL3* and *GSDMB*) mapping to Signal 4 variants, demonstrating pleiotropic associations with asthma adjusted for sex and age at baseline (asthma_all_) in UKB-European (UKB-EUR) participants, using Genotype Tissue Expression Project (GTEx) lung eQTL data. In the top plot, grey dots represent -log₁₀(P-values) for SNPs from the GWAS of asthma_all_. Rhombuses represent −log₁₀(P-values) for probes from the SMR test in the lung: solid maroon rhombuses indicate probes that pass the HEIDI test (*P*_HEIDI_ ≥ 0.05), hollow maroon rhombuses indicate probes that fail the HEIDI test, and blue hollow rhombuses indicate probes that were not significant in SMR analysis. The middle plot shows eQTL association results. The bottom plot indicates the genomic locations of genes tagged by the SMR probes. The SMR P-values for *ORMDL3* and *GSDMB* are 9.0 × 10⁻¹⁰ and 1.1 × 10⁻⁸, respectively.

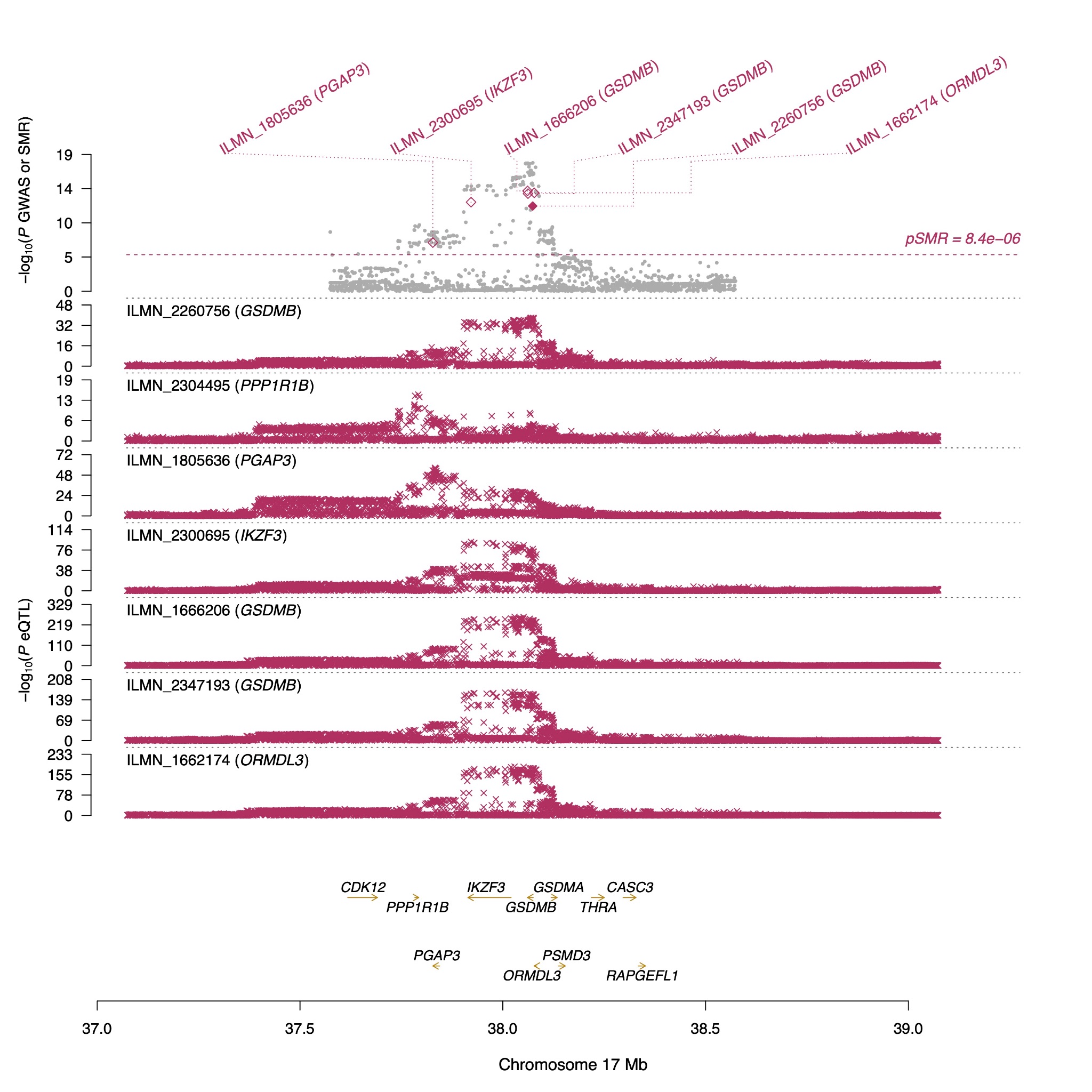

**Figure S17. Significant gene identified from Summary-data-based Mendelian Randomization (SMR) and Heterogeneity in Dependent Instruments (HEIDI) analysis in peripheral blood.** The plot highlights the gene GSDMB, mapping to Signal 4 variants, demonstrating pleiotropic association with asthma adjusted for sex and age at baseline (asthma_all_) in UKB-European (UKB-EUR) participants, using Consortium for the Architecture of Gene Expression (CAGE) eQTL data. In the top plot, grey dots represent -log₁₀(P-values) for SNPs from the GWAS of asthma_all_. Rhombuses represent −log₁₀(P-values) for probes from the SMR test in peripheral blood: solid maroon rhombuses indicate probes that pass the HEIDI test (*P*_HEIDI_ ≥ 0.05), hollow maroon rhombuses indicate probes that fail the HEIDI test, and blue hollow rhombuses indicate probes that were not significant in SMR analysis. The middle plot shows eQTL association results. The bottom plot indicates the genomic location of genes tagged by the SMR probes. The SMR P-value for *GSDMB* is 1.9 × 10⁻¹².
